# Sex differences in TB treatment outcomes: Retrospective cohort study and meta-analysis

**DOI:** 10.1101/2021.04.26.21256155

**Authors:** Vignesh Chidambaram, Nyan Lynn Tun, Marie Gilbert Majella, Jennie Ruelas Castillo, Samuel K. Ayeh, Amudha Kumar, Pranita Neupane, Ranjith Kumar Sivakumar, Ei Phyo Win, Enoch J. Abbey, Siqing Wang, Alyssa Zimmerman, Jaime Blanck, Akshay Gupte, Jann-Yuan Wang, Petros C. Karakousis

## Abstract

**Rationale:** Although the incidence of tuberculosis (TB) is higher in males compared to females, the relationship of sex with TB treatment outcomes has not been adequately studied.

**Objectives and Methods:** We performed a retrospective cohort study and a systematic review and meta-analysis of observational studies during the last 10 years to assess the sex differences in clinical and microbiological outcomes in tuberculosis.

**Measurements and Main Results:** In our cohort of 2,894 patients with drug-susceptible pulmonary TB (1,975 males and 919 females), males had higher adjusted hazards of mortality due to all causes (HR 1·43,95%CI 1.03-1.98) and infections (HR 1.70, 95%CI 1.09-2.64) at 9 months and higher adjusted odds ratio for sputum culture (OR 1.56,95%CI 1.05-2.33) and similar odds ratio for smear positivity (OR 1.27, 0.71-2.27) at 2 months compared to females. Among 7896 articles retrieved, 398 articles were included in our systematic review with a total of 3,957,216 patients. The odds of all-cause mortality was higher in males compared to females in the pooled unadjusted (OR 1.26, 95%CI 1.19-1.34) and adjusted (OR 1.31, 95%CI 1.18-1.45) analyses. Relative to females, males had higher pooled odds of sputum culture (OR 1.44,95% CI 1.14-1.81) and sputum smear (OR 1.58,95%CI 1.41-1.77) positivity at the end of the intensive phase, and upon completion of treatment.

**Conclusions:** During TB treatment, males have higher all-cause-, infection- and TB-related mortality, as well as higher rates of sputum smear and culture positivity, both after the intensive phase and at the completion of TB treatment, after adjusting for confounding factors.

## Introduction

The World Health Organization reported nearly 10 million new cases of active tuberculosis(TB) in 2019(1). Although *Mycobacterium tuberculosis*(*Mtb*) infects both sexes and all age groups, adult men have a 1.8-fold higher incidence of active TB disease(1, 2). These sex disparities have been reported irrespective of geographical locale(3), and studies have shown that the ratio of prevalence to notification of TB is nonetheless higher in men(4, 5).

Many potential medical and cultural confounding factors such as the increased prevalence of diabetes, alcohol use, and smoking among males, and decreased access to health care among females may account for sex differences in TB (1, 3). Clinical studies have reported that men had higher sputum bacterial loads and greater severity of TB-related lung disease on chest imaging than women(6, 7). A few population-based studies have shown male patients to have poor outcomes following tuberculosis(8, 9). These clinical observations are supported by animal model data showing more extensive lung pathology, higher lung bacterial burdens, and accelerated mortality in males relative to females following *Mtb* infection(10, 11).

Although prior clinical observational studies have highlighted sex differences in TB disease, the findings are often unclear and inconsistent because of the inherent heterogeneity of study populations and the level of care in low-income countries, as well as due to the lack of disaggregated data related to sex in many studies. We analyzed a retrospective cohort of Taiwanese patients with drug-susceptible pulmonary TB to better understand the role of sex on mortality and sputum microbiological status after TB treatment initiation, after adjusting for common confounders. We also performed a systematic review and meta-analysis on this topic, given the lack of generalizability from individual cohort studies, and to better assess the impact of other important parameters, including HIV-coinfection status, site of TB involvement, level of drug resistance, duration of follow up, and study country on the effect of sex differences.

## Methods

### Retrospective cohort study

#### Study design and population

Our retrospective cohort consisted of adult patients (age > 18 years) with culture-confirmed, drug-susceptible pulmonary TB, treated according to the American Thoracic Society guidelines(12), enrolled at the National Taiwan University Hospital (NTUH) in Taipei from 2000 to 2016(13). There were no exclusion criteria. The institutional review boards at Johns Hopkins University and NTUH approved the study.

Baseline characteristics on age, sex, body mass index (BMI), smoking status, alcohol abuse, comorbidities, HIV co-infection, baseline sputum smear acid-fast bacilli (AFB) positivity, and presence of cavity on chest radiography were collected from the NTUH database at the time of TB diagnosis. The Charlson Comorbidity Index (CCI) was calculated from the parameters obtained from the database(14).

#### Exposure and Outcomes

To ascertain biological sex differences, we considered male sex as the exposure group, and female sex as the comparison group. The primary outcomes were all-cause and infection-related mortality at 9 months following ATT initiation. Infection-related mortality was a composite outcome of death due to pneumonia, sepsis, and TB. Secondary outcomes were positivity of sputum cultures and sputum smear AFB by microscopy at 2 months after ATT initiation.

#### Statistical analysis

Participant characteristics stratified by sex were compared using two-sided t-tests and Chi-square tests for continuous and categorical variables, respectively. Kaplan-Meier analysis and Cox Proportional Hazards regression were used to measure the association between sex and all-cause and infection-related mortality in separate models. Person-time at risk of outcome stratified by sex was calculated from the time of TB treatment initiation up to 9 months, or loss to follow-up or death, whichever occurred first. The association of sex with sputum culture and smear positivity at 2 months was analyzed using univariable and multivariable logistic regression. Potential confounders for multivariable analyses were identified by literature review and exploratory data analysis. Potential confounding factors included in the CCI were not adjusted for separately if CCI was included in the multivariable analytical model.

### Systematic Review

#### Search strategy and study selection

The systematic review was conducted according to the PRISMA guidelines(15). The literature searches were performed in PubMed, Embase, and Web of Science on August 15, 2020, using the search strategy detailed in the supplementary document (Section I). We captured both research articles and letters in the English language published in the last ten years. We included only articles published in peer-reviewed academic journals; conference abstracts were not included.

Studies were required to report sex-disaggregated data on at least one of the following outcomes on adult TB patients treated with multidrug anti-TB therapy (ATT): all-cause mortality, mortality due to TB, sputum AFB smear or culture positivity during or at the end of TB treatment, or ‘treatment success’ according to the WHO definitions for reporting TB outcomes(16). We included prospective and retrospective cohort studies and case-control studies. We excluded case reports, case series, and cross-sectional studies. We utilized the COVIDENCE platform for the systematic review(17). After removing the duplicates, the titles and abstracts of the retrieved articles were screened by at least two authors (VC, NT, AK, or PN) independently and the disagreements were resolved by VC. At least two authors (VC, NT, AK, or MM) independently performed the full-text screening, and the conflicts were resolved by VC.

#### Data extraction and quality assessment

At least two authors extracted data from the articles (VC, NT, MM, RK, SA, EW, EA, SW, or AZ) in the Qualtrics platform (18), and discrepancies were resolved by VC. Data on the country of study, funding source, patient comorbidities, site of TB involvement, pattern of resistance to ATT, HIV-TB co-infection, duration of treatment, and time points for outcomes were extracted. Data on treatment outcomes were extracted either as raw data or as pre-calculated effect sizes, namely odds ratio (OR), relative risk (RR), or hazard ratio (HR), along with 95% confidence interval (CI), as reported in the studies. RR for mortality in males compared to females was reported only in 7 studies, and were therefore converted to OR to facilitate pooling of the effect sizes(19). We performed quality assessment using the New-Castle Ottawa scale for observational studies(NOS)(20).

#### Data analysis

For each of the outcomes, we performed a meta-analysis of the OR using the random-effects model. We pooled HR separately for each of the outcomes. We considered a two-sided probability of < 0.05 as significant. We assessed heterogeneity using the I^2^ statistics. When the I^2^ was > 60%, we performed subgroup analyses and meta-regression (Detailed in the supplementary document – Section IIa) with respect to the TB-HIV co-infection status, resistance to ATT, and extra-pulmonary involvement, time of outcome assessment, study country’s income status classification according to World Bank(21), incidence of TB infection and HIV-TB co-infection(22). Publication bias was assessed by funnel plot and Egger’s test. Statistical analyses were performed using STATA/IC 16.0 software (StataCorp, College Station, Texas)(23). The study protocol is registered with PROSPERO (CRD42020219050).

## Results

### Retrospective cohort

In our cohort of 2894 patients with culture-confirmed, drug-susceptible pulmonary TB, 1975 (68.2%) were males, and 919 (31.8%) were females (Table 1). Males had a higher median age than females (68.9 years vs 58.2 years, p <0.001). A greater proportion of males had comorbidities, history of smoking, and alcohol abuse compared to females. Males had a greater proportion of cavitary disease (15.7% vs 11.1%; p<0.001) at diagnosis (Table 1). Baseline sputum AFB smear positivity rates were similar in both sexes.

**Table 1:** Characteristics of the study participants in the retrospective cohort from Taiwan stratified by sex (N=2894)

| Study Characteristics | Measure | Data available for (n) | Total (n=2894) | Males(n=1975) | Females(n=919) | p-value |
| --- | --- | --- | --- | --- | --- | --- |
| Age (years) | Median (IQR) | 2894 | 66.6 (49.1 – 77.8) | 68.9 (53.4– 78.8) | 58.2 (38.2– 75.4) | <0.001 |
| BMI | Mean (SD) | 2178 | 21.3 ± 4.2 | 21.4 ± 4.4 | 21.0 ± 3.6 | 0.049 |
| Ever smoker | No. (%) | 2296 | 930(40.5%) | 884(56.1%) | 46(6.4%) | <0.001 |
| Alcohol abuse | No. (%) | 2894 | 81(2.8%) | 79(4.0%) | 2(0.2%) | <0.001 |
| Diabetes mellitus | No. (%) | 2888 | 533(18.46%) | 403(20.4%) | 130(14.2%) | <0.001 |
| Hypertension | No. (%) | 2894 | 1052(36.4%) | 759(38.4%) | 293(31.9%) | 0.001 |
| CVD | No. (%) | 2894 | 352(12.2%) | 265(13.4%) | 87(9.5%) | 0.002 |
| COPD | No. (%) | 2894 | 451(15.6%) | 374(18.9%) | 77(8.4%) | <0.001 |
| Cancer | No. (%) | 2888 | 459(15.9%) | 369(18.7%) | 90(9.8%) | <0.001 |
| Cirrhosis | No. (%) | 2894 | 5(1.7%) | 43(2.2%) | 7(0.8%) | 0.007 |
| Transplant | No. (%) | 2894 | 26(0.9%) | 18(0.9%) | 8(0.9%) | 0.914 |
| HIV | No. (%) | 2894 | 65(2.3%) | 64(3.2%) | 1(0.1%) | <0.001 |
| CCI | Median (IQR) | 2894 | 4(2-6) | 4(2-6) | 3(1-5) | <0.001 |
| Sputum Smear AFB positivity at baseline | No. (%) | 2842 | 1215(42.75%) | 820(42.40%) | 395(43.50%) | 0.579 |
| Mean smear grade at baseline | Mean (SD) | 2842 | 0.99(1.36) | 1.00(1.39) | 0.96(1.32) | 0.503 |
| Prior TB | No. (%) | 1598 | 98(6.1%) | 75(6.84%) | 23(4.58%) | 0.080 |
| Cavity | No. (%) | 2894 | 413(14.27%) | 311(15.75%) | 102(11.09%) | <0.001 |

AFB = Acid fast bacilli. BMI= body mass index. IQR= interquartile range. CCI= Charlson Comorbidity Index. COPD= Chronic obstructive pulmonary disease. CVD= Cardiovascular diseases. HIV= Human Immunodeficiency Virus. TB=Tuberculosis.

During the first 9 months after ATT initiation, the log-rank test of the Kaplan-Meier analysis showed that males had significantly shorter survival compared to females (Figure 1A; p< 0.001). After adjusting for confounders such as BMI, CCI, COPD, alcoholism, smoking, cavitary disease (Table 2), males had a HR of 1.43(95%CI 1.08-1.98) for all-cause mortality. Infection-related causes accounted for 55.7% (303/544 patients) of all deaths in the first 9 months (Table 2). In the Cox regression analysis, males had an adjusted HR of 1.70 (95%CI 1.09-2.64) for infection-related mortality (Figure 1B).

**Table 2:** Association of male sex with tuberculosis treatment outcomes in the retrospective cohort using Cox and Logistic regression models (N=2894)

| Characteristic | Total | Men | Women | Estimate | Unadjusted Effect size | (95% CI) | p-value | Adjusted Effect size<sup>#</sup> | 95% CI | p-value |
| --- | --- | --- | --- | --- | --- | --- | --- | --- | --- | --- |
| **All-cause mortality (n=2667)** | 544(20.4%) | 427(23.2%) | 117(14.1%) | HR | 1.75 | 1.42-2.14 | <0.001 | 1.43 | 1.03-1.98 | 0.032 |
| **Infection related mortality (n=2667)** | 303(11.4%) | 231(12.6%) | 72(8.7%) | HR | 1.52 | 1.17-1.99 | 0.002 | 1.70 | 1.09-2.64 | 0.009 |
| **2-month Sputum culture positivity (n=1640)** | 265(16.2%) | 203(18.4%) | 62(11.6%) | OR | 1.72 | 1.27-2.34 | 0.003 | 1.56 | 1.05-2.33 | 0.028 |
| **2-month Sputum smear AFB (n=1640)** | 118(7.2%) | 91(8.3%) | 27(5.0%) | OR | 1.70 | 1.09-2.63 | 0.015 | 1.27 | 0.71-2.27 | 0.42 |

Abbreviations: AFB=Acid Fast bacilli. HR=Hazard Ratio. OR=Odds Ratio.

**Figure 1:**
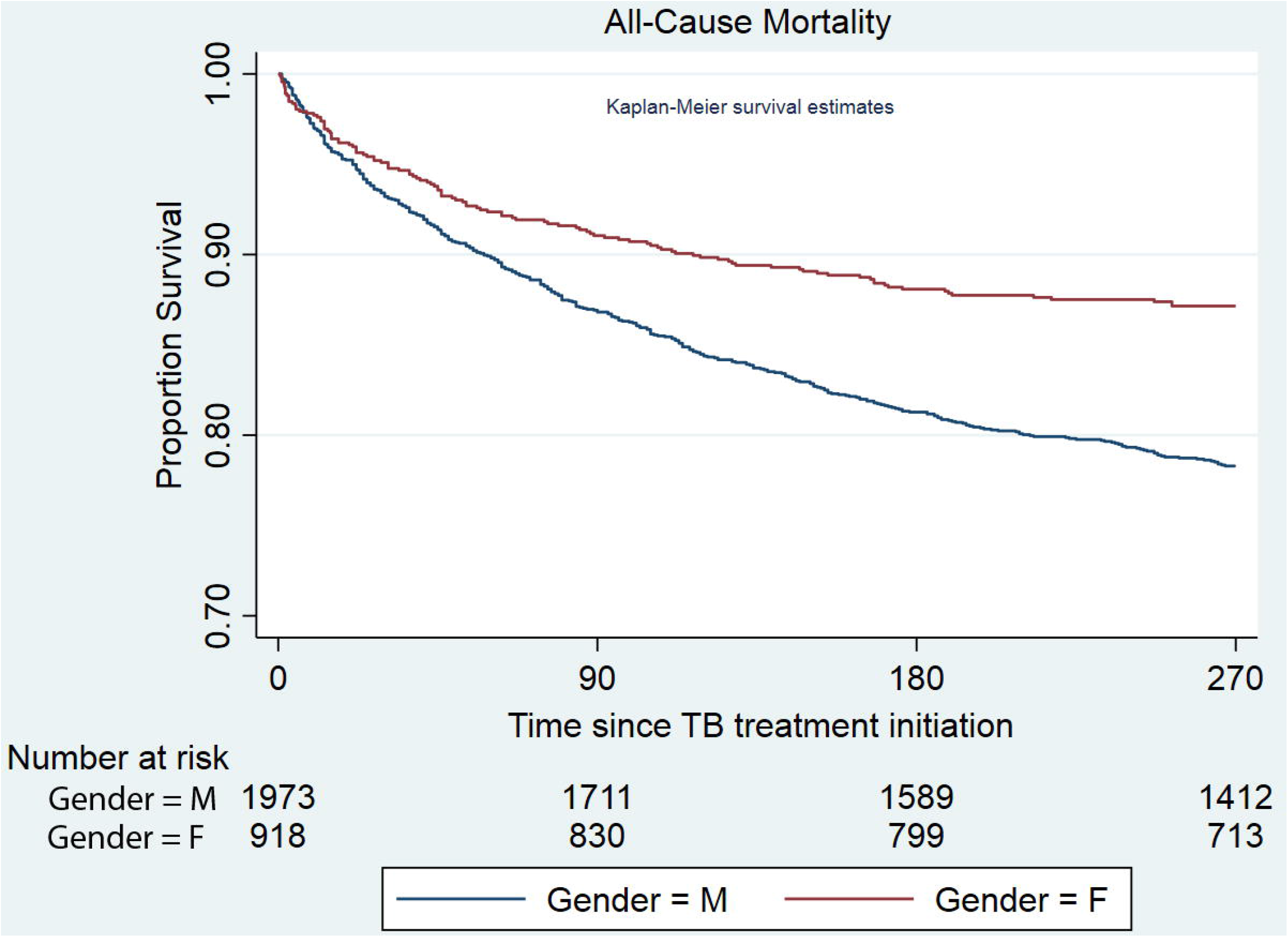
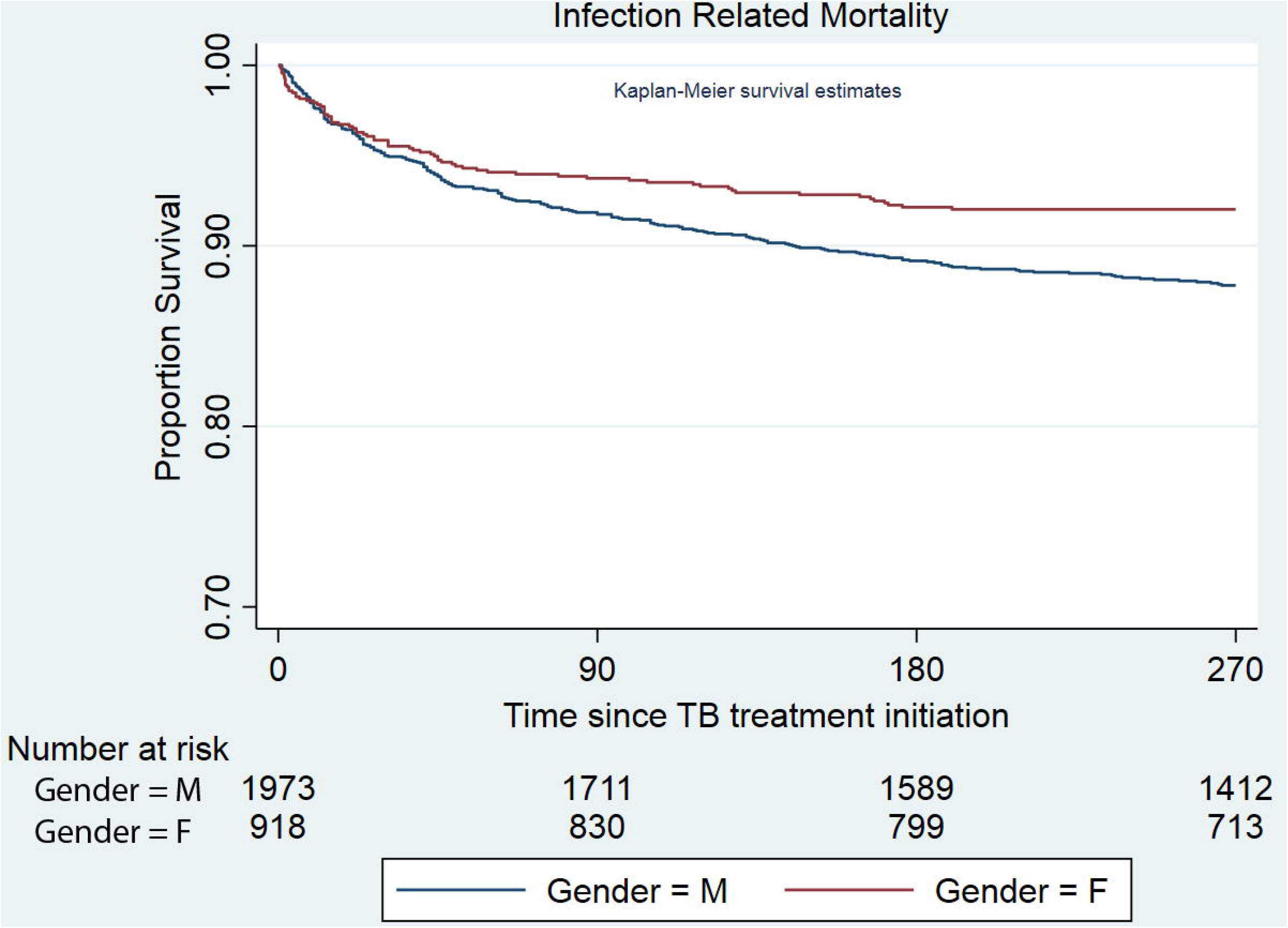
Kaplan-Meier Survival Graphs for 1A) All-cause mortality and 1B) Infection related mortality among patients treated for tuberculosis in the Retrospective cohort. M=Male. F=Female.

Males had significantly higher sputum culture (18.4% vs 11.6%, p=0.003) and AFB smear (8.3% vs 5.0%, p=0.015) positivity at 2 months compared to females by chi^2^ test, with a considerable difference primarily among individuals less than 50 years (Figure 2A and 2B). Multivariable logistic regression revealed higher odds of sputum-culture positivity in males (OR 1.56, 95%CI 1.05-2.33, p=0.03), but the odds of sputum AFB smear positivity rate (OR 1.27, 95%CI 0.71-2.27, p=0.42) was not different between the sexes at 2 months. Sensitivity analyses by additionally adjusting for age in the multivariable regression analyses also yielded similar results (Supplementary section III, Supplementary table 1).

**Figure 2:**
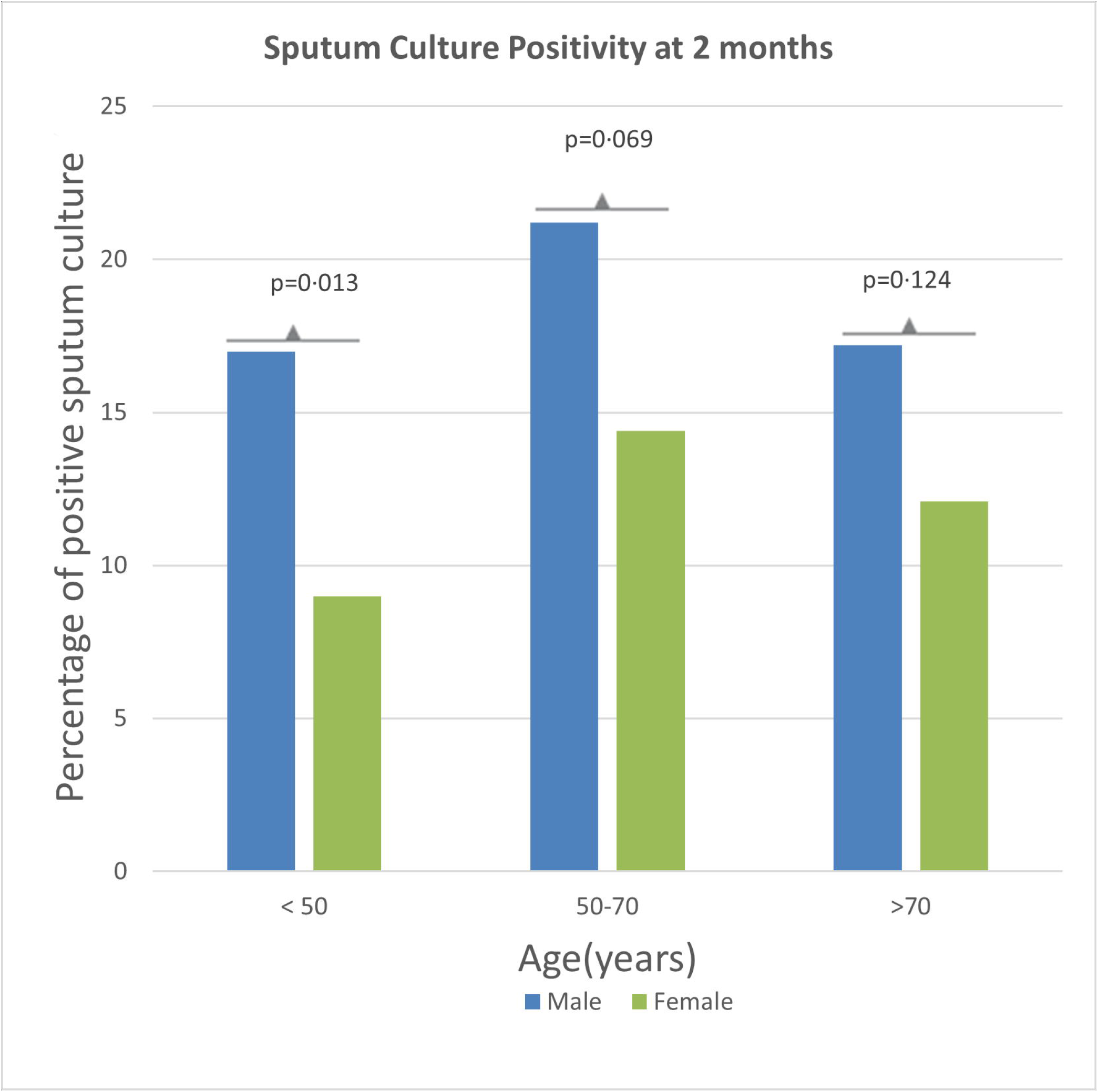
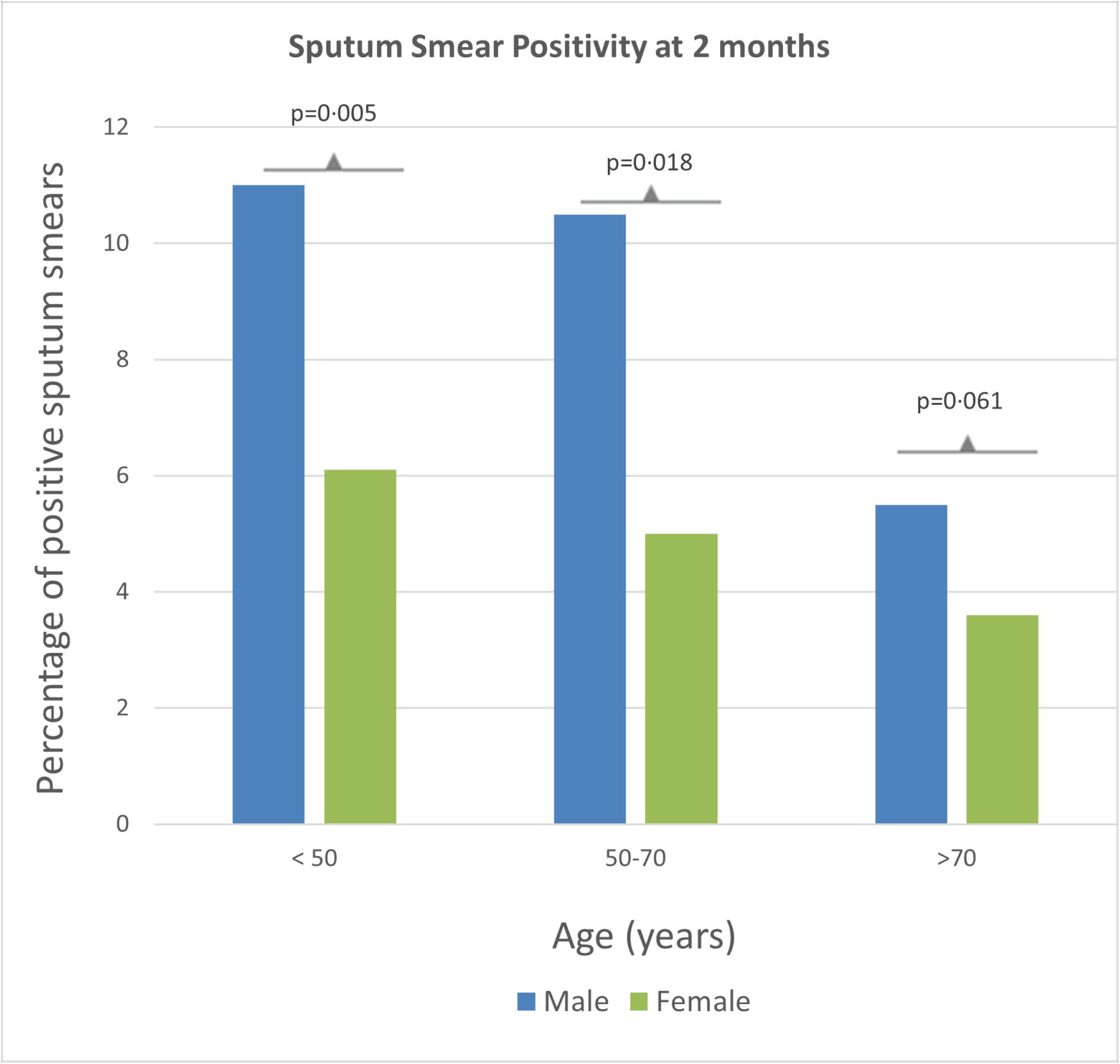
Microbiological outcomes after two months of tuberculosis treatment in the retrospective cohort. A. Sputum culture positivity. B. Sputum AFB smear positivity at 2 months since treatment initiation

### Systematic review

Among the 7896 studies screened, we selected 740 studies for full text review (Figure 3). Finally, data from 398 studies, reporting on a total of 3,957,216 individuals, were analyzed in our review (Supplementary table 2). These reports originated from 81 countries, with 56 studies from low-income, 235 studies from middle-income, and 94 studies from high-income countries, and 13 studies from multiple countries(21). We included 230 studies and 228 studies from countries with high-TB and high TB-HIV burdens, respectively(22). In our review, a total of 136 studies included only pulmonary TB (PTB), 20 studies included only extra-pulmonary TB (EPTB), and 242 studies included both PTB and EPTB patients. Among the included studies, 97 reported outcomes in patients with drug-susceptible TB, and another 100 reported outcomes in drug-resistant TB (60 studies included only MDR-TB patients, 5 studies reported only on XDR TB). The duration of ATT varied from 6 months to 24 months, depending on drug resistance profiles and site of TB involvement. HIV prevalence in the studies ranged from 0% to 100%. Forty-six studies included only patients with HIV-TB co-infection.

**Figure 3:**
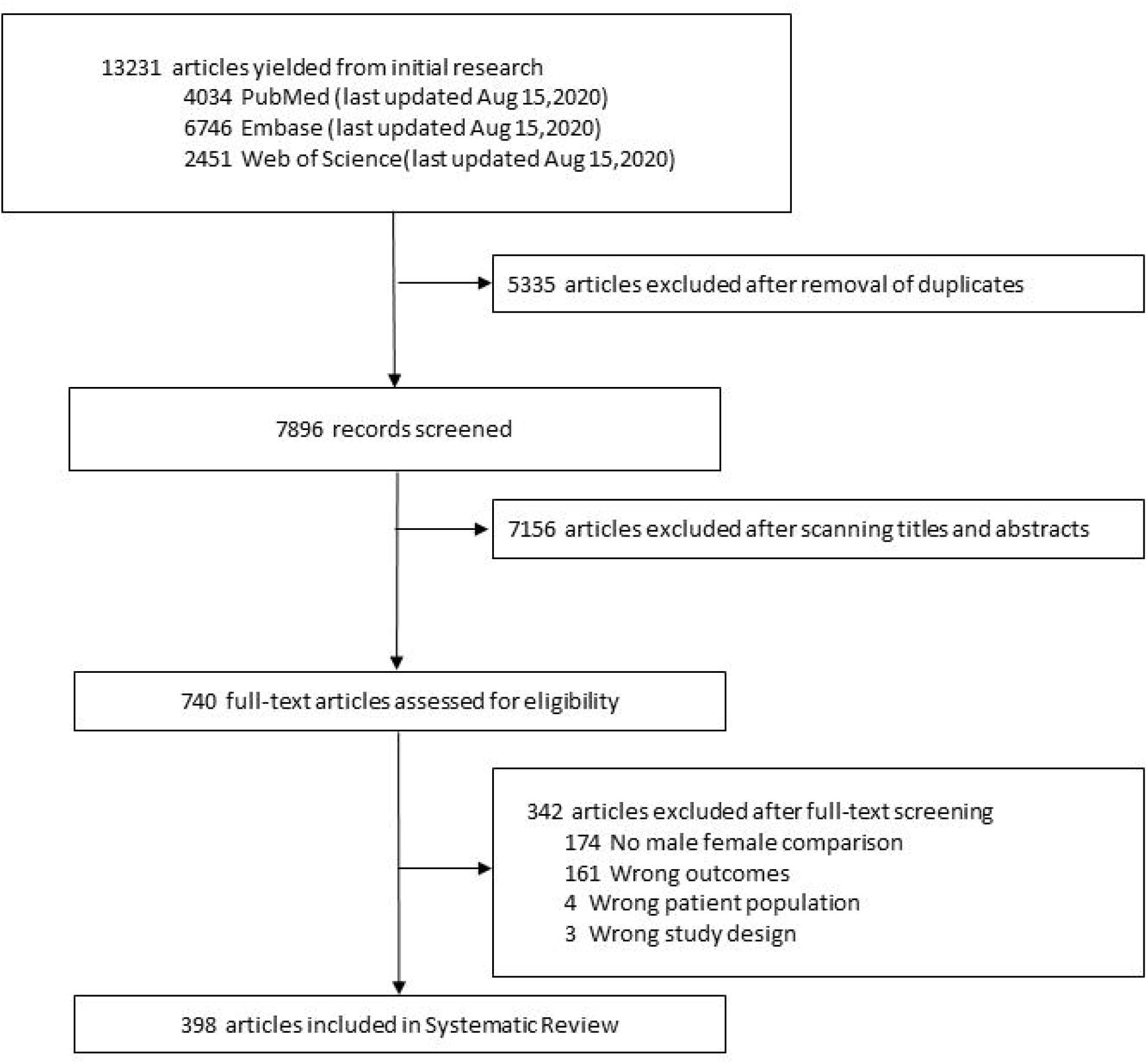
Study Selection for the systematic review

Among the included studies, 197 studies reported all-cause mortality, 12 reported death due to TB, 23 reported sputum culture positivity, 34 reported sputum AFB smear positivity after ATT initiation, and 130 reported TB treatment success (Supplementary table 2). In our review, there were 7 case-control, 75 prospective cohort, and 316 retrospective cohort studies. The cohorts were of varying duration, extending from 1981 to 2019. Among the included studies, 242 studies had data collected from hospital records, while 156 studies had programmatic data (from registries or administrative data) obtained from a centralized data source. There were 334 studies from countries that have implemented the DOTS (Directly Observed Treatment, Short-course) program and 51 from countries without the DOTS program.

Quality assessment using the New-Castle Ottawa scale (NOS) revealed that two studies (0.5%) scored the maximum of 9 points, 190 studies (47.7%) scored 8, 182 studies (45.7%) scored 7, and 11 studies (2.7%) scored 6. A total of 13 studies were identified as low-quality studies (NOS ≤ 5) (Supplementary table 2).

Males had higher unadjusted pooled effect sizes for mortality compared to females with a pooled OR of 1.26 (95%CI 1.19-1.34) from 135 studies (I^2^=77.9%), and a pooled HR of 1.17 (95%CI 1.07-1.27) from 44 studies (I^2^=64.5%) (Table 3). Eighty-two of the 197 studies reporting all-cause mortality made adjustments for at least one confounding variable. Commonly adjusted confounding parameters included age, HIV co-infection, site of TB, and BMI (Supplementary Table 3). The pooled adjusted OR was 1.26 (95%CI 1.16-1.37) among 52 studies (I^2^=90.9%) and the pooled adjusted HR was 1.20 (95%CI 1.05-1.37) among 30 studies (I^2^=82.5%) (Table 3).

**Table 3:** Association of male sex with all-cause mortality when compared to females among patients with tuberculosis using random effects meta-analysis

| Characteritics |  |  | All-cause mortality |  |  |  |  |
| --- | --- | --- | --- | --- | --- | --- | --- |
|  |  | No. of studies | OR [95%CI] | I<sup>2</sup> | No. of studies | HR [95%CI] | I<sup>2</sup> |
|  | Unadjusted |  |  |  |  |  |  |
| All studies |  | 135 | 1.26[1.19-1.34] | 77.90 | 44 | 1.17[1.07-1.27] | 63.59 |
| During treatment |  | 122 | 1.26[1.19-1.34] | 71.30 | 26 | 1.16[1.06-1.27] | 59.72 |
| General setting |  | 111 | 1.26[1.19-1.35] | 77.39 | 25 | 1.18[1.05-1.30] | 66.87 |
| ICU setting |  | 11 | 0.92[0.71-1.18] | 0 | 1 | 2.57[0.73-9.08] | .. |
| During follow up |  | 13 | 1.39[1.09-1.77] | 78.32 | 18 | 1.14[1.00-1.30] | 60.27 |
|  | Adjusted |  |  |  |  |  |  |
| All studies |  | 52 | 1.31[1.18-1.45] | 74.80 | 30 | 1.19[1.05-1.67] | 82.5 |

Abbreviations: HR=Hazard Ratio. OR= Odds Ratio; ICU= Intensive care unit.

Considerable heterogeneity was present in the meta-analyses of OR and HR for all-cause mortality. Studies reporting mortality of TB patients in the intensive care unit showed no significant difference in mortality when stratified by sex (Table 3). Further subgroup analyses were performed after excluding studies reporting outcomes in patients receiving intensive care (Table 4). Meta-regression analysis showed that the timepoint used for assessing mortality did not change the association between male sex and this outcome (Figure 4). Detailed results of the subgroup and meta-regression analysis on all-cause mortality are described in the supplementary document (Section IIb).

**Table 4:**
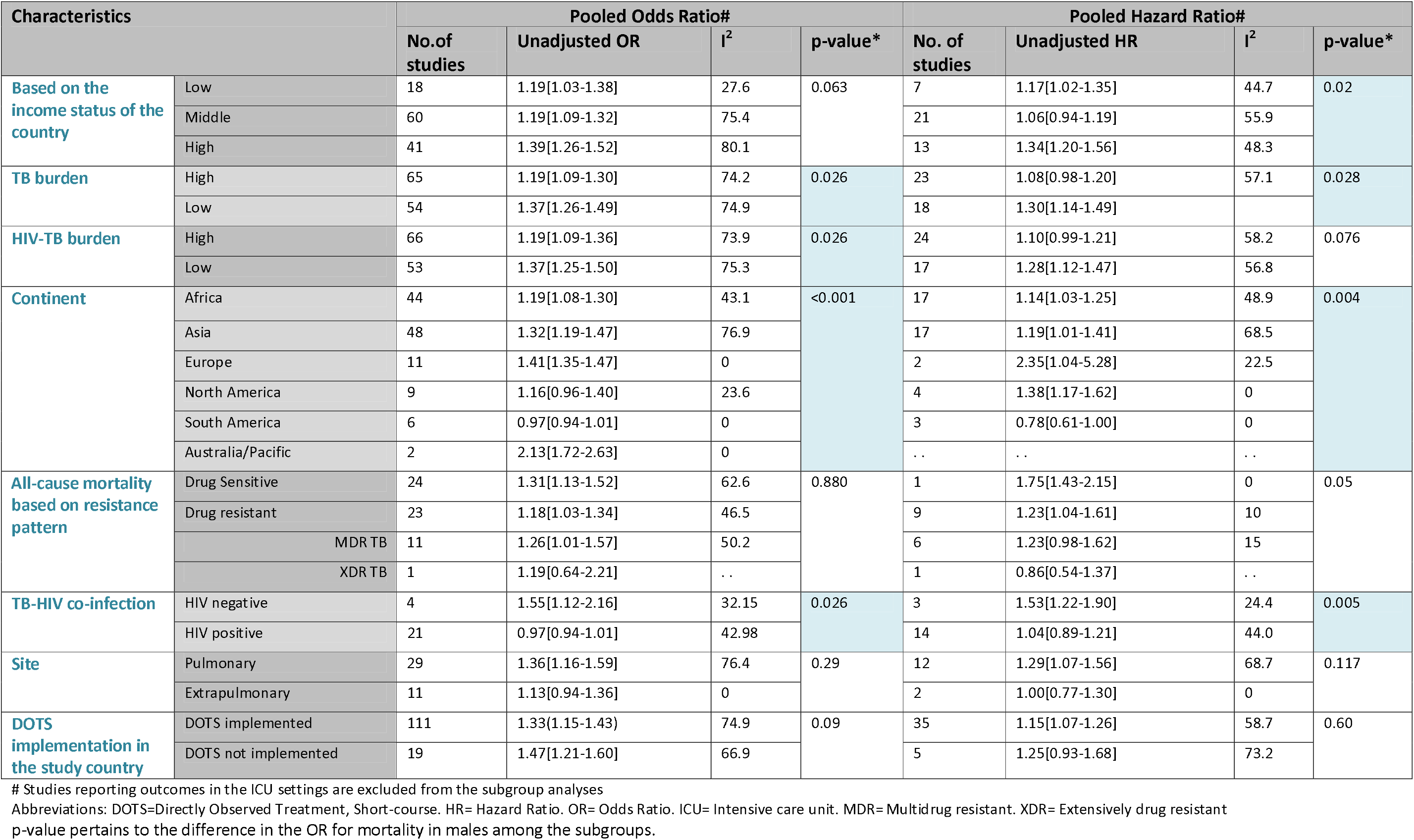
Subgroup analysis of the pooled effect sizes for the association of male sex with all-cause mortality when compared to females among patients with tuberculosis.

**Figure 4:**
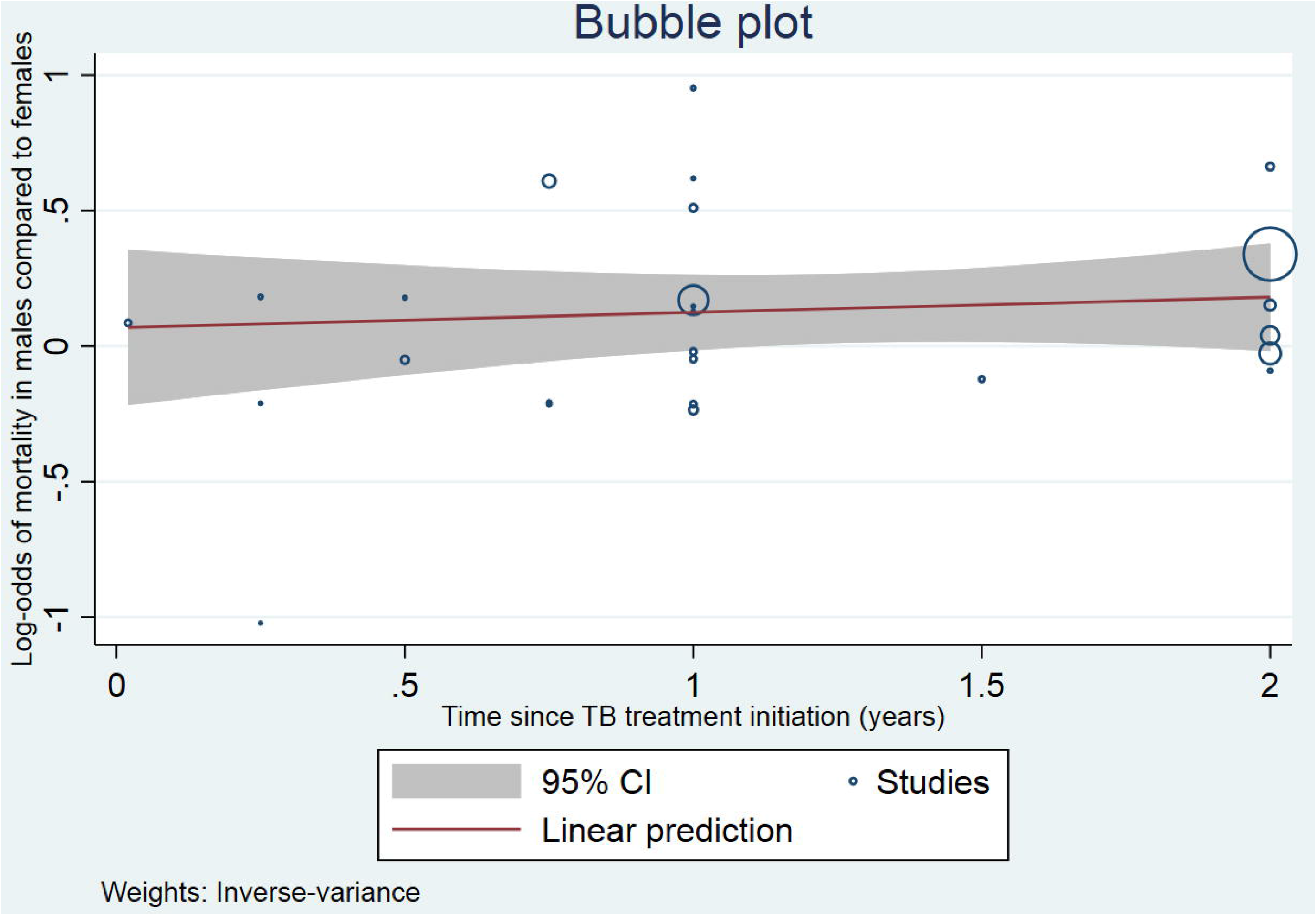
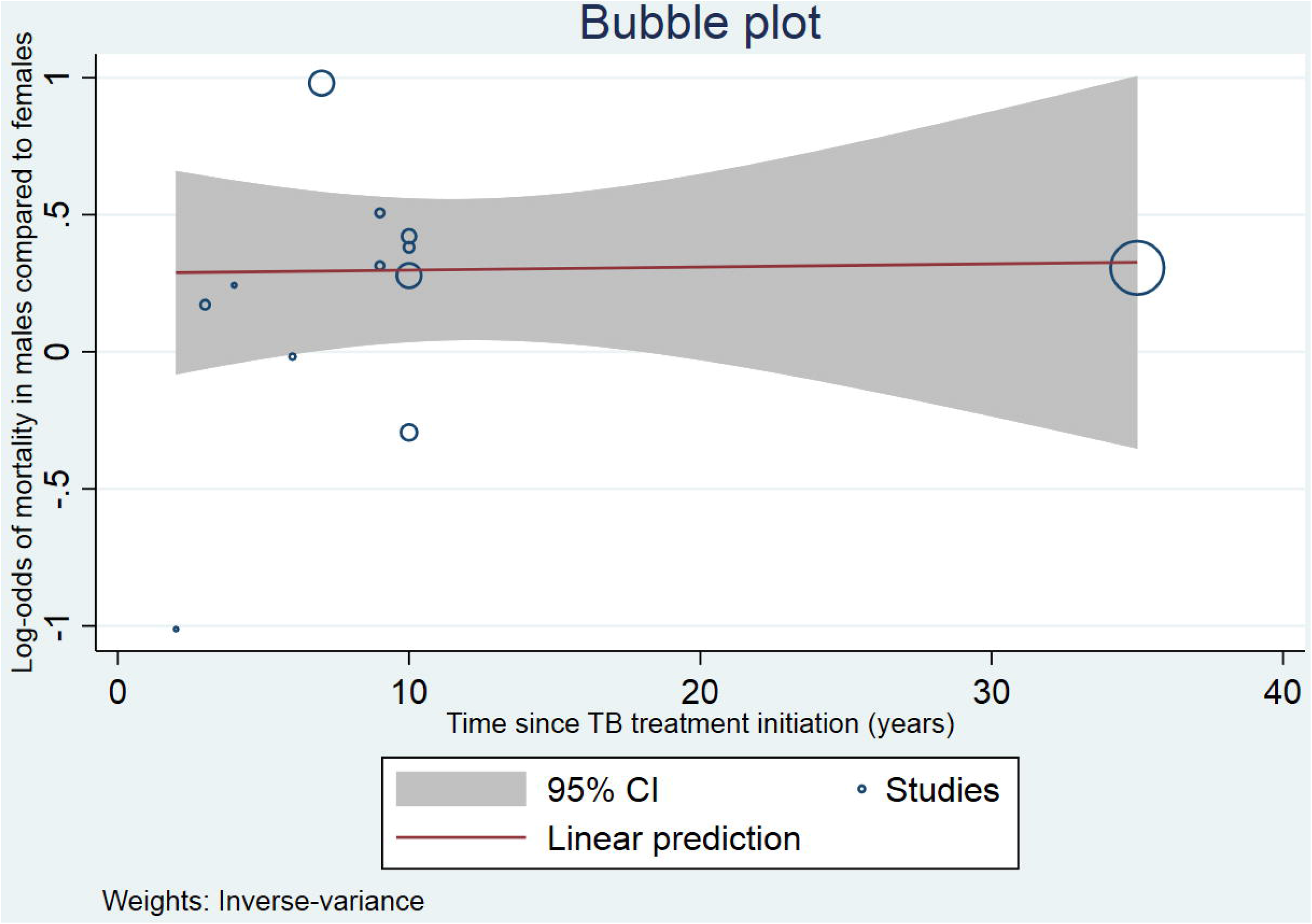
Bubble plot showing the association between log-odds of mortality in males compared to females and the time of assessment of mortality A) During TB treatment and B) During follow up.

We included 12 studies that reported death due to TB (Table 5). The pooled OR for death due to TB for males compared to females was 1.28 (95%CI 1.1-1.6) in 10 studies and the pooled HR was 1.34 (95%CI 1.09-1.55) in 2 studies. The pooled OR for death due to non-TB causes for males compared to females with TB was 1.3(95%CI 1.1-1.7) in 6 studies.

**Table 5:** Studies reporting the association of male sex with death due to TB and non-TB causes when compared to females in patients with tuberculosis

| Author (Year) | Country | Type | Data | HIV (%) | Site | Males | Females | OR/HR | Death due to TB | Death due to non-TB causes | All-cause mortality | Timeline |
| --- | --- | --- | --- | --- | --- | --- | --- | --- | --- | --- | --- | --- |
| Macedo 2013 | Brazil | Retrospective cohort | Centralized data source | 13.5 | Both PTB and EPTB | 13489 | 1385 | OR | 0.8[0.6-1.1] | 1.3[0.9-1.8] | .. | During treatment |
| Prado 2017 | Brazil | Retrospective cohort | Centralized data source | 100 | Both PTB and EPTB | 48348 | 19934 | OR | 0.9[0.8-1.0] | 0.9[0.8-1.0] | 0.9[0.8-1.0] | During treatment |
| Wu 2015 | Taiwan | Retrospective cohort | Centralized data source | .. | Both PTB and EPTB | 4165 | 1419 | OR | 1.1[0.9-1.3] | 1.2[1.1-1.3] | .. | During treatment |
| Shuldiner 2014 | Israel | Retrospective cohort | Centralized data source | 6.1 | Both PTB and EPTB | 2612 | 1925 | OR | 1.3[1.0-1.6] | .. | .. | During treatment |
| Yen 2013 | Taiwan | Retrospective cohort | Centralized data source | .. | PTB | 2455 | 1032 | OR | 1.7[1.1-2.4] | 1.8[1.5-2.3] | .. | During treatment |
| Dale 2016 | Australia | Retrospective cohort | Centralized data source | 1.1 | Both PTB and EPTB | 2125 | 1831 | OR | 1.8[1.1-2.9] | 2.1[1.2-3.6] | 1.9[1.3-2.7] | During treatment |
| Baez 2016 | Mexico | Prospective cohort | Centralized data source | 2.4 | PTB | 462 | 342 | HR | 1.6[0.5-5.5] | .. | 1.6[1.1-2.4] | During treatment |
| Yilmaz 2015 | Turkey | Retrospective cohort | Centralized data source | .. | Both PTB and EPTB | 1339 | 1111 | OR | 1.5[0.8-2.7] | .. | .. | 9 years FU |
| Wang 2013 | China | Retrospective cohort | Centralized data source | .. | PTB | 3173 | 1098 | OR | 2.9[1.7-5.6] | .. | 2.6[2.1-3.3] | 7 years FU |
| Golub 2019 | South Korea | Retrospective cohort | Centralized data source | .. | PTB | 819051 | 448513 | OR | 1.7[1.4-2.0] | .. | .. | 12 years FU |
| Wen 2010 | Taiwan | Prospective cohort | Centralized data source | .. | PTB | 3118 | 1918 | HR | 5.7[3.1-10.5] | .. | .. | 10 years FU |
| Macedo 2013 | Brazil | Retrospective cohort | Centralized data source | 13.5 | Both PTB and EPTB | 13489 | 1385 | OR | 0.8[0.6-1.1] | 1.3[0.9-1.8] | .. | During treatment |
| Pooled Estimates |  |  |  |  |  |  |  |  | 1.28[1.1-1.6] | 1.3[1.1-1.7] | 1.6[1.1-2.5] | .. |

Abbreviations: EPTB= Extra-Pulmonary tuberculosis. HR= Hazard Ratio. OR= Odds Ratio. PTB=Pulmonary tuberculosis. p-value pertains to the difference in the OR for mortality in males among the subgroups.

Males had a pooled unadjusted OR of 1.60 (95%CI 1.28-1.99) in 12 studies (I^2^=24.6%) and pooled adjusted OR of 1.81 (95%CI 1.39-2.34) in 4 studies (I^2^=0%) (Table 6) compared to females at 2 or 3 months of ATT compared to females. Among 7 studies that reported HR for sputum-culture conversion to negativity, males had a pooled unadjusted HR of 0.83 (95%CI 0.70-0.97) and a pooled adjusted HR of 0.76(95%CI 0.59-0.97) compared to females (Supplementary section IV). With respect to sputum AFB smear positivity, males had a pooled unadjusted OR of 1.42 (95%CI 1.26-1.60) in 29 studies (I^2^=17.1%) and pooled adjusted OR of 1.45 (1.31-1.61) in 6 studies (I^2^=0%) compared to females (Table 6). Our meta-analysis revealed that odds of treatment success was lower in males in the meta-analysis of both adjusted and unadjusted effect sizes (Supplementary Table 5). There was very low heterogeneity in the data related to sputum culture or AFB smear positivity. Thus, these outcomes did not warrant further subgroup analysis.

**Table 6:** Association of male sex with Sputum smear and sputum culture positivity when compared to females among patients with tuberculosis using random effects meta-analysis.

| Outcome | Time of assessment | Estimate | Unadjusted |  |  |  | Adjusted |  |  |  |
| --- | --- | --- | --- | --- | --- | --- | --- | --- | --- | --- |
|  |  |  | No. of studies | Effect size | I<sup>2</sup> | p-value | No. of studies | Effect size | I<sup>2</sup> | p-value |
| **Sputum culture for AFB** | All studies | OR | 13 | 1.60[1.28-1.99] | 24.6 | .. | 4 | 1.81[1.39-2.34] | 0 | .. |
|  | End of intensive phase | OR | 13 | 1.60[1.28-1.99] | 24.6 |  | 4 | 1.81[1.39-2.34] | 0 |  |
|  | End of Treatment | OR | .. | .. | .. |  | .. | .. | .. |  |
| **Sputum Smear for AFB** | All studies | OR | 29 | 1.40[1.24-1.58] | 21.06 | 0.06 | 6 | 1.45[1.31-1.61] | 0 | 0.48 |
|  | End of intensive phase | OR | 6 | 1.58[1.41-1.77] | 0 |  | 3 | 1.42[1.26-1.60] | 0 |  |
|  | End of Treatment | OR | 22 | 1.23[1.02-1.48] | 28.27 |  | 3 | 1.51[1.16-1.98] | 26.06 |  |

Abbreviations: EPTB= Extra-Pulmonary tuberculosis. OR= Odds Ratio. PTB=Pulmonary tuberculosis.
p-value pertains to the difference in the OR for males between the subgroups (End of intensive phase and end of treatment).

In the meta-regression analysis, mean age of the population and the proportions of patients with diabetes mellitus, hypertension, alcohol abuse, smoking, cardiovascular events, COPD and HIV did not significantly modify the association between male sex and any of the outcomes assessed (Supplementary Table 6). Publication bias measured by Egger’s test was not present for any of the outcomes (Supplementary Section IV).

## Discussion

Findings from our retrospective cohort and systematic review, comprising a total of 398 peer-reviewed publications from over 80 high-, middle-, and low-income countries, suggest that male sex is associated with higher all-cause mortality and death due to TB, both during treatment and during follow up after ATT. This association held true even after adjusted for confounders in the cohort study and during extensive meta-regression analysis in the systematic review. With respect to microbiological outcomes, males had higher positivity for *Mtb* by both AFB smear microscopy and culture, at the end of the intensive phase as well as at ATT completion. In addition, male sex was found to be associated with lower treatment success rates.

Our cohort study only included patients with drug-susceptible pulmonary TB from Taiwan, which is a high-income country with excellent access to health care resources. Our systematic review helped to expand and corroborate our findings with countries with variable health care resources. The meta-regression analysis demonstrated that mortality in males compared to females was significantly higher in high-income countries than in low- or middle-income countries (Table 4). This suggests that sex differences in TB treatment outcomes are more apparent in high-income settings with minimal differences in healthcare access between the sexes. It has been widely believed that socioeconomic and cultural factors are responsible for the observed sex bias. Patient-related delays and health care system delays in the diagnosis of TB and initiation of ATT have been shown to be more pronounced in women(24–26) due to financial dependence on the family, fear of social isolation and accessing less qualified providers were considered as major reasons for these treatment delays(27–29).

In a cohort of TB patients from Australia, males had significantly higher rates of hemoptysis, and loss of body weight. and were more likely to have a sputum smear and chest imaging performed, with earlier treatment initiation(30). Males had higher rates of abnormal chest X-ray and pleural effusion, and higher odds of TB-related and all-cause mortality in spite of comparable treatment completion rates between the sexes(30). These observations are consistent with the hypothesis that males exhibit more severe TB disease at presentation and have worse outcomes despite better healthcare access, and prompt treatment initiation.

Similarly, in a cohort of non-smoking TB patients without comorbidities, Tan *et al*. demonstrated that despite the delay in treatment initiation in women, high-resolution computed tomography (CT) showed more severe lung damage in men at baseline(7). Additionally, scoring of the CT images pre- and post-ATT showed a more rapid response to therapy in women than in men(7). Likewise, in a study by Chu *et al*. of 594 non-smoking men and women with newly diagnosed TB, multivariate logistic regression analysis revealed that males had 2.7 times higher odds of cavitary disease, and lung injury scores(6).

In our cohort, men had disproportionately high rates of smoking and cavitary disease compared to women. Matsumoto *et al*. found that male current smokers had higher rates of cavitary disease compared to female smokers(31). This could be explained by increased expression of matrix metalloproteinase in the lungs in men(32). Furthermore, the presence of lung cavitation is known to be associated with delayed sputum conversion during ATT(33). In our cohort, male sex was associated with a higher odds of sputum culture positivity at 2 months of ATT, after adjusting for confounding factors, such as smoking and cavitary disease. Likewise, our systematic review showed higher pooled unadjusted and adjusted odds of sputum AFB smear and culture positivity in males following treatment initiation. These findings emphasize that male sex is a risk factor for unfavorable microbiological outcomes during ATT.

The phenomenon that male sex predisposes to infectious diseases has been recognized in liver abscess due to *Entamoeba histolytica*(34, 35), respiratory and systemic infection due to *Streptococcus pneumoniae*(36), and higher mortality due to SARS(37) and COVID-19(38, 39). Although such differences have been noted in TB outcomes, these have been largely attributed to social, economic, and cultural determinants rather than biological ones. More recently, studies have demonstrated how sex hormones can differentially regulate female and male immune responses to pathogens(40). Yamamoto and colleagues reported that peritoneal macrophages derived from male mice exhibited reduced antimicrobial activity and promoted mycobacterial growth(41, 42). Testosterone was shown to reduce the expression of TLR4 (toll-like receptor 4) in mouse macrophages(43) and to have immunosuppressive effects by increasing inhibitory cytokines, reducing Ig production, and inhibiting T and B cell maturation(44–47). In contrast, estradiol has been shown to enhance macrophage activation(48); and in turn have a greater clearance of pneumococcal pneumonia in female mice(49). Various studies have shown more severe disease in male mice following nontuberculous mycobacteria and *Mtb* infections, manifested by higher lung bacillary burdens, increased mortality and significantly increased levels of pro-inflammatory cytokines(10, 11), consistent with an excessive inflammatory response in males relative to females(50).

In the systematic review, males had a higher pooled OR of mortality in studies with exclusively HIV-negative TB patients when compared to that in studies on HIV-TB co-infected patients. Prior literature from Africa has shown that males had worse outcomes compared to females in HIV-negative patients, but better outcomes compared to females among HIV-co-infected patients(51, 52). Thus, HIV co-infection appears to reduce the potential protective effect of female sex on TB treatment outcomes. This conclusion aligns well with the evidence in pre-clinical models that immunomodulation is responsible for the difference in outcomes between males and females.

Although men have a higher incidence of TB, women have an increased incidence of EPTB compared to men(53–55). In our systematic review, the rates of mortality in men compared to women did not significantly differ between the studies on PTB and studies on EPTB. In studies with varying proportions of PTB and EPTB, the proportion of EPTB in the study population did not modify the association between mortality and sex. Likewise, the drug sensitivity of the study participants to ATT did not have any impact on the higher mortality rates in males.

Our study has several strengths. The large sample size of our cohort study aided in the analysis of various confounding factors, such as comorbidities and social habits. As the cohort data were obtained from a single institution, heterogeneity in treatment decisions were minimized. Our systematic review included 398 studies from 81 countries with variable health care access, allowing for generalizability of our results. The large number of studies on all-cause mortality in our systematic review enabled us to perform subgroup analyses and to arrive at various inferences. Our systematic review included several studies on MDR TB and XDR TB, enabling our conclusions to apply to those with drug-resistant TB.

Several limitations are noted. In our cohort study, we were unable to assess parameters such as occupational history, symptoms at diagnosis, and delay in diagnosis. In our systematic review, the drug sensitivity characteristics, study setting, and time of outcome assessment varied widely and were not reported consistently among the included studies. Several studies in our review, performed from program registries, did not report data on baseline characteristics stratified by sex. During meta-analysis of adjusted effect sizes, individual studies were adjusted for comparable but different parameters. As the cohorts spanned over long periods of time, the treatment regimens followed may not be uniform across studies.

Our results have several important therapeutic implications. Recognizing the association between male sex and adverse treatment outcomes should inform medical decision-making in TB treatment programs and motivate further focused research into the biological (immunological and genetic) basis of these sex-based differences and in turn novel host-directed therapies to improve clinical, pathological, and microbiological outcomes during TB treatment in both sexes.

## Data Availability

The data is available on request to the corresponding author.

## Contributors

VC and PK conceived the idea for the review. VC, NT, MM, JB and PK designed and undertook the literature review. VC, NT, MM, SA, AK, PN, EW and EA screened the articles and extracted the data with help from AZ and SW. VC, MM and NT performed the statistical analysis, figures, and appendix. VC, MM, AK and RK analyzed and interpreted the data. VC, JRC, MM, AK, RK, and PK wrote the first draft of the manuscript. VC, AG, JW and PK revised the subsequent drafts of the manuscript. All authors reviewed the final draft of the manuscript.

## Data sharing

All the forest plots and figures are included in the supplementary document of the manuscript. Cohort datasets will be available on further communication to the corresponding author.

## Acknowledgments

The cohort study was supported by the National Institute of Allergy and Infectious Diseases (NIAID)/ National Institutes of Health (NIH) grants UH3AI122309 and K24AI143447 to P.C.K.; and the Taiwan Centers for Disease Control grant MOHW-105-CDC-C-114-000103 to J.-Y.W.

