## Supplementary appendix for "Sex differences in TB treatment outcomes: Retrospective cohort study and meta-analysis"

| Section | Content | Page |
|---|---|---|
| **I** | Search Strategy | 3 |
| **IIa** | Supplementary Methods | 5 |
| **IIb** | Supplementary Results | 7 |
| **III** | Supplementary tables | 8 |
| **IV** | Supplementary figures (Forest charts and funnel plots) | 33 |
| **V** | References | 64 |

### Section I

#### Search Strategy

(Studies restricted to the last 10 years)

**PubMed**

(("Tuberculosis"[Mesh]  OR "Mycobacterium tuberculosis"[Mesh] OR "tuberculosis"[tiab] OR "TB"[tiab] OR "tuberculous"[tiab] OR "Tuberculosis, Multidrug-Resistant"[Mesh] OR "multi drug resistant tuberculosis"[tiab] OR "MDR TB"[tiab] OR "Extensively Drug-Resistant Tuberculosis"[Mesh] OR "extensively drug resistant tuberculosis"[tiab] OR "XDR TB"[tiab] OR "latent tuberculosis infection"[tiab] OR "LTBI"[tiab] OR “tuberculoses”[tiab]) AND ("Sex Characteristics"[Mesh] OR "Sex Factors"[Mesh] OR "Male"[Mesh] OR "Female"[Mesh] OR "Male"[tiab] OR "Female"[tiab] OR "males"[tiab] OR "females" OR "Sex"[tiab] OR "sexes"[tiab] OR "gender"[tiab] OR "genders"[tiab]) AND ("Disease-Free Survival"[Mesh] OR "Treatment Failure"[Mesh] OR "Fatal Outcome"[Mesh] OR "Survival Rate"[Mesh] OR "Mortality"[Mesh] OR "Recurrence"[Mesh] OR "relapse"[tiab] OR "relapsed"[tiab] OR "relapsing"[tiab] OR "survival"[tiab] OR "survived"[tiab] OR "death"[tiab] OR "deaths"[tiab] OR "mortality"[tiab] OR "Recurrence"[tiab] OR "recurrences"[tiab] OR (("Sputum"[Mesh] OR "sputum"[tiab] OR "sputums"[tiab]) AND (months*[tiab])) AND ("Clinical Study"[Publication Type:NoExp] OR "Clinical Trial"[Publication Type] OR "Observational Study"[Publication Type] OR "Controlled Clinical Trial"[Publication Type] OR "Retraction of Publication"[Publication Type] OR "Systematic Review"[Publication Type] OR "Epidemiologic Studies"[Mesh] OR "systematic review"[tiab] OR "meta-analysis"[tiab] OR "meta analysis"[tiab] OR "metaanalysis"[tiab] OR "systematic overview"[tiab] OR "scoping review"[tiab] OR "integrative review"[tiab] OR "quantitative review"[tiab] OR "quantitative overview"[tiab] OR "cohort"[tiab] OR "case control"[tiab] OR "case controlled"[tiab] OR "controlled trial"[tiab] OR "controlled trials"[tiab] OR "clinical trial"[tiab] OR "clinical trials"[tiab] OR "random"[tiab] OR "randomly"[tiab] OR "randomized"[tiab] OR "randomised"[tiab] OR "single blind"[tiab] OR "double blind"[tiab] OR "single blinding"[tiab] OR "double blinding"[tiab] OR "single blinded"[tiab] OR "double blinded"[tiab] OR (("retrospective"[tiab] OR "retrospectively"[tiab] OR "prospective"[tiab] OR "observational"[tiab] OR "longitudinal"[tiab] OR "longitudinally"[tiab] OR "follow up"[tiab]) AND ("study"[tiab] OR "studies"[tiab])) OR "clinical study"[tiab] OR "clinical studies"[tiab] OR "validation study"[tiab] OR ("study"[tiab] AND "participants"[tiab]))) NOT ("animals"[mh] NOT ("animals"[mh] AND "humans"[mh]))

**Embase**

('tuberculosis'/exp OR 'Mycobacterium tuberculosis'/exp OR ‘tuberculosis’:ti,ab OR ‘TB’:ti,ab OR ‘tuberculous’:ti,ab OR 'drug resistant tuberculosis'/exp OR 'extensively drug resistant tuberculosis'/exp OR 'multidrug resistant tuberculosis'/exp OR ‘multi drug resistant tuberculosis’:ti,ab OR ‘MDR TB’:ti,ab OR ‘extensively drug resistant tuberculosis’:ti,ab OR ‘XDR TB’:ti,ab OR ‘latent tuberculosis infection’:ti,ab OR ‘LTBI’:ti,ab OR ‘tuberculoses’:ti,ab) AND ('sex difference'/exp OR 'sexual characteristics'/exp

OR 'sex factor'/exp OR 'male'/exp OR 'female'/exp OR ‘Male’:ti,ab OR ‘Female’:ti,ab OR ‘males’:ti,ab OR ‘females’:ti,ab OR ‘Sex’:ti,ab OR ‘sexes’:ti,ab OR ‘gender’:ti,ab OR ‘genders’:ti,ab) AND ('disease free survival'/exp OR 'treatment failure'/exp OR 'fatality'/exp OR 'mortality rate'/exp OR 'survival rate'/exp OR 'mortality'/exp OR 'recurrent disease'/exp OR ‘relapse’:ti,ab OR ‘relapsed’:ti,ab OR ‘relapsing’:ti,ab OR ‘survival’:ti,ab OR ‘survived’:ti,ab OR ‘death’:ti,ab OR ‘deaths’:ti,ab OR ‘mortality’:ti,ab OR ‘Recurrence’:ti,ab OR ‘recurrences’:ti,ab OR (('sputum'/exp OR ‘sputum’:ti,ab OR ‘sputums’:ti,ab) AND (‘months*’:ti,ab))) AND ('clinical study'/de OR 'case control study'/exp OR 'prospective study'/exp OR 'retrospective study'/exp OR 'major clinical study'/exp OR 'cross-sectional study'/exp OR 'cohort analysis'/exp OR 'clinical trial'/exp OR 'observational study'/exp OR 'controlled clinical trial'/exp OR 'retraction notice'/exp OR 'systematic review'/exp OR ‘systematic review’:ti,ab OR ‘meta-analysis’:ti,ab OR ‘meta analysis’:ti,ab OR ‘metaanalysis’:ti,ab OR ‘systematic overview’:ti,ab OR ‘scoping review’:ti,ab OR ‘integrative review’:ti,ab OR ‘quantitative review’:ti,ab OR ‘quantitative overview’:ti,ab OR ‘cohort’:ti,ab OR ‘case control’:ti,ab OR ‘case controlled’:ti,ab OR ‘controlled trial’:ti,ab OR ‘controlled trials’:ti,ab OR ‘clinical trial’:ti,ab OR ‘clinical trials’:ti,ab OR ‘random’:ti,ab OR ‘randomly’:ti,ab OR ‘randomized’:ti,ab OR ‘randomised’:ti,ab OR ‘single blind’:ti,ab OR ‘double blind’:ti,ab OR ‘single blinding’:ti,ab OR ‘double blinding’:ti,ab OR ‘single blinded’:ti,ab OR ‘double blinded’:ti,ab OR ((‘retrospective’:ti,ab OR ‘retrospectively’:ti,ab OR ‘prospective’:ti,ab OR ‘observational’:ti,ab OR ‘longitudinal’:ti,ab OR ‘longitudinally’:ti,ab OR ‘follow up’:ti,ab) AND (‘study’:ti,ab OR ‘studies’:ti,ab)) OR ‘clinical study’:ti,ab OR ‘clinical studies’:ti,ab OR ‘validation study’:ti,ab OR (‘study’:ti,ab AND ‘participants’:ti,ab)) NOT ('animal'/exp NOT ('animal'/exp AND 'human'/exp))

**Web of Science**

TS=("tuberculosis" OR "TB" OR "tuberculous" OR "multi drug resistant tuberculosis" OR "MDR TB" OR "extensively drug resistant tuberculosis" OR "XDR TB" OR "latent tuberculosis infection" OR "LTBI" OR “tuberculoses”) AND TS=("Male" OR "Female" OR "males" OR "females" OR "Sex" OR "sexes" OR "gender" OR "genders") AND TS=(“Treatment Failure” OR "Fatal Outcome” OR "relapse" OR "relapsed" OR "relapsing" OR “survival" OR "survived" OR "death" OR "deaths" OR "mortality" OR "Recurrence" OR "recurrences" OR (("sputum" OR "sputums") AND (month*))) AND TS=((retract* NEAR/3 publication*) OR "epidemiologic studies" OR “epidemiologic study” OR "epidemiological studies" OR “epidemiological study” OR "systematic review" OR "meta-analysis" OR "meta analysis" OR "metaanalysis" OR "systematic overview" OR "scoping review" OR "integrative review" OR "quantitative review" OR "quantitative overview" OR "cohort" OR "case control" OR "case controlled" OR "controlled trial" OR "controlled trials" OR "clinical trial" OR "clinical trials" OR "random" OR "randomly" OR "randomized" OR "randomised" OR "single blind" OR "double blind" OR "single blinding" OR "double blinding" OR "single blinded" OR "double blinded" OR (("retrospective" OR "retrospectively" OR "prospective" OR "observational" OR "longitudinal" OR "longitudinally" OR "follow up") AND ("study" OR "studies")) OR "clinical study" OR "clinical studies" OR "validation study" OR ("study" AND "participants"))

### <u>Section IIa</u>

### Supplementary Methods

#### <u>Systematic Review</u>

##### Search strategy and study selection

The systematic review was conducted according to the PRISMA guidelines[399]. The literature searches were performed in PubMed, Embase, and Web of Science on August 15, 2020, using the search strategy detailed in the supplementary document (Section I), to capture eligible reports published in the last ten years. We included both research articles and letters in the English language. Only articles published in peer-reviewed academic journals were included; conference abstracts were not included.

Studies were required to report sex-disaggregated data on at least one of the following outcomes on adult tuberculosis patients treated with multidrug anti-TB therapy (ATT): all-cause mortality, mortality due to TB, sputum AFB smear or culture positivity during or at the end of TB treatment, or 'treatment success' according to the WHO definitions for reporting TB outcomes[400]. *Treatment success* is a ratio of *favorable outcome* comprising 'cure' or 'treatment completion' to *unfavorable outcome* comprising 'failure,' 'death,' 'default' or 'loss to follow up'[400]. We included prospective and retrospective cohort studies and case-control studies. We excluded case reports, case series, and cross-sectional studies. Efforts were made to avoid overlap of patients across studies by collecting information on the name of the hospital, study period, and the names of the investigators. We utilized the COVIDENCE platform for the systematic review[401]. After removing the duplicates, the titles and abstracts of the retrieved articles were screened by at least two authors (VC, NT, AK, or PN) independently and the disagreements were resolved by VC. At least two authors (VC, NT, AK, or MM) independently performed the full-text screening, and the conflicts were resolved by VC.

##### Data extraction and quality assessment

At least two authors extracted data from the articles (VC, NT, MM, RK, SA, EW, EA, SW, or AZ) in the Qualtrics platform[402], and discrepancies were resolved by VC. We used the data extraction form developed using the Qualtrics platform[402]. Data on the country of study, funding source, patient comorbidities, site of TB involvement, pattern of resistance to ATT, HIV-TB co-infection, duration of treatment, and time points for outcomes. Data on treatment outcomes were extracted either as raw data or as pre-calculated effect sizes, namely odds ratio (OR), relative risk (RR), or hazard ratio (HR), along with 95% confidence interval (CI), as reported in the studies. RR for mortality in males compared to females was reported only in 7 studies, and were therefore converted to OR to facilitate pooling of the effect sizes[403]. We performed quality assessment using the New-Castle Ottawa scale for observational studies (NOS)[404].

**Data analysis**

For each of the outcomes, we performed a meta-analysis of the OR using the random-effects model. We pooled HR separately for each of the outcomes. We pooled the estimates for the effect sizes if they were adjusted for similar variables in the individual studies after documenting the variables adjusted for in the studies. We considered a two-sided probability of $< 0.05$ as significant. We performed the analysis of heterogeneity using the $I^2$ statistics. When the $I^2$ was $> 60\%$, we performed subgroup analyses and meta-regression with respect to the TB-HIV co-infection status, resistance to ATT, and extra-pulmonary involvement. We also performed subgroup analysis based on characteristics of the study country such as income status classification according to World Bank[405], incidence of TB infection and incidence of TB-HIV co-infection[406]. We also performed meta-regression using data such as time point (in years) of assessment of mortality in each study, mean age, and the proportion of HIV and other comorbidities (diabetes, hypertension, cardiovascular diseases, chronic obstructive pulmonary disease (COPD), smoking, and alcohol use) in the study population. We assessed the impact of the difference in the log-odds of treatment completion, default, and lost to follow up between males and females using meta-regression. We performed sensitivity analyses by excluding studies that looked at mortality in patients in the intensive care unit. Publication bias was assessed by funnel plot, and Egger’s test. We performed the analysis using STATA 16-1C. The study protocol is registered with PROSPERO (CRD42020219050).

**Role of the funding source**

The funder had no role in study design, data collection, data analysis, data interpretation, or writing of the report.

### Section IIb

#### Supplementary Results

Subgroup analysis based on the income status of the study country showed that high-income countries had a higher OR of mortality (OR 1.39, 95%CI 1.26-1.52) in males compared to females when compared to low-income (OR 1.19, 95%CI 1.03-1.38) and middle-income (OR 1.20, 95%CI 1.08-1.32) countries. Subgroup analysis based on the TB and TB-HIV burden showed that low-burden countries had a higher OR of mortality in males compared to females when compared to high-burden countries. Subgroup analysis based on the continent of study showed that all regions except South America consistently showed higher pooled effect sizes among males. Implementation of the DOTS program did not change the association between male sex and higher mortality. Both studies reporting on drug-resistant and those reporting on drug-susceptible TB consistently showed higher mortality in males. Studies focused exclusively on HIV-TB co-infected individuals failed to show a direction of association between mortality and sex. Subgroup analysis based on study design did not show a difference among the cohort and case-control studies, in the association between mortality and male sex.

Meta-regression analysis did not result in a statistically significant association between difference in rates of default, loss to follow up and treatment completion between males and females, and the OR for mortality in males (Supplementary table 4). However, an increase in the HIV proportion in females compared to males resulted in a decrease in the OR for overall mortality in males following meta-regression (Supplementary table 4).

### **Section III**

#### Supplementary Tables

**Supplementary Table 1: Age-adjusted effect sizes for the association of male sex with the outcomes (Taiwan Cohort)**

| Characteristic | Estimate | Adjusted Effect size[#] | 95%CI | p-value | Age adjusted effect size[&] | 95%CI | p-value |
|---|---|---|---|---|---|---|---|
| **All-cause mortality** | HR | 1.43 | 1.03-1.98 | 0.032 | 1.45 | 1.04-2.04 | 0.027 |
| **Infection related mortality** | HR | 1.70 | 1.09-2.64 | 0.009 | 1.69 | 1.09-2.63 | 0.020 |
| **2-month Sputum culture positivity** | OR | 1.56 | 1.05-2.33 | 0.028 | 1.59 | 1.02-2.43 | 0.043 |
| **2-month Sputum smear AFB** | OR | 1.27 | 0.71-2.27 | 0.42 | 1.43 | 0.75-2.77 | 0.290 |

AFB= Acid Fast Bacilli. CI= Confidence Interval. HR= Hazard ratio. OR= Odds Ratio.

**Supplementary Table 2: Study characteristics for the studies included in our systematic review.**

| Name of the Author | Year of publication | Country of study | Study Period (From) | Study Period (To) | Outcomes | | | | | Type of study | Data collection | Drug resistance | TB-HIV co-infection (%) | EPTB (%) | Age (years) Mean / Median* | Number of Male | Number of Female | NOS (Score) |
|---|---|---|---|---|---|---|---|---|---|---|---|---|---|---|---|---|---|---|
| | | | | | All-cause mortality | Death due to TB | Sputum Culture | Sputum Smear | Treatment success | | | | | | | | | |
| Sariem[1] | 2020 | Nigeria | 2001 | 2015 | | | | | * | RC | Hosp | 6 | 44.29 | 11.3 | 35.5 | 5904 | 4252 | 8 |
| Vo[2] | 2020 | Vietnam | 2014 | NA | | | | | * | RC | Prog | 6 | 11 | 25 | 41 | 3791 | 1711 | 8 |
| Stosic[3] | 2020 | Serbia | 2005 | 2015 | * | | | | * | RC | Prog | 6 | NA | 12.6 | 46 | 1444 | 934 | 8 |
| Izudi[4] | 2020 | Uganda | 2010 | 2018 | | | | | * | RC | Hosp | 6 | 40.1 | 0 | 38.5 | 141 | 46 | 9 |
| Ahmad[5] | 2020 | Pakistan | 2011 | 2014 | | | | | * | RC | Hosp | 6 | NA | 35.2 | ·· | 120 | 132 | 8 |
| Matambo[6] | 2020 | Zimbabwe | 2010 | 2015 | | | | | * | RC | Prog | 1 | 77.7 | 0 | 34 | 230 | 241 | 7 |
| Hodgkinson[7] | 2020 | Kenya | 2004 | 2017 | * | | | | | RC | Hosp | 6 | 100 | ·· | ·· | 1129 | 2543 | 8 |
| Wang[8] | 2020 | China | 2006 | 2011 | * | | | | | RC | Hosp | 1 | 0 | ·· | 46.45 | 262 | 94 | 8 |
| Liu[9] | 2020 | China | 2018 | 2019 | * | | | | | RC | Hosp | 6 | 0 | 0 | 57.82 | 101 | 52 | 8 |
| Geleso[10] | 2020 | Ethiopia | 2016 | 2017 | * | | | | | RC | Hosp | 6 | 26.7 | 28.7 | ·· | 225 | 172 | 8 |
| Washington[11] | 2020 | India | 2018 | 2019 | * | | | | * | PC | Hosp | 6 | 1.8 | 24.4 | ·· | 2952 | 1797 | 8 |
| Khunthason[12] | 2020 | Thailand | 2014 | 2017 | | | | | * | RC | Prog | 6 | 6 | 12.3 | ·· | 492 | 261 | 7 |
| Arpagaus[13] | 2020 | Tanzania | 2013 | 2017 | | | | | * | PC | Hosp | 6 | 100 | 30.48 | 36.5* | 1111 | 2018 | 8 |
| Zheng[14] | 2020 | China | 2014 | 2015 | | | | | * | PC | Hosp | 2 | NA | 0 | 45.4 | 49 | 9 | 8 |
| Du[15] | 2019 | China | 2008 | 2010 | | | | | * | PC | Hosp | 0 | NA | 0 | 44 | 318 | 174 | 8 |
| Ramos[16] | 2020 | Ethiopia | 1998 | 2015 | * | | | | | RC | Hosp | 6 | 5.3 | 31.26 | ·· | 1172 | 1080 | 6 |
| Van[17] | 2020 | Vietnam | 2011 | 2015 | | | | | * | RC | Hosp | 2 | 9.6 | 3 | 43 | 1715 | 551 | 8 |

| | | | | | | | | | | | | | | | | | | |
|---|---|---|---|---|---|---|---|---|---|---|---|---|---|---|---|---|---|---|
| Singla[18] | 2020 | India | 2017 | 2019 | * | | | | | CC | Hosp | 6 | NA | 0 | ·· | 150 | 80 | 8 |
| Shao[19] | 2020 | China | 2013 | 2018 | | | | * | * | RC | Hosp | 3 | NA | 0 | 48* | 47 | 16 | 8 |
| Olayanju[20] | 2019 | South Africa | 2014 | 2018 | | | | | * | PC | Hosp | 1 | 52.45 | ·· | 33* | 74 | 48 | 8 |
| Lee[21] | 2020 | South Korea | 2012 | 2017 | | | * | | | RC | Hosp | 0 | 0 | 0 | 49* | 48 | 19 | 8 |
| Schwœbel[22] | 2020 | Multicountry | 2013 | 2015 | | | | | * | RC | Prog | 2 | 19.8 | ·· | ·· | 668 | 338 | 7 |
| Humphrey[23] | 2020 | Multicountry | 2012 | 2014 | * | | | | | RC | Prog | 6 | 100 | 21 | 36* | 1181 | 910 | 7 |
| Piubello[24] | 2019 | Nigeria | 2008 | 2016 | | | | | * | RC | Hosp | 2 | 0.04 | 68.2 | 32 | 206 | 43 | 8 |
| Shi[25] | 2020 | China | 2012 | 2015 | | | | | * | PC | Hosp | 2 | NA | ·· | 47.6* | 161 | 81 | 8 |
| Gao[26] | 2020 | China | 2018 | 2019 | | | | | * | RC | Hosp | 1 | 0.6 | ·· | 40* | 132 | 45 | 8 |
| Rizvi[27] | 2020 | India | 2012 | 2018 | * | | | | | PC | Hosp | 6 | 2.8 | 100 | 31.12 | 368 | 353 | 8 |
| Charoensakulchai[28] | 2020 | Thailand | 2019 | 2019 | | | | | * | RC | Hosp | 0 | NA | 0 | 52* | 554 | 232 | 8 |
| Lakoh[29] | 2020 | Sierra Leone | 2017 | 2017 | * | | | | * | RC | Hosp | 0 | 31.9 | 3.7 | ·· | 766 | 339 | 8 |
| Alene[30] | 2019 | Multicountry | 2010 | 2014 | * | | | | * | RC | Hosp | 2 | NA | 0 | 38 | 211 | 114 | 6 |
| Tok[31] | 2020 | Malaysia | 2014 | 2017 | | | | | * | RC | Prog | 0 | 6.0 | 13.0 | 42.7 | 62660 | 34845 | 7 |
| Lin[32] | 2020 | China | 2015 | 2016 | | | | | * | PC | Hosp | 0 | NA | ·· | 53 | 215 | 91 | 8 |
| Schmit[33] | 2020 | USA | 2011 | 2016 | * | | | | | RC | Prog | 6 | 100 | 20.0 | ·· | 30408 | 18596 | 7 |
| Gonah[34] | 2020 | Zimbabwe | 2013 | 2016 | * | | | | * | RC | Prog | 2 | 0.52 | ·· | ·· | 89 | 85 | 7 |
| Tanue[35] | 2019 | Cameroon | 2010 | 2017 | | | | | * | RC | Hosp | 6 | 100 | 0 | 37.07 | 450 | 591 | 8 |
| Makhmudova[36] | 2019 | Tajikistan | 2012 | 2013 | * | | | * | | RC | Hosp | 1 | 2.5 | 0 | 32 | 342 | 259 | 8 |
| Cheng[37] | 2019 | Taiwan | 2006 | 2016 | * | | | | | RC | Hosp | 6 | NA | 46 | 53.2* | 9 | 21 | 8 |
| Pradipta[38] | 2018 | Netherlands | 2005 | 2015 | | | | | * | RC | Prog | 0 | 4 | 46.9 | ·· | 3426 | 2248 | 7 |
| Holden[39] | 2019 | Denmark | 2009 | 2014 | | | | | * | RC | Prog | 0 | 3 | 0 | 44* | 1082 | 599 | 7 |
| Arroyo[40] | 2019 | Brazil | 2006 | 2015 | * | | | | | RC | Prog | 2 | 13 | 2.5 | ·· | 564 | 238 | 7 |
| Bhering[41] | 2019 | Brazil | 2000 | 2016 | * | | | | * | RC | Prog | 1 | 7.9 | 2.3 | ·· | 1466 | 803 | 7 |

| | | | | | | | | | | | | | | | | | | |
|---|---|---|---|---|---|---|---|---|---|---|---|---|---|---|---|---|---|---|
| Cohen[42] | 2019 | Malawi | 2013 | 2014 | | | | | * | PC | Hosp | 0 | 82.9 | 16.5 | 37* | 102 | 56 | 5 |
| Bhering[43] | 2019 | Portugal | 2000 | 2014 | * | | | | * | RC | Prog | 2 | 39.8 | 9.4 | 39* | 180 | 85 | 7 |
| Komiya[44] | 2020 | Japan | 2013 | 2015 | * | | * | | | RC | Hosp | 6 | NA | 0 | 82* | 94 | 91 | 8 |
| Mahwire[45] | 2019 | South Africa | 2012 | 2014 | | | | * | | RC | Hosp | 2 | 76.3 | 0 | ·· | 357 | 361 | 5 |
| Shariff[46] | 2019 | Malaysia | 2009 | 2013 | | | | | * | RC | Hosp | 1 | 0 | 2.4 | ·· | 303 | 100 | 5 |
| Bouton[47] | 2019 | Ghana | 2010 | 2016 | * | | | | | RC | Hosp | 6 | 24.0 | 30.4 | 41.8 | 264 | 130 | 5 |
| Ogyiri[48] | 2019 | Ghana | 2013 | 2015 | | | | | * | RC | Hosp | 0 | 20.3 | 24.2 | ·· | 344 | 182 | 6 |
| Zürcher[49] | 2019 | Multicountry | 2013 | 2016 | * | | | | | PC | Hosp | 6 | 43 | ·· | 33.2* | 362 | 272 | 8 |
| Pettit[50] | 2019 | Multicountry | 2012 | 2013 | | | | | * | RC | Hosp | 0 | 100 | 24 | ·· | 1089 | 773 | 8 |
| Zurcher[51] | 2019 | Multicountry | 2012 | 2014 | * | | | | | RC | Hosp | 0 | 100 | 28 | 35.5* | 1593 | 1102 | 8 |
| Golub[52] | 2019 | South Korea | 2001 | 2011 | | * | | | | PC | Hosp | 6 | NA | ·· | 46 | 819051 | 448513 | 8 |
| Byashalira[53] | 2020 | Tanzania | 2018 | 2018 | * | | | | | PC | Hosp | 0 | 100 | ·· | 40* | 45 | 52 | 8 |
| Min[54] | 2019 | South Korea | 2014 | 2017 | * | | | | | RC | Hosp | 0 | 0.9 | 22.9 | 83.8 | 55 | 54 | 8 |
| Batool[55] | 2019 | Pakistan | 2008 | 2016 | | | | | * | RC | Hosp | 1 | NA | 0 | 29 | 99 | 94 | 8 |
| Hamdouni[56] | 2019 | Morocco | 2014 | 2016 | | | | | * | RC | Hosp | 1 | NA | 42.1 | 35.5* | 72 | 79 | 8 |
| Khan[57] | 2019 | Malaysia | 2006 | 2008 | | | | | * | RC | Hosp | 6 | 15.38 | 100 | ·· | 778 | 444 | 8 |
| Tola[58] | 2019 | Ethiopia | 2012 | 2017 | | | | | * | RC | Hosp | 6 | 100 | 34.1 | ·· | 161 | 188 | 8 |
| Musaazi[59] | 2018 | Uganda | 2009 | 2015 | * | | | | | RC | Hosp | 6 | 100 | 41.6 | ·· | | | 8 |
| Balaky[60] | 2019 | Iraq | 2012 | 2019 | * | | | | | RC | Hosp | 6 | NA | 63.5 | 40.5 | 320 | 408 | 6 |
| Aguilar[61] | 2019 | Brazil | 2007 | 2015 | | | | | * | CC | Hosp | 6 | NA | 0 | ·· | 179 | 105 | 8 |
| Ali[62] | 2019 | Sudan | 2013 | 2017 | | | | | * | PC | Hosp | 2 | 1.9 | 2.6 | ·· | 117 | 39 | 8 |
| Pedrazzoli[63] | 2019 | UK | 2001 | 2014 | * | | | | | RC | Prog | 6 | 5 | 44.03 | ·· | 62628 | 49146 | 7 |
| Dedefo[64] | 2019 | Ethiopia | 2014 | 2017 | * | | | * | * | RC | Hosp | 6 | 32.9 | ·· | 37.91 | 103 | 96 | 5 |
| Pradipta[65] | 2019 | Netherlands | 2005 | 2015 | * | | | | * | RC | Prog | 1 | NA | 48.1 | ·· | 295 | 250 | 7 |

| | | | | | | | | | | | | | | | | | | |
|---|---|---|---|---|---|---|---|---|---|---|---|---|---|---|---|---|---|---|
| Somsong[66] | 2018 | Thailand | 2014 | 2015 | * | | | | * | RC | Prog | 6 | NA | 0 | .. | 13505 | 6636 | 7 |
| Nguyen[67] | 2018 | USA | 2010 | 2016 | * | | | | | RC | Prog | 6 | 6.3 | 14.2 | .. | 5872 | 3130 | 7 |
| Nandasena[68] | 2018 | Sri Lanka | 2013 | 2013 | | | | * | | PC | Prog | 6 | NA | 29.75 | .. | 451 | 218 | 7 |
| Prudhivi[69] | 2019 | India | 2014 | 2016 | * | | | | * | RC | Hosp | 6 | 23 | 0 | 47.13 | 734 | 379 | 8 |
| Zhang[70] | 2018 | China | 2007 | 2017 | * | | | | | RC | Prog | 6 | 100 | .. | 38.01 | 605 | 123 | 7 |
| Rossetto[71] | 2018 | Brazil | 2009 | 2013 | * | | | | | RC | Prog | 6 | 100 | 0 | .. | 1588 | 831 | 7 |
| Ohene[72] | 2018 | Ghana | 2010 | 2013 | * | | | | | RC | Prog | 6 | 40.84 | 100 | 40.4 | 2090 | 1243 | 7 |
| Khan[73] | 2019 | Pakistan | 2012 | 2016 | | | | | * | RC | Hosp | 2 | 1 | 0 | 37.07 | 72 | 114 | 8 |
| Holmberg[74] | 2019 | Finland | 1998 | 2015 | * | | | | | RC | Prog | 6 | 100 | .. | .. | 451 | 218 | 7 |
| Sadykova[75] | 2019 | Kazakhstan | 2014 | 2016 | | | | | * | RC | Hosp | 0 | 3.4 | 11.2 | .. | 22648 | 14278 | 8 |
| Dangeti[76] | 2017 | India | 2014 | 2015 | | | | | * | CC | Hosp | 6 | 0 | 100 | .. | 28 | 12 | 8 |
| Javaid[77] | 2018 | Pakistan | 2012 | 2014 | | | * | | | RC | Hosp | 2 | 1 | 0 | 30.7 | 189 | 239 | 8 |
| Ambaw[78] | 2017 | Ethiopia | 2014 | 2016 | | | | | * | PC | Hosp | 0 | 11.4 | 42.8 | 30 | 348 | 300 | 8 |
| Parmar[79] | 2018 | India | 2007 | 2011 | | | * | | * | RC | Prog | 2 | 1.6 | 0 | 35 | 2564 | 1148 | 7 |
| Adane[80] | 2018 | Ethiopia | 2010 | 2015 | | | | | * | RC | Hosp | 6 | 11 | 45 | .. | 480 | 16 | 8 |
| Worku[81] | 2018 | Ethiopia | 2008 | 2016 | | | | | * | RC | Hosp | 6 | 24.2 | 36.9 | .. | 516 | 469 | 8 |
| Tafess[82] | 2018 | Ethiopia | 2004 | 2014 | * | | | | * | RC | Hosp | 6 | 16.4 | 36.1 | 25 | 248 | 232 | 4 |
| Javaid[83] | 2017 | Pakistan | 2012 | 2014 | | | | | * | RC | Hosp | 2 | NA | 1.3 | .. | 235 | 300 | 8 |
| Muyaya[84] | 2018 | Botswana | 2013 | 2013 | * | | | | | RC | Hosp | 6 | 100 | 27 | .. | 170 | 130 | 8 |
| Ferreira[85] | 2018 | Brazil | 2011 | 2014 | * | | | | | RC | Hosp | 6 | 71.6 | 52.5 | 37 | 84 | 36 | 5 |
| Muluye[86] | 2018 | Ethiopia | 2012 | 2016 | | | | | * | RC | Hosp | 6 | 6.8 | 43.1 | .. | 586 | 409 | 5 |
| Tshitenge[87] | 2018 | Botswana | 2013 | 2015 | * | | | | | RC | Hosp | 6 | 36.1 | 18 | .. | 608 | 478 | 5 |
| Evans[88] | 2018 | South Africa | 2011 | 2014 | * | | | | * | RC | Prog | 6 | 26.1 | .. | 36* | 704 | 603 | 7 |
| Adamu[89] | 2018 | Nigeria | 2010 | 2014 | | | | | * | RC | Hosp | 6 | 39.8 | 32.19 | .. | 596 | 785 | 8 |

| | | | | | | | | | | | | | | | | | | |
|---|---|---|---|---|---|---|---|---|---|---|---|---|---|---|---|---|---|---|
| Kaplan[90] | 2018 | South Africa | 2009 | 2013 | * | | | | | RC | Hosp | 6 | 50.8 | 20.5 | ·· | 35098 | 25384 | 8 |
| Bulabula[91] | 2019 | Democratic Republic of Congo | 2012 | 2017 | | | | | * | RC | Prog | 2 | 0.8 | 0 | 35* | 9029 | 7330 | 7 |
| Frank[92] | 2019 | Georgia | 2011 | 2013 | * | | | | * | RC | Hosp | 4 | 2.9 | 11.7 | 34 | 77 | 34 | 8 |
| Garg[93] | 2019 | India | 2017 | 2017 | | | | | * | RC | Hosp | 3 | NA | 0 | ·· | 35 | 17 | 6 |
| Lee[94] | 2019 | South Korea | 2005 | 2017 | | | | | * | RC | Hosp | 1 | NA | ·· | 39.7 | 79 | 50 | 8 |
| Crabtree-Ram ́ırez[95] | 2018 | Multicountry | 2000 | 2015 | * | | | | | RC | Hosp | 6 | 100 | 53 | 35* | 581 | 178 | 8 |
| Han[96] | 2017 | South Korea | 1996 | 2015 | * | | | | | RC | Hosp | 6 | NA | ·· | 61 | 44 | 52 | 8 |
| Ejeta[97] | 2018 | Ethiopia | 2008 | 2017 | | | | | * | RC | Hosp | 6 | 27.9 | 18.2 | ·· | 2306 | 1838 | 8 |
| Diallo[98] | 2018 | Burkina Faso | 2010 | 2014 | | | | * | | CC | Hosp | 6 | 8.9 | 0 | ·· | 428 | 258 | 8 |
| Melese[99] | 2018 | Ethiopia | 2008 | 2013 | * | | | * | * | RC | Hosp | 6 | 13.5 | 38.6 | 34.9 | 173 | 130 | 8 |
| Viana[100] | 2017 | Brazil | 2012 | 2013 | * | | | | | RC | Prog | 1 | 5.4 | 1.2 | ·· | 179 | 78 | 7 |
| Wu[101] | 2018 | China | 2011 | 2016 | * | | | | | RC | Hosp | 6 | NA | 16.7 | 45.1 | 39 | 9 | 8 |
| Feng[102] | 2018 | Taiwan | 2012 | 2015 | * | | * | | | PC | Hosp | 6 | NA | 17.5 | 64 | 140 | 72 | 8 |
| Sekaggya-Wiltshire[103] | 2018 | Switzerland | 2013 | 2015 | | | * | | * | PC | Hosp | 6 | 100 | 0 | ·· | 134 | 93 | 8 |
| Khac Thai[104] | 2018 | Vietnam | 2008 | 2011 | | | | | * | RC | Hosp | 3 | 0 | 0 | ·· | 177 | 62 | 8 |
| Shimazaki[105] | 2017 | Phillipines | 2011 | 2013 | * | | | | | PC | Hosp | 6 | 0 | 0 | 46.9 | 317 | 149 | 8 |
| Nguyen[106] | 2018 | USA | 2010 | 2014 | * | | | | | RC | Prog | 6 | 6 | 28 | 52* | 5132 | 3329 | 7 |
| Mishkin[107] | 2018 | Kazakhstan | 2014 | 2015 | * | | | | * | RC | Prog | 6 | 100 | ·· | ·· | 239 | 79 | 7 |
| Azeez[108] | 2018 | South Africa | 2010 | 2016 | | | | | * | RC | Hosp | 1 | 100 | ·· | ·· | 530 | 380 | 8 |
| Velavan[109] | 2018 | India | 2014 | 2015 | | | | | * | RC | Hosp | 6 | 1 | 7.98 | ·· | 319 | 73 | 8 |
| Yu [110] | 2018 | Taiwan | 2007 | 2012 | * | | | * | | RC | Hosp | 2 | NA | 0 | ·· | 500 | 186 | 8 |
| Lin[111] | 2017 | Taiwan | 2007 | 2017 | | | | | * | RC | Hosp | 2 | 0 | 0 | ·· | 122 | 45 | 8 |
| Lv[112] | 2018 | China | 2011 | 2012 | | | | | * | PC | Hosp | 2 | NA | 0 | ·· | 33 | 59 | 8 |
| Kuehne[113] | 2018 | Germany | 2002 | 2014 | | | | | * | RC | Prog | 6 | NA | 0 | ·· | 1138 | 336 | 6 |

| | | | | | | | | | | | | | | | | | | |
|---|---|---|---|---|---|---|---|---|---|---|---|---|---|---|---|---|---|---|
| Hameed[114] | 2019 | Pakistan | 2018 | 2019 | * | | | | | RC | Hosp | 0 | 1.8 | 0 | ·· | 92 | 78 | 5 |
| Eugene Wickett[115] | 2018 | Liberia | 2015 | 2017 | * | | | | | RC | Hosp | 6 | NA | ·· | ·· | 285 | 275 | 8 |
| L. Zhang[116] | 2017 | China | 2009 | 2013 | | | | | * | PC | Hosp | 2 | NA | 0 | ·· | 382 | 155 | 8 |
| Jose Gabriel Cornejo Garcia[117] | 2018 | Peru | 2012 | 2014 | * | | * | | | RC | Prog | 3 | 4.3 | 0 | ·· | 652 | 295 | 7 |
| Mok[118] | 2018 | South Korea | 2014 | 2015 | * | | | | * | RC | Prog | 0 | NA | 20.3 | 52.5 | 2765 | 1967 | 7 |
| Trébucq[119] | 2017 | Multicountry | 2013 | 2014 | | | | | * | PC | Hosp | 2 | 19.9 | 0 | ·· | 667 | 339 | 7 |
| Fitsum Weldegebreal[120] | 2018 | Ethiopia | 2008 | 2014 | * | | | * | * | RC | Hosp | 6 | 100 | 44.8 | 32.52 | 362 | 265 | 8 |
| Ige[121] | 2018 | Nigeria | 2010 | 2013 | | | | | * | RC | Hosp | 2 | 19.1 | 0 | ·· | 76 | 39 | 8 |
| Chingonzoh[122] | 2018 | South Africa | 2011 | 2013 | * | | | | | RC | Prog | 1 | 65 | 0 | 37* | 1992 | 1737 | 7 |
| Ji[123] | 2018 | China | 2011 | 2015 | * | | | | | RC | Hosp | 6 | 100 | 0 | 39 | 325 | 34 | 5 |
| Velayutham[124] | 2018 | India | NA | NA | | | | | * | PC | Hosp | 6 | 0.9 | 0 | ·· | 1125 | 440 | 8 |
| Obregón[125] | 2018 | Peru | 2010 | 2013 | | | | | * | RC | Prog | 2 | 2.8 | 0 | 27* | 940 | 490 | 7 |
| Alarcón[126] | 2018 | Peru | 2011 | 2014 | | | | | * | RC | Prog | 1 | 4 | 0 | ·· | 415 | 195 | 7 |
| Nguyen[127] | 2016 | USA | NA | NA | * | | | | | RC | Prog | 0 | 100 | ·· | ·· | 346 | 104 | 7 |
| Thao[128] | 2017 | Vietnam | 2001 | 2015 | * | | | | | RC | Hosp | 6 | 44.025 | 100 | ·· | 1185 | 514 | 7 |
| Fan[129] | 2017 | China | 2011 | 2013 | | | | | * | RC | Hosp | 2 | NA | 0 | ·· | 127 | 52 | 7 |
| Kibuule[130] | 2018 | Namibia | 2004 | 2016 | | | | | * | RC | Prog | 6 | 47.9 | 19.2 | ·· | 44300 | 32556 | 7 |
| Pizzol[131] | 2018 | Mozambique | 2016 | 2016 | | | | * | | PC | Hosp | 6 | 43.5 | 0 | 31 | 203 | 98 | 7 |
| Kirirabwa[132] | 2018 | Uganda | 2014 | 2015 | | | | | * | RC | Hosp | 6 | 49.9 | 16.5 | ·· | 312 | 202 | 7 |
| Wakamatsu[133] | 2018 | Japan | 1994 | 2016 | * | | | | | RC | Hosp | 6 | NA | 100 | 83 | 18 | 50 | 8 |
| Silva[134] | 2018 | Brazil | 2007 | 2014 | * | | | | | RC | Hosp | 6 | 100 | 48.7 | ·· | 672 | 252 | 8 |
| Aibana[135] | 2019 | Ukraine | 2012 | 2015 | * | | | | * | RC | Hosp | 0 | 16.2 | 0 | 40 | 351 | 111 | 6 |
| Singhi[136] | 2018 | India | 2015 | 2015 | | | | | * | RC | Hosp | 6 | 0.1 | 33 | ·· | 793 | 697 | 5 |
| Wen[137] | 2018 | China | 2005 | 2013 | | | | | * | RC | Prog | 6 | NA | 0 | 51* | 16939 | 6059 | 7 |

| | | | | | | | | | | | | | | | | | | |
|---|---|---|---|---|---|---|---|---|---|---|---|---|---|---|---|---|---|---|
| Ratan Kumar[138] | 2018 | India | 2012 | 2015 | * | | | * | * | RC | Hosp | 6 | 100 | 65.5 | .. | 266 | 188 | 8 |
| Engelbrecht[139] | 2017 | Malaysia | 2009 | 2012 | | | | | * | RC | Prog | 6 | 6 | 23.3 | .. | 5276 | 4715 | 7 |
| Suryawanshi[140] | 2017 | india | 2011 | 2012 | * | | | | * | RC | Hosp | 2 | 5.6 | 2.55 | .. | 2009 | 1401 | 8 |
| Mattila[141] | 2016 | Finland | 1978 | 1980 | * | | | | | RC | Hosp | 6 | NA | 0 | .. | 3125 | 3576 | 8 |
| Cardoso[142] | 2017 | Brazil | 2010 | 2014 | * | | | * | | RC | Prog | 6 | 25.3 | 32.25 | 40.32 | 114 | 108 | 7 |
| Tatar[143] | 2017 | Turkey | 2004 | 2010 | * | | | | | RC | Hosp | 6 | NA | 0 | 54 | 33 | 7 | 8 |
| Alene[144] | 2018 | Ethiopia | 2010 | 2011 | | | * | * | | RC | Hosp | 2 | NA | 0 | .. | 239 | 107 | 8 |
| Nagu[145] | 2017 | Tanzania | 2010 | 2011 | * | | | | | PC | Hosp | 6 | 100 | 0 | .. | 1138 | 558 | 8 |
| Nagu[146] | 2017 | Tanzania | 2014 | 2015 | * | | | | | RC | Hosp | 6 | 30.6 | 7.6 | 48.4 | 168 | 85 | 8 |
| Lee[147] | 2017 | South Korea | 2006 | 2011 | | | | | * | RC | Hosp | 6 | NA | 0 | 35 | 39 | 37 | 8 |
| Bastos[148] | 2017 | Brazil | 2007 | 2013 | | | | | * | RC | Prog | 2 | 9 | 3 | 39.5 | 1310 | 662 | 7 |
| Alene[149] | 2017 | Ethiopia | 2010 | 2015 | | | | | * | RC | Hosp | 6 | 21.1 | 6.2 | .. | 147 | 95 | 8 |
| Ahmad[150] | 2017 | Pakistan | 2011 | 2012 | | | | | * | RC | Hosp | 6 | NA | 22.7 | .. | 259 | 234 | 8 |
| Schechter[151] | 2017 | USA | 2008 | 2015 | | | * | | | RC | Hosp | 6 | 37 | 0 | 47.6 | 182 | 54 | 8 |
| Sinshaw[152] | 2017 | Ethiopia | 2010 | 2016 | | | | | * | RC | Hosp | 6 | 100 | 28.8 | .. | 175 | 133 | 6 |
| Aibana[153] | 2017 | Ukraine | 2012 | 2015 | * | | | * | * | RC | Hosp | 2 | 21.7 | .. | 38.2* | 292 | 86 | 7 |
| Loh[154] | 2017 | Singapore | 2005 | 2010 | * | | | | | RC | Hosp | 1 | 5.3 | 0 | 59.9 | 50 | 25 | 7 |
| Nagu[155] | 2016 | Tanzania | 2010 | 2011 | * | | | | * | PC | Hosp | 6 | 29.1 | .. | .. | 579 | 282 | 7 |
| Getnet[156] | 2017 | Ethiopia | 2009 | 2014 | | | | | * | RC | Hosp | 6 | 4.6 | 25.8 | .. | 814 | 564 | 7 |
| Ko[157] | 2017 | Taiwan | 2000 | 2010 | * | | | | | RC | Prog | 6 | 0.5 | .. | 67.3 | 1878 | 860 | 7 |
| Yen[158] | 2017 | Taiwan | 2006 | 2014 | * | | | | | RC | Hosp | 6 | NA | 7.44 | .. | 3712 | 1299 | 8 |
| Raberahona,[159] | 2017 | Madagascar | 2007 | 2014 | * | | | | | RC | Hosp | 6 | 4 | 100 | .. | 42 | 33 | 8 |
| Prado[160] | 2016 | Brazil | 2001 | 2011 | * | * | | | | RC | Prog | 6 | 100 | 30.8 | .. | 26692 | 10745 | 7 |
| Sam[161] | 2018 | Cambodia | 2006 | 2016 | | | | | * | RC | Hosp | 2 | 20 | 3.7 | 45 | 369 | 213 | 8 |

| | | | | | | | | | | | | | | | | | |
|---|---|---|---|---|---|---|---|---|---|---|---|---|---|---|---|---|---|
| Kuhlin[162] | 2017 | Uzbekistan | 2003 | 2016 | | | | | * | RC | Prog | 2 | NA | 0 | 30.5* | 1189 | 1257 | 8 |
| Ndjeka[163] | 2018 | South Africa | 2014 | 2016 | * | | | * | * | PC | Hosp | 1 | 67 | 0 | 27* | 101 | 99 | 8 |
| Christopher Martin Sauer[164] | 2018 | Multicountry | 2000 | 2016 | | | | | * | RC | Prog | 6 | NA | 33.9 | .. | 440 | 203 | 7 |
| Diktanas[165] | 2018 | Lithuania | 2015 | 2016 | | | * | | | RC | Hosp | 6 | 0 | 1.17 | 49.5 | 59 | 28 | 8 |
| Nguyen[166] | 2018 | USA | 2010 | 2016 | * | | | | | RC | Prog | 6 | 8 | 13.92 | .. | 2191 | 1186 | 7 |
| Mundra[167] | 2017 | India | 2015 | 2016 | | | | | * | CC | Hosp | 6 | 5.4 | 20.36 | .. | 171 | 104 | 8 |
| Asres[168] | 2018 | Ethiopia | 2015 | 2016 | | | | | * | PC | Hosp | 6 | 9.3 | 20.3 | .. | 2943 | 1892 | 6 |
| Gaborit[169] | 2018 | France | 2002 | 2013 | | | | * | | RC | Prog | 2 | 8.9 | 12.2 | .. | 100 | 34 | 7 |
| Jackson[170] | 2017 | India | 2012 | 2014 | | | | | * | RC | Hosp | 6 | NA | 35 | .. | 4907 | 3508 | 8 |
| Piparva[171] | 2017 | India | 2013 | 2014 | | | | | * | RC | Hosp | 0 | 2.83 | 21.99 | .. | 952 | 388 | 8 |
| Wang[172] | 2017 | China | 2009 | 2011 | | | | | * | RC | Hosp | 6 | NA | 0 | 43 | 300 | 95 | 8 |
| Wondale[173] | 2017 | Ethiopia | 2004 | 2014 | * | | | | * | RC | Hosp | 6 | 14.6 | 25.9 | 30.1 | 707 | 465 | 7 |
| Kefale[174] | 2017 | Ethiopia | 2012 | 2015 | * | | | | * | RC | Hosp | 6 | 100 | 14.36 | 30.55 | 97 | 91 | 8 |
| Gunda[175] | 2017 | Tanzania | 2016 | 2017 | | | | * | | PC | Hosp | 6 | 35.26 | 0 | 39 | 97 | 59 | 8 |
| Nair[176] | 2016 | India | 2009 | 2011 | | | | | * | RC | Prog | 2 | 2 | .. | .. | 535 | 252 | 7 |
| Kapata[177] | 2017 | Zambia | 2012 | 2014 | * | | | | | RC | Prog | 2 | 73.47 | .. | 36* | 41 | 26 | 7 |
| Huerga[178] | 2016 | Kenya | 2006 | 2012 | | | | | * | RC | Hosp | 2 | 25.6 | .. | .. | 93 | 76 | 8 |
| Mehta[179] | 2017 | India | 2002 | 2009 | * | | | | | RC | Hosp | 0 | 11.4 | 100 | .. | 29 | 28 | 8 |
| Mert[180] | 2017 | Turkey | 1981 | 2015 | * | | | | | RC | Hosp | 0 | 6 | 100 | .. | 142 | 121 | 8 |
| Heijden[181] | 2017 | South Africa | 2000 | 2012 | | | | | * | RC | Hosp | 6 | 79 | 5 | .. | 11004 | 5864 | 8 |
| Georghiou[182] | 2017 | USA | 2012 | 2013 | * | | | | | PC | Hosp | 1 | 15 | 0 | .. | 296 | 155 | 6 |
| Seifert[183] | 2017 | Multicountry | 2012 | 2013 | * | | | | | PC | Hosp | 6 | 11 | .. | .. | 542 | 292 | 7 |
| Shin[184] | 2017 | Botswana | 2006 | 2013 | * | | | | * | RC | Prog | 2 | 44.4 | 7.1 | 36* | 322 | 265 | 7 |
| Thu[185] | 2018 | Myanmar | 2012 | 2014 | | | | | * | RC | Prog | 2 | 10 | .. | 38 | 1399 | 786 | 7 |

| | | | | | | | | | | | | | | | | | | |
|---|---|---|---|---|---|---|---|---|---|---|---|---|---|---|---|---|---|---|
| Janssen[186] | 2016 | South Africa | 2014 | 2014 | * | | | | | PC | Hosp | 0 | 100 | ·· | ·· | 26 | 33 | 8 |
| Teklu[187] | 2017 | Ethiopia | 2005 | NA | * | | | | | RC | Hosp | 6 | 100 | 6.8 | 32* | 1148 | 2441 | 8 |
| Dale[188] | 2017 | Australia | 2002 | 2015 | * | | | | | RC | Prog | 0 | 1.5 | 45 | ·· | 2655 | 2212 | 7 |
| Ramachandran[189] | 2017 | India | 2013 | 2015 | | | | | * | PC | Hosp | 6 | 1.0 | 35 | ·· | 1291 | 621 | 8 |
| M[190] | 2017 | Nigeria | 2010 | 2014 | * | | | | * | RC | Prog | 6 | 20.6 | 3.7 | ·· | 553 | 410 | 7 |
| El-Shabrawy[191] | 2016 | Egypt | 2013 | 2014 | | | | | * | RC | Hosp | 0 | 0 | 11.5 | 35.7 | 290 | 190 | 8 |
| Jaber[192] | 2017 | Yemen | 2014 | 2015 | | | | | * | PC | Hosp | 0 | NA | 0 | ·· | 150 | 123 | 7 |
| Masjedi[193] | 2017 | Iran | 2012 | 2014 | | | | | * | PC | Hosp | 0 | 0 | 0 | ·· | 96 | 9 | 8 |
| Heysell[194] | 2016 | Russia | 2014 | 2014 | | | | | * | PC | Hosp | 2 | 100 | ·· | 34 | 62 | 34 | 8 |
| Le[195] | 2016 | South Korea | 2005 | 2012 | | | | | * | RC | Hosp | 3 | NA | 0 | 54 | 92 | 48 | 8 |
| Parchure[196] | 2016 | India | 2004 | 2013 | * | | | | | RC | Hosp | 6 | 100 | 67 | ·· | 549 | 220 | 6 |
| Patel[197] | 2016 | India | 2010 | 2013 | | | | | * | PC | Hosp | 2 | 1.4 | 0 | ·· | 102 | 40 | 8 |
| Dovonou[198] | 2017 | Benin | 2007 | 2011 | | | | | * | RC | Hosp | 0 | 8.71 | 0 | 38 | 179 | 85 | 8 |
| Gebreegziabher[199] | 2016 | Ethiopia | 2013 | 2015 | | | | | * | PC | Hosp | 0 | 11.7 | 0 | ·· | 423 | 283 | 8 |
| Balabanova[200] | 2016 | Multicountry | 2009 | 2012 | * | | | | | PC | Hosp | 1 | 3 | 0 | ·· | 581 | 156 | 8 |
| Mlotshwa[201] | 2016 | South Africa | 2007 | 2013 | | | | * | | RC | Prog | 0 | 22.17 | 0 | 36 | 11641 | 7556 | 7 |
| Snyder[202] | 2016 | Brazil | 2010 | 2010 | | | | | * | RC | Prog | 0 | NA | 18 | 37* | 4284 | 2317 | 7 |
| Ngahane[203] | 2016 | Cameroon | 2007 | 2013 | * | | | | | RC | Hosp | 0 | NA | 19 | 33 | 5110 | 4136 | 5 |
| Ade[204] | 2016 | Benin | 2013 | 2013 | | | | | * | RC | Prog | 0 | 18.2 | 0 | ·· | 2063 | 1066 | 7 |
| Cabrera-Gayta´n[205] | 2016 | Mexico | 2006 | 2014 | | | | | * | RC | Prog | 0 | 4.2 | 0 | 49* | 18415 | 12937 | 7 |
| Yung-Feng Yen[206] | 2016 | Taiwan | 2011 | 2012 | * | | | | | RC | Prog | 0 | 0.7 | 8 | 64.6 | 693 | 326 | 7 |
| Nagai[207] | 2016 | Japan | 2007 | 2015 | * | | | | | RC | Hosp | 0 | 0 | 0 | 72 | 211 | 134 | 8 |
| Asres[208] | 2016 | Ethiopia | 2008 | 2014 | | | | | * | RC | Hosp | 0 | 9.7 | 23 | 30.8 | 447 | 343 | 7 |
| Zenebe[209] | 2016 | Ethiopia | 2011 | 2013 | | | | | * | RC | Hosp | 0 | 16 | 0 | ·· | 238 | 142 | 8 |

| | | | | | | | | | | | | | | | | | | |
|---|---|---|---|---|---|---|---|---|---|---|---|---|---|---|---|---|---|---|
| Gebrezgabiher[210] | 2016 | Ethiopia | 2008 | 2013 | | | | | * | RC | Hosp | 0 | NA | 11.5 | .. | 942 | 595 | 8 |
| Khazaei[211] | 2016 | Iran | 2005 | 2013 | | | | | * | RC | Prog | 0 | 2.7 | 0 | 56.8 | 266 | 244 | 7 |
| Scott[212] | 2017 | USA | 2006 | 2013 | | | * | | | RC | Prog | 0 | 6 | .. | .. | 20477 | 10659 | 7 |
| Gunda[213] | 2016 | Tanzania | 2015 | 2015 | * | | | | | RC | Hosp | 0 | 51.07 | .. | 38* | 361 | 340 | 7 |
| Mekonnen[214] | 2016 | Ethiopia | 2011 | 2014 | | | | | * | RC | Hosp | 0 | 24 | 46.4 | 32.8 | 528 | 421 | 7 |
| Boaz[215] | 2016 | Tanzania | 2012 | 2013 | * | | | | | PC | Hosp | 0 | 100 | 100 | 39 | 19 | 41 | 7 |
| Schnippel[216] | 2015 | South Africa | 2009 | 2011 | * | | | | * | RC | Prog | 1 | 53.2 | 1 | 35* | 9207 | 8406 | 7 |
| Kerkhoff[217] | 2015 | Taiwan | 2002 | 2006 | * | | | | | RC | Hosp | 0 | NA | 0 | 33* | 451 | 1070 | 8 |
| Lettow[218] | 2015 | Malawi | 2013 | 2013 | * | | | | | PC | Hosp | 0 | 55 | .. | 40* | 165 | 183 | 8 |
| Ejeta[219] | 2015 | Ethiopia | 2009 | 2013 | | | | | * | RC | Hosp | 0 | 17.1 | .. | 29.91 | 637 | 537 | 7 |
| Mpagama[220] | 2015 | Tanzania | 2009 | 2013 | | | | | * | RC | Hosp | 0 | 20.4 | .. | 43.4 | 162 | 23 | 8 |
| Pepper[221] | 2015 | South Africa | 2007 | 2009 | * | | | | | RC | Prog | 6 | 24.76 | 21.9 | 33* | 2633 | 1380 | 7 |
| Trajman[222] | 2015 | Brazil | 2012 | 2012 | | | | | * | RC | Prog | 0 | 9.8 | 0 | .. | 1190 | 666 | 7 |
| Wejse[223] | 2015 | Guinea-Bissau | 2003 | 2013 | * | | | | | PC | Prog | 6 | 28.9 | 0 | .. | 813 | 499 | 7 |
| Nglazi[224] | 2015 | South Africa | 2009 | 2011 | * | | | | | RC | Prog | 0 | 55 | 24.8 | 33* | 389 | 408 | 7 |
| a-Villaordun˜ a[225] | 2015 | Uganda | 1997 | 2003 | * | | | | | RC | Prog | 6 | 38.5 | .. | 39.2 | 195 | 89 | 7 |
| Jung[226] | 2016 | South Korea | 2005 | 2015 | | | | | * | RC | Hosp | 0 | 0 | 100 | .. | 40 | 47 | 8 |
| Bastos[227] | 2016 | Portugul | 2007 | 2013 | * | | | | | RC | Hosp | 6 | 19 | 16.6 | 47* | 501 | 180 | 8 |
| García-Basteiro[228] | 2016 | Mozambique | 2011 | 2012 | * | | | | | RC | Prog | 0 | 71.79 | 16.6 | 36.4 | 1096 | 856 | 7 |
| Dale[229] | 2016 | Australia | 2002 | 2013 | * | * | | | | RC | Prog | 6 | 1.1 | 60 | .. | 2125 | 1831 | 7 |
| Shuldiner[230] | 2016 | Isreal | 2000 | 2010 | * | | | | | RC | Prog | 6 | 4.1 | 18 | .. | 1551 | 1265 | 7 |
| Ba´ez-Saldaña[231] | 2016 | Mexico | 1995 | 2010 | * | * | | | * | PC | Hosp | 6 | 2.4 | 0 | 46 | 462 | 342 | 8 |
| Kaplan[232] | 2016 | South Africa | 2010 | 2014 | | | | | * | RC | Hosp | 0 | 100 | .. | .. | 12781 | 10429 | 7 |
| Gebremariam[233] | 2016 | Ethiopia | 2008 | 2014 | * | | | | | RC | Hosp | 0 | 10 | 26.9 | 28.5 | 886 | 763 | 8 |

| | | | | | | | | | | | | | | | | | | |
|---|---|---|---|---|---|---|---|---|---|---|---|---|---|---|---|---|---|---|
| Lee[234] | 2016 | South Korea | 2012 | 2013 | * | | | | | RC | Hosp | 6 | 0.5 | 18 | 61 | 141 | 53 | 8 |
| Ranzani[235] | 2016 | Brazil | 2009 | 2013 | | | | | * | RC | Prog | 0 | 8.7 | 0 | ·· | 44572 | 17245 | 7 |
| Mhimbira[236] | 2016 | Tanzania | 2010 | 2013 | * | | | | * | RC | Prog | 0 | 39.9 | 17.7 | 35* | 2943 | 1892 | 7 |
| Mohammadzadeh[237] | 2016 | Iran | 2012 | 2013 | | | | | * | RC | Hosp | 0 | NA | 0 | ·· | 79 | 88 | 8 |
| Yang[238] | 2016 | South Korea | 1989 | 2014 | * | | | | | RC | Hosp | 0 | 0 | 0 | 62 | 79 | 45 | 8 |
| Berhanu[239] | 2016 | South Africa | 2013 | 2014 | * | | | | | PC | Hosp | 2 | 83.2 | 24.8 | 36* | 104 | 110 | 8 |
| Feltrin[240] | 2016 | Brazil | 1996 | 2004 | * | | | | | RC | Prog | 0 | NA | 20.5 | ·· | 3169 | 1278 | 7 |
| Junior[241] | 2016 | Brazil | 2012 | 2012 | | | | | * | RC | Prog | 6 | NA | ·· | ·· | 2559 | 1495 | 7 |
| Behnaz[242] | 2015 | Iran | 2007 | 2012 | | | | * | * | RC | Hosp | 0 | 1.9 | 0 | 52.93 | 114 | 97 | |
| Milanov[243] | 2015 | Bulgaria | 2009 | 2010 | | | | | * | RC | Prog | 1 | 0 | ·· | 42.8 | 35 | 15 | 7 |
| Shahrezaei[244] | 2015 | Iran | 2006 | 2011 | * | | | | | RC | Prog | 0 | 0.3 | 32 | ·· | 737 | 655 | 7 |
| Bigna[245] | 2015 | Cameroon | 2006 | 2013 | * | | | | | RC | Hosp | 0 | 100 | 39 | 39.5 | 52 | 47 | 7 |
| Yu-Shiuan Lin[246] | 2015 | Taiwan | 2006 | 2011 | * | | | | | RC | Prog | 6 | NA | 0 | 79.9 | 1968 | 608 | 7 |
| Djouma[247] | 2015 | Cameroon | 2013 | 2013 | | | | * | | RC | Prog | 0 | 22.1 | 0 | 36.7 | 269 | 356 | 7 |
| N. Kwak[248] | 2015 | South Korea | 2006 | 2010 | | | | | * | RC | Hosp | 1 | NA | 20.3 | 37* | 69 | 54 | 8 |
| Calligaro[249] | 2015 | South Africa | 2010 | 2013 | * | | | | | PC | Hosp | 6 | NA | ·· | ·· | 197 | 144 | 7 |
| Igari[250] | 2015 | Japan | 2007 | 2012 | * | | | | | RC | Hosp | 0 | 0.26 | 0 | 62.1 | 548 | 211 | 7 |
| Meressa[251] | 2015 | Ethiopia | 2009 | 2014 | | | | | * | PC | Hosp | 2 | 21.7 | 7.0 | 27* | 325 | 287 | 8 |
| Hang[252] | 2015 | Japan | 2007 | 2009 | | | | | * | PC | Hosp | 6 | 5.1 | 0 | ·· | 341 | 89 | 7 |
| Ukwaja[253] | 2015 | Nigeria | NA | NA | | | | | * | RC | Hosp | 0 | 16.29 | 0 | ·· | 660 | 368 | 8 |
| ZHANG[254] | 2015 | China | 2011 | 2014 | | | | | * | PC | Hosp | 2 | NA | 0 | 47.4 | 119 | 41 | 7 |
| Huyen[255] | 2016 | Vietnam | 2011 | 2013 | | | | | * | RC | Prog | 0 | 100 | 29.1 | ·· | 900 | 210 | 7 |
| Park[256] | 2016 | South Korea | 2005 | 2010 | | | * | | | RC | Hosp | 2 | NA | 0 | 41.7 | 164 | 54 | 7 |
| Nabukenya-Mudiope[257] | 2015 | Uganda | 2010 | 2010 | | | | | * | RC | Prog | 0 | 58.1 | 5.6 | ·· | 523 | 207 | 7 |

| | | | | | | | | | | | | | | | | | | |
|---|---|---|---|---|---|---|---|---|---|---|---|---|---|---|---|---|---|---|
| Umanah[258] | 2015 | South Africa | 2007 | 2010 | * | | | | | RC | Hosp | 2 | 100 | 20.7 | ·· | 457 | 490 | 8 |
| Chien[259] | 2014 | Taiwan | 2004 | 2011 | | | | | * | RC | Hosp | 3 | NA | 0 | 64* | 299 | 96 | 8 |
| Yamana[260] | 2015 | Japan | 2010 | 2013 | * | | | | | RC | Prog | 0 | 0.1 | 0 | 74.5 | 566 | 311 | 7 |
| Dobler[261] | 2015 | Mongolia | 2010 | 2011 | | | | | * | RC | Prog | 0 | NA | 0 | ·· | 974 | 794 | 7 |
| Sawadogo[262] | 2015 | Burkina Faso | 2009 | 2009 | | | | | * | CC | Prog | 0 | NA | 0 | 39.5* | 129 | 71 | 7 |
| Oshi[263] | 2014 | Nigeria | 2011 | 2012 | | | | | * | RC | Prog | 0 | 20.50 | 5.6 | ·· | 963 | 705 | 7 |
| Chiang[264] | 2015 | Taiwan | 2005 | 2010 | | | | | * | RC | Hosp | 0 | 0.3 | 0 | ·· | 1094 | 379 | 8 |
| Kosgei[265] | 2015 | Kenya | 2013 | 2013 | | | | | * | RC | Prog | 0 | 31.12 | 0 | ·· | 9947 | 6109 | 7 |
| Yilmaz[266] | 2015 | Turkey | 2005 | 2011 | | * | | | | RC | Prog | 6 | NA | 39.4 | 32.15 | 1339 | 1111 | 7 |
| Sengul[267] | 2015 | Turkey | 2005 | 2011 | | | | | * | RC | Hosp | 6 | NA | 0 | ·· | 480 | 258 | 8 |
| Delgado-Sánchez[268] | 2015 | Mexico | 2000 | 2012 | | | | | * | RC | Prog | 6 | NA | ·· | 46* | 115189 | 66189 | 7 |
| Sun[269] | 2015 | China | 2001 | 2002 | * | | | | | RC | Prog | 6 | 0 | ·· | 49* | 171 | 63 | 7 |
| Wu[270] | 2015 | Taiwan | 2006 | 2008 | | * | | | | RC | Prog | 0 | NA | ·· | 49* | 4165 | 1419 | 7 |
| Balkema[271] | 2014 | South Africa | 2012 | 2013 | * | | | | | PC | Hosp | 0 | 53 | 44.6 | 36.5 | 38 | 45 | 8 |
| Lanoix[272] | 2014 | France | 2000 | 2009 | * | | | | | RC | Hosp | 0 | 41 | ·· | 47.4 | 77 | 20 | 8 |
| Dangisso[273] | 2014 | Uganda | 2003 | 2012 | | | | | * | RC | Prog | 6 | 66.67 | 17.44 | 29 | 20193 | 16867 | 7 |
| Marais[274] | 2014 | South Africa | 2004 | 2007 | * | | | | | RC | Prog | 2 | 62.7 | ·· | ·· | 170 | 154 | 7 |
| Lucenko[275] | 2014 | Latvia | 2006 | 2010 | | | | | * | RC | Prog | 0 | 7 | 6 | 42* | 1704 | 772 | 7 |
| Jain[276] | 2013 | India | 2009 | 2009 | | | | | * | PC | Prog | 2 | NA | ·· | 31.42 | 81 | 49 | 7 |
| Atif[277] | 2014 | Malaysia | 2010 | 2011 | * | | | | * | RC | Hosp | 0 | NA | 0 | 49.11 | 236 | 100 | 8 |
| Pusch[278] | 2014 | USA | 2000 | 2005 | * | | | | | RC | Prog | 0 | 21 | 100 | 49 | 224 | 214 | 7 |
| Aung[279] | 2014 | Bangladesh | 2005 | 2011 | | | | | * | PC | Prog | 2 | NA | ·· | ·· | 364 | 151 | 7 |
| Han[280] | 2013 | Multicountry | NA | NA | * | | | | | RC | Prog | 0 | 100 | 23 | 34* | 609 | 159 | 7 |
| Oshi[281] | 2014 | Nigeria | 2011 | 2012 | | | | | * | RC | Prog | 0 | 22.5 | 5.6 | ·· | 963 | 705 | 7 |

| | | | | | | | | | | | | | | | | | | |
|---|---|---|---|---|---|---|---|---|---|---|---|---|---|---|---|---|---|---|
| Peltzer[282] | 2014 | South Africa | NA | NA | | | | | * | PC | Prog | 0 | 54.3 | ·· | ·· | 873 | 302 | 7 |
| Choi[283] | 2014 | South Korea | 2005 | 2012 | | | | | * | PC | Hosp | 0 | NA | 0 | 44 | 563 | 106 | 7 |
| Khaliaukin[284] | 2014 | Belarus | 2009 | 2010 | | | | | * | RC | Prog | 2 | 15 | ·· | 45* | 367 | 72 | 7 |
| Kuksa[285] | 2014 | Latvia | 2000 | 2010 | | | | | * | RC | Prog | 4 | 12 | 7 | 39 | 1333 | 446 | 7 |
| Shuldiner[286] | 2014 | Isreal | 2000 | 2010 | | * | | | | RC | Prog | 6 | 8.9 | 18 | ·· | 2612 | 1925 | 7 |
| Pietersen[287] | 2014 | South Africa | 2008 | 2012 | * | | * | | | RC | Prog | 4 | 42.72 | ·· | 33* | 58 | 49 | 7 |
| Babalik[288] | 2014 | Turkey | 2006 | 2010 | | | | | * | RC | Prog | 6 | NA | ·· | ·· | 14402 | 9443 | 7 |
| Uyei[289] | 2014 | South Africa | 2007 | 2008 | * | | | | * | RC | Prog | 0 | 100 | ·· | ·· | 588 | 749 | 7 |
| Velásquez[290] | 2014 | Russia | 2000 | 2004 | | | | | * | RC | Prog | 6 | 0.8 | 8.5 | 35.9 | 531 | 107 | 7 |
| Baghaei[291] | 2014 | Iran | 2004 | 2011 | * | | | | | RC | Hosp | 0 | 1.2 | ·· | 65.92 | 96 | 124 | 8 |
| Djibuti[292] | 2014 | Georgia | 2004 | 2004 | | | | | * | PC | Hosp | 0 | NA | ·· | 35* | 149 | 44 | 8 |
| Bastard[293] | 2015 | Armenia | 2008 | 2010 | | | | | * | RC | Prog | 6 | NA | ·· | 38* | 338 | 65 | 7 |
| Hamusse[294] | 2014 | Ethiopia | 1997 | 2011 | | | | | * | RC | Prog | 0 | 7.9 | 0 | ·· | 7734 | 6487 | 7 |
| Wingfield[295] | 2014 | Peru | 2002 | 2009 | | | | | * | PC | Prog | 6 | NA | ·· | 31 | 517 | 359 | 7 |
| Putri[296] | 2014 | Indonesia | 2009 | 2011 | | | * | | | RC | Hosp | 1 | 0 | 0 | 37 | 115 | 98 | 7 |
| Esmael[297] | 2014 | Ethiopia | 2008 | 2013 | | | | | * | RC | Hosp | 0 | 44.76 | 25.2 | 30.6 | 371 | 346 | 8 |
| Alobu[298] | 2014 | Nigeria | 2011 | 2012 | * | | | | | RC | Prog | 0 | 79.5 | 5.6 | ·· | 963 | 705 | 7 |
| Kwon[299] | 2014 | South Korea | 2009 | 2010 | * | | | | | RC | Hosp | 0 | 0 | 0 | 50* | 1410 | 1071 | 7 |
| Alobu[300] | 2014 | Nigeria | 2011 | 2012 | | | | | * | CC | Hosp | 0 | 13.6 | 0 | ·· | 602 | 383 | 7 |
| Pefura-Yone[301] | 2014 | Cameroon | 2009 | 2012 | | | * | | | PC | Hosp | 6 | 27.9 | 0 | 33* | 126 | 46 | 8 |
| Ershova[302] | 2014 | USA | 1993 | 2008 | * | | | | | RC | Prog | 6 | NA | ·· | ·· | 19048 | 12178 | 7 |
| Thomas Iype[303] | 2014 | India | 2010 | 2011 | * | | | | | PC | Hosp | 6 | NA | 100 | ·· | 22 | 21 | 7 |
| Viswanathan[304] | 2014 | India | 2011 | 2012 | | | | * | | PC | Hosp | 6 | NA | 12.9 | ·· | 159 | 50 | 7 |
| Alo[305] | 2014 | Fiji | 2010 | 2012 | | | | | * | RC | Hosp | 6 | NA | 16.96 | ·· | 224 | 168 | 8 |

| | | | | | | | | | | | | | | | | | | |
|---|---|---|---|---|---|---|---|---|---|---|---|---|---|---|---|---|---|---|
| Przybylski[306] | 2014 | Poland | 2001 | 2010 | | | | | * | RC | Hosp | 6 | 0.3 | 6.7 | 51.5 | 1343 | 662 | 8 |
| O'Donnell[307] | 2013 | South Africa | 2006 | 2007 | | | * | | | RC | Prog | 4 | 71.9 | 9.7 | 35* | 49 | 65 | 7 |
| Getahun[308] | 2013 | Ethiopia | 2004 | 2009 | * | | | * | * | RC | Hosp | 6 | NA | 40.5 | 30.1 | 3017 | 3433 | 8 |
| Anderson[309] | 2013 | UK | 2004 | 2012 | | | | | * | PC | Hosp | 2 | 12.56 | 29.9 | ·· | 103 | 101 | 8 |
| Manda[310] | 2013 | South Africa | 2000 | 2004 | * | | | | | RC | Prog | 2 | 39.2 | ·· | ·· | 1290 | 786 | 7 |
| Babalik[311] | 2012 | Turkey | 2006 | 2009 | * | | | * | * | RC | Prog | 6 | NA | 0 | ·· | 8019 | 3167 | 7 |
| Patra[312] | 2013 | India | 2005 | 2010 | | | | | * | RC | Hosp | 6 | 0.2 | 29.2 | ·· | 1343 | 1093 | 8 |
| Horita[313] | 2013 | Japan | 2008 | 2011 | * | | | | | RC | Hosp | 6 | 0 | 0 | 64.9 | 297 | 135 | 8 |
| Blöndal[314] | 2013 | Estonia | 2002 | 2011 | * | | | | * | RC | Prog | 6 | 5.9 | ·· | ·· | 1777 | 674 | 7 |
| Schwartz[315] | 2013 | Botswana | 2005 | 2010 | * | | | | | RC | Hosp | 6 | 100 | 0 | 35.5* | 187 | 152 | 8 |
| Abouzeid[316] | 2013 | South Africa | 2001 | 2010 | * | | | | | RC | Prog | 6 | 2.8 | ·· | ·· | 19738 | 13929 | 7 |
| Kabali[317] | 2013 | Tanzania | 2001 | 2008 | * | | | | | PC | Hosp | 6 | NA | ·· | ·· | 92 | 287 | 8 |
| Deepa[318] | 2013 | India | 2011 | 2011 | | | | | * | RC | Prog | 6 | 5.8 | 0 | ·· | 1440 | 507 | 7 |
| Kang[319] | 2013 | South Korea | 2000 | 2002 | * | | | | * | RC | Hosp | 2 | 1.5 | 3.8 | ·· | 1039 | 368 | 8 |
| Sulaiman[320] | 2013 | Malaysia | 2006 | 2007 | | | | | * | RC | Hosp | 6 | 0 | 17.6 | ·· | 916 | 351 | 8 |
| Reed[321] | 2013 | South Korea | NA | NA | * | * | | | | PC | Hosp | 6 | NA | 0 | ·· | 551 | 106 | 8 |
| Nakanwagi-Mukwaya[322] | 2013 | Uganda | 2009 | 2010 | * | | | * | * | RC | Prog | 6 | 64.164 | ·· | 36 | 224 | 107 | 7 |
| Uchimura[323] | 2013 | Japan | 2007 | 2010 | * | | | * | * | RC | Prog | 6 | 0.2 | 21.4 | ·· | 61107 | 35582 | 7 |
| Hoa[324] | 2012 | Vietnam | 2008 | 2008 | | | | | * | RC | Prog | 6 | NA | 13.6 | ·· | 1721 | 792 | 7 |
| Ugarte-Gil[325] | 2013 | Peru | 2010 | 2011 | | | | | * | PC | Hosp | 6 | NA | 0 | 28* | 160 | 131 | 8 |
| Mitnick[326] | 2013 | Peru | 1999 | 2002 | * | | | | | RC | Prog | 2 | 1.5 | 9 | 31.4 | 408 | 261 | 7 |
| Gler[327] | 2013 | Phillipines | 2005 | 2008 | | | | | * | PC | Hosp | 2 | NA | ·· | ·· | 271 | 168 | 8 |
| Kenangalem[328] | 2013 | Papua New Guinea | 2008 | 2009 | | | * | | | PC | Hosp | 6 | 12.5 | ·· | ·· | 127 | 59 | 8 |
| Macedo[329] | 2013 | Brazil | 2007 | 2011 | | * | | | | RC | Prog | 6 | 22.1 | 8.96 | ·· | 13489 | 1385 | 7 |

| | | | | | | | | | | | | | | | | | | |
|---|---|---|---|---|---|---|---|---|---|---|---|---|---|---|---|---|---|---|
| Antoine[330] | 2013 | France | 2009 | 2009 | | | | | * | RC | Prog | 6 | NA | 0 | ·· | 1232 | 778 | 7 |
| Tang[331] | 2013 | China | 2006 | 2011 | | | | | * | RC | Hosp | 1 | 0 | ·· | ·· | 395 | 191 | 8 |
| Ananthakrishnan[332] | 2013 | India | 2011 | 2011 | * | | | | * | RC | Prog | 6 | 2 | 24.49 | ·· | 1193 | 292 | 7 |
| Pazarli[333] | 2013 | Turkey | 2000 | 2005 | | | | | * | RC | Hosp | 2 | 0 | 0 | 40.5 | 81 | 22 | 8 |
| Tweya[334] | 2013 | Malawi | 2008 | 2010 | * | | | * | * | RC | Hosp | 6 | 56 | 0 | 31 | 1460 | 901 | 8 |
| Yen[335] | 2013 | Taiwan | 2006 | 2010 | * | * | | | | RC | Prog | 6 | NA | 0 | 64.2 | 2454 | 1033 | 7 |
| Mpagama[336] | 2013 | Tanzania | 2009 | 2011 | * | | | | | RC | Hosp | 2 | 15 | 0 | 36* | 39 | 19 | 8 |
| Ismail[337] | 2013 | Malaysia | 2010 | 2010 | | | | | * | PC | Prog | 6 | 100 | ·· | ·· | 194 | 25 | 7 |
| Wang[338] | 2013 | China | 2004 | 2008 | * | * | | | | RC | Prog | 6 | NA | 0 | 54.5 | 601 | 107 | 7 |
| Shaweno[339] | 2012 | Ethiopia | 2006 | 2010 | * | | | | | RC | Hosp | 6 | 100 | 18.1 | 30* | 417 | 323 | 8 |
| Limmahakhun[340] | 2012 | Thailand | 2000 | 2009 | * | | | | | RC | Hosp | 6 | 100 | 57.9 | 38.8 | 100 | 69 | 8 |
| Takarinda[341] | 2012 | Zimbabwe | 2009 | 2009 | * | | | | * | RC | Prog | 6 | 84.9 | ·· | ·· | 135 | 90 | 7 |
| Hoa[342] | 2012 | Multicountry | 2003 | 2005 | * | | | * | * | RC | Prog | 6 | NA | 6.3 | ·· | 22833 | 10469 | 7 |
| Cavanaugh[343] | 2012 | Russia | 2002 | 2005 | | | | | * | RC | Prog | 2 | 0 | ·· | 42 | 165 | 35 | 7 |
| Bloss[344] | 2012 | Taiwan | 2007 | 2008 | | | | | * | PC | Prog | 0 | NA | 0 | ·· | 8248 | 3280 | 7 |
| Kassa[345] | 2012 | Ethiopia | 2005 | 2009 | * | | | | | RC | Hosp | 6 | 100 | 30.7 | 35* | 1737 | 2473 | 8 |
| Girardi[346] | 2012 | Italy | NA | NA | * | | | | * | PC | Prog | 6 | 100 | 19.91 | ·· | 199 | 47 | 7 |
| Palacios[347] | 2012 | Peru | 1996 | 2005 | * | | | | | RC | Prog | 2 | 100 | 48.1 | 30.5* | 40 | 12 | 7 |
| Feng[348] | 2012 | Taiwan | 2007 | 2009 | * | | * | * | | PC | Hosp | 6 | 3.1 | 0 | 64.7 | 819 | 240 | 8 |
| Mukherjee[349] | 2012 | India | 1999 | 2005 | * | | | * | * | RC | Prog | 6 | NA | 9.29 | ·· | 2498 | 1107 | 7 |
| Chirwa[350] | 2013 | Malawi | 2007 | 2008 | | | | | * | RC | Hosp | 6 | 65.1 | 0 | 36 | 302 | 222 | 8 |
| Mnisi[351] | 2013 | South Africa | 2007 | 2009 | | | | | * | RC | Prog | 6 | 54 | 0 | 33.7 | 198 | 2 | 7 |
| Riou[352] | 2012 | South Africa | 2000 | 2001 | | | * | | | RC | Hosp | 0 | 52 | 0 | ·· | 17 | 3 | 8 |
| Janols[353] | 2012 | Ethiopia | 2007 | 2009 | * | | | * | * | PC | Hosp | 6 | 53.6 | 0 | ·· | 133 | 117 | 8 |

| | | | | | | | | | | | | | | | | | | |
|---|---|---|---|---|---|---|---|---|---|---|---|---|---|---|---|---|---|---|
| Koh[354] | 2012 | South Korea | 2007 | 2009 | | | | | * | RC | Hosp | 1 | NA | 0 | 33* | 26 | 25 | 8 |
| Yee[355] | 2012 | Canada | 1990 | 2000 | | | | | * | RC | Prog | 6 | NA | 17.63 | .. | 184 | 94 | 7 |
| Kattan[356] | 2012 | USA | 2007 | 2009 | * | | | | | RC | Prog | 6 | 5.04 | 48.48 | .. | 163 | 137 | 7 |
| Valade[357] | 2012 | France | 2000 | 2009 | * | | | | | RC | Hosp | 6 | 23 | 28 | 41* | 40 | 13 | 8 |
| Tabarsi[358] | 2012 | Iran | 2004 | 2007 | * | | | | | RC | Hosp | 6 | 100 | 7.2 | 38 | 107 | 4 | 8 |
| Van'tHoog[359] | 2012 | Kenya | 2006 | 2008 | * | | | | * | RC | Prog | 6 | 56 | 20.12 | .. | 5064 | 5811 | 7 |
| Gegia[360] | 2012 | Georgia | 2007 | 2009 | | | | | * | RC | Prog | 3 | NA | 0 | 35* | 711 | 198 | 7 |
| Baghaei[361] | 2012 | Peru | 2000 | 2006 | | | | | * | RC | Hosp | 6 | 1.522 | 16.09 | 31.6 | 255 | 205 | 8 |
| Belo[362] | 2011 | Brazil | 2005 | 2008 | * | | | * | * | PC | Hosp | 6 | NA | .. | 37.7* | 303 | 157 | 8 |
| Srinath[363] | 2011 | India | 2008 | 2008 | | | | | * | RC | Prog | 6 | 5.5 | 16 | .. | 674 | 334 | 7 |
| Takarinda[364] | 2011 | Zimbabwe | 2009 | 2009 | * | | | | * | RC | Prog | 6 | 80 | 12 | 36* | 866 | 934 | 7 |
| Jeon[365] | 2011 | South Korea | 2004 | 2004 | * | | | | * | RC | Hosp | 2 | NA | 6.4 | 44.8 | 156 | 46 | 8 |
| Jonnalagada[366] | 2011 | India | 1994 | 2007 | * | | | | | PC | Prog | 6 | 22 | .. | 36 | 231262 | 255079 | 7 |
| Dooley[367] | 2011 | Morocco | 2007 | 2008 | | | | | * | RC | Prog | 6 | 1 | 0 | 37 | 240 | 106 | 7 |
| Farley[368] | 2011 | South Africa | 2000 | 2004 | * | | | | | PC | Prog | 2 | 37.913 | 2 | 36.5 | 448 | 309 | 7 |
| Bendayan[369] | 2011 | Israel | 2000 | 2005 | * | | | | | RC | Hosp | 2 | 6.1 | .. | 42.7 | 102 | 30 | 8 |
| Christensen[370] | 2011 | Denmark | 1972 | 2008 | * | | | | | RC | Prog | 6 | 0.661 | 100 | 39.5* | 27 | 28 | 7 |
| Visser[371] | 2012 | South Africa | 2005 | 2008 | | | * | | | RC | Hosp | 0 | 10.5 | 0 | 30 | 79 | 34 | 8 |
| Arentz[372] | 2011 | Kenya | 2008 | 2009 | | | | | * | RC | Hosp | 6 | 40.96 | 22 | .. | 104 | 79 | 8 |
| Nahid[373] | 2011 | USA | 1990 | 2001 | * | | | | | RC | Prog | 0 | 36 | 0 | .. | 452 | 113 | 7 |
| Burton[374] | 2011 | Ghana | 2009 | 2009 | * | | | | | RC | Hosp | 6 | 35.4 | 37.2 | 42 | 370 | 229 | 8 |
| Blanc[375] | 2011 | Cambodia | 2006 | 2009 | * | | | | | RC | Hosp | 6 | 100 | .. | 35* | 425 | 236 | 8 |
| Jung[376] | 2010 | USA | 1990 | 2006 | * | | | | | RC | Prog | 6 | 15.1 | .. | .. | 32966 | 20539 | 7 |
| Kim[377] | 2010 | South Korea | 2000 | 2002 | * | | | | | RC | Prog | 2 | 1.5 | 3.8 | 42.9 | 53 | 22 | 7 |

| | | | | | | | | | | | | | | | | |
|---|---|---|---|---|---|---|---|---|---|---|---|---|---|---|---|---|
| Vasankari[378] | 2010 | Finland | 1995 | 1996 | * | | | | * | RC | Prog | 6 | NA | 100 | 70.1* | 94 | 137 | 7 |
| Misra[379] | 2010 | India | 2005 | 2008 | * | | | | | PC | Hosp | 6 | NA | 100 | 30* | 51 | 48 | 8 |
| Wen[380] | 2010 | Taiwan | 1994 | 2007 | | * | | | | PC | Prog | 6 | NA | 0 | ·· | 231262 | 255079 | 7 |
| Hsu[381] | 2010 | Taiwan | 2000 | 2006 | * | | | | | RC | Hosp | 6 | NA | 100 | 54.9 | 71 | 37 | 8 |
| Sassaki[382] | 2010 | Brazil | 2001 | 2004 | | | | | * | RC | Prog | 6 | NA | 0 | ·· | 3150 | 1600 | 7 |
| Ferrer[383] | 2010 | USA | 1994 | 2007 | | | | | * | RC | Prog | 6 | 0 | 0 | ·· | 23 | 12 | 7 |
| Buu[384] | 2010 | Vietnam | 2005 | 2007 | | | | | * | PC | Hosp | 6 | NA | 0 | ·· | 843 | 263 | 8 |
| Silva[385] | 2010 | Brazil | 2005 | 2007 | * | | | | | RC | Hosp | 6 | 68.7 | 64.1 | 43.2 | 30 | 37 | 8 |
| Shaw[386] | 2010 | USA | 2000 | 2005 | * | | | | | RC | Prog | 6 | 19.52 | 100 | ·· | 127 | 83 | 7 |
| Vinnard[387] | 2010 | USA | 1993 | 2005 | * | | | | | RC | Prog | 3 | 40.84 | 100 | ·· | 1150 | 746 | 7 |
| Uwizeye[388] | 2011 | Rwanda | 2006 | 2006 | * | | | * | | RC | Prog | 6 | 45.85 | 27.7 | ·· | 1012 | 661 | 7 |
| Jacobson[389] | 2011 | South Africa | 2000 | 2009 | | | | | * | RC | Hosp | 3 | 21 | 0 | 38.2 | 101 | 25 | 8 |
| Joseph[390] | 2011 | India | 2008 | 2009 | | | | * | | RC | Prog | 6 | NA | 0 | 38.78 | 224 | 62 | 7 |
| Fawibe[391] | 2011 | Nigeria | 1999 | 2008 | * | | | | | RC | Hosp | 6 | 12.1 | 0 | 36 | 45 | 29 | 8 |
| Marais[392] | 2011 | South Africa | 2009 | 2009 | * | | | | | RC | Hosp | 6 | 88.3 | 100 | ·· | 60 | 60 | 8 |
| Horne[393] | 2010 | USA | 1993 | 2005 | * | | | | | RC | Prog | 6 | 4.8 | 23.2 | 45 | 2102 | 1349 | 7 |
| Silva[394] | 2010 | Brazil | 2005 | 2007 | * | | | | | RC | Hosp | 6 | 70.8 | 65.7 | ·· | 202 | 109 | 8 |
| Leimane[395] | 2010 | Latvia | 2000 | 2004 | | | | | * | RC | Prog | 1 | 3.1 | 0 | ·· | 780 | 247 | 7 |
| Dheda[396] | 2010 | South Africa | 2002 | 2008 | * | | * | | | RC | Hosp | 4 | 49 | ·· | 33 | 85 | 89 | 8 |
| Senkoro[397] | 2010 | Tanzania | 2008 | 2008 | | | * | | | PC | Hosp | 6 | 33.7 | 0 | 35.7 | 345 | 153 | 8 |
| Chidambaram[398] | 2020 | Taiwan | 2000 | 2016 | * | | * | * | | RC | Hosp | 0 | 2.3 | 0 | 66.6 | 1975 | 919 | 8 |

***Outcome reported.**

**CC=Case control. PC=Prospective cohort. RC=Retrospective cohort. EPTB=Extrapulmonary Tuberculosis.**
**Hosp= Hospital Data. Prog=Programmatic data. NOS= Newcastle Ottawa Score.**

**Drug resistance,**

**0=Drug sensitive**
**1=Drug resistant (not defined)**
**2=Only MDR**
**3=Only mono-resistant**
**4=Only XDR**
**6=Drug sensitivity characteristics not available**

**Supplementary Table 3: Parameters adjusted for with respect to the outcomes in the individual studies.**

| Study | Age | HIV | Type | BMI | Smoking | Education | Diabetes | Alcohol | Race | SES | Cavity | Hb | Hypertension | Bacteria |
|---|---|---|---|---|---|---|---|---|---|---|---|---|---|---|
| Shimazaki[105] | * | | | * | * | | | * | | | | * | | * |
| Chidambaram[398] | * | * | | * | * | | * | * | | | * | | * | |
| Nagu[146] | * | * | | * | * | * | | | | | * | | * | |
| Ko[157] | * | * | * | | | | | | | | | | * | |
| Umanah[258] | * | | * | * | | | | | | | * | * | | |
| Ferreira[85] | * | * | * | | * | | | * | | | | * | | |
| Bigna[245] | * | | * | | | * | | | | | | * | | |
| Gunda[213] | * | * | | | * | | | | | | | * | | |
| Han[96] | * | | | * | | | | | | | | * | | |
| Balabanova[200] | * | * | * | | * | | | * | | * | * | | | |
| Atif[277] | * | | | | * | | * | * | | | * | | | |
| Kang[319] | * | | | * | | | * | | | | * | | | |
| Nahid[373] | * | * | | | | | | | | | * | | | |
| Shaw[386] | * | * | | | | | * | | * | * | | | | |
| Aibana[135] | * | * | | | | | | * | | * | | | | |
| Pradipta[65] | * | | | | | | * | | | * | | | | |
| Girardi[346] | * | * | * | | | * | | | | * | | | | |
| Nagu[145] | * | * | | | | | | | | * | | | | |
| Uyei[289] | * | | | | | | | | | * | | | | |
| Sun[269] | * | | | | | | | | | * | | | | |
| Pedrazzoli[63] | * | | * | | | | | | * | | | | | |
| Jung[376] | * | | | | | | | | * | | | | | |

| | | | | | | | | | |
|---|---|---|---|---|---|---|---|---|---|
| Christensen[370] | * | | | | | | | | * |
| Rossetto[71] | * | | | | | | | | * |
| Arroyo[40] | * | * | | | * | * | * | * | |
| Reed[321] | * | | | | * | * | * | * | |
| Jonnalagada[366] | * | | | * | | * | * | * | |
| Washington[11] | | * | * | | | * | * | * | |
| Wakamatsu[133] | * | | | | * | | * | * | |
| Ba´ez-Saldaña[231] | * | * | | | | | * | * | |
| Georghiou[182] | * | * | | * | * | | * | | |
| Yamana[260] | * | | | * | * | | * | | |
| Seifert[183] | * | * | | * | | | * | | |
| Mehta[179] | * | | | | | | * | | |
| Nagu[155] | * | * | | * | * | * | | | |
| Mattila[141] | * | | | * | * | * | | | |
| Yung-Feng Yen[206] | * | | | * | | * | | | |
| Stosic[3] | * | | * | | | * | | | |
| Kwon[299] | * | | | | * | | | | |
| Frank[92] | * | | | | * | | | | |
| Min[54] | * | | | | * | | | | |
| Mitnick[326] | * | * | * | * | | | | | |
| Humphrey[23] | * | | * | * | | | | | |
| Zurcher[51] | * | | * | * | | | | | |
| Gonah[34] | * | * | | * | | | | | |
| Wejse[223] | * | * | | * | | | | | |

| | | | |
|---|---|---|---|
| Shaweno[339] | * | * | * |
| Ershova[302] | * | * | * |
| Han[280] | * | * | * |
| Pepper[221] | * | * | * |
| Alobu[298] | * | * | * |
| Pusch[278] | * | * | * |
| Mhimbira[236] | * | * | * |
| Gebremariam[233] | * | * | * |
| García-Basteiro[228] | * | * | * |
| Nglazi[224] | * | * | * |
| Lakoh[29] | * | * | * |
| Ramos[16] | | * | * |
| Hoa[342] | * | | * |
| Mok[118] | * | | * |
| Kaplan[90] | * | | * |
| Balaky[60] | * | | * |
| Musaazi[59] | * | | * |
| Crabtree-Ram´ırez[95] | * | | * |
| Janols[353] | * | * | |
| Manda[310] | * | * | |
| Shuldiner[230] | * | * | |
| Shin[184] | * | * | |
| Evans[88] | * | * | |
| Ohene[72] | * | * | |

| | | |
|---|---|---|
| Schnippel[216] | * | * |
| Ngahane[203] | * | * |
| Zürcher[49] | * | * |
| Bhering[43] | | * |
| Vasankari[378] | * | |
| Kassa[345] | * | |
| Babalik[311] | * | |
| Somsong[66] | * | |
| Makhmudova[36] | * | |
| Marais[274] | * | |
| Igari[250] | * | |
| Lettow[218] | * | |
| Teklu[187] | * | |

*Parameter adjusted for in the study

HIV= Human Immunodeficiency Virus. BMI= Body Mass Index. SES=Socio-economic status. Hb= Hemoglobin.

**Supplementary Table 4: Meta-regression analysis to assess the impact of the difference in Default, loss to follow up, treatment completion and HIV status on the association of male sex with all-cause mortality.**

| Characteristic | Increase in log-odds in males compared to females | Number of studies | Difference in log-odds between males and females | p-value |
|---|---|---|---|---|
| **Default** | 1 | 21 | 0.16[-0.21 to 0.55] | 0.384 |
| **Loss to follow up** | 1 | 17 | 0.12[-0.32 to 0.56] | 0.588 |
| **Treatment completion** | 1 | 18 | 0.05[-0.30 to 0.41] | 0.781 |
| **HIV** | 1 | 15 | 0.39[0.14 to 0.66] | 0.002 |

HIV = Human Immuno-deficiency virus.

**Supplementary Table 5: Association of male sex with treatment success in patients with Tuberculosis**

| Outcome | Estimate | No. of studies | Unadjusted Effect size | $I^2$ | No. of studies | Adjusted Effect size | $I^2$ |
|---|---|---|---|---|---|---|---|
| **Success** | OR | 130 | 0.74[0.70-0.80] | 90.54 | 40 | 0.87[0.81-0.92] | 45.2 |
| | HR | 35 | 0.72[0.62-0.82] | 75.25 | 20 | 0.82[0.73-0.92] | 25.6 |

HR=Hazard ratio. OR=Odds ratio.

**Supplementary Table 6: Meta-regression analysis to assess the impact of comorbidities of the study participants on the association of male sex with tuberculosis treatment outcomes.**

| Characteristics in the study population | Increase in the variable | All-cause mortality | | | Treatment Success | | | Sputum Smear positivity | | | Sputum Culture positivity | | |
|---|---|---|---|---|---|---|---|---|---|---|---|---|---|
| | | No. of studies | Change in log odds | p-value | No. of studies | Change in log odds | p-value | No. of studies | Change in log odds | p-value | No. of studies | Change in log odds | p-value |
| **Age** | 10 years | 75 | 0.03[-0.02 to 0.09] | 0.299 | 48 | -0.05[-0.16 to 0.05] | 0.295 | 20 | 0.14[0 to 0.29] | 0.051 | 10 | 0.14[0 to 0.29] | 0.05 |
| **Diabetes** | 10% | 40 | 0[-0.012 to 0.012] | 0.995 | 37 | 0.03[-0.11 to 0.16] | 0.680 | 9 | - | - | 7 | - | - |
| **Hypertension** | 10% | 10 | 0.06[-0.14 to 0.26] | 0.533 | 5 | -0.05[-0.23 to 0.13] | 0.571 | 3 | - | - | - | - | - |
| **Alcohol** | 10% | 28 | 0[-0.09 to 0.09] | 0.865 | 18 | -0.03[-0.15 to 0.08] | 0.587 | 6 | - | - | 4 | - | - |
| **Smoking** | 10% | 26 | 0.05[0 to 0.11] | 0.050 | 26 | -0.07[-0.13 to 0.11] | 0.897 | 5 | - | - | 6 | - | - |
| **Cardiovascular diseases** | 10% | 10 | -0.26[-0.68 to 0.16] | 0.217 | 4 | -0.16[-0.37 to 0.05] | 0.133 | 2 | - | - | 2 | - | - |
| **COPD** | 10% | 12 | -0.06[-0.17 to 0.07] | 0.444 | 7 | -0.05[-0.94 to 0.85] | 0.912 | 3 | - | - | 4 | - | - |
| **HIV** | 10% | 108 | -0.02[-0.05 to -0.01] | 0.003 | 89 | 0.01[-0.02 to 0.03] | 0.679 | 20 | -0.01[-0.05 to 0.05] | 0.873 | 9 | - | - |

COPD=Chronic obstructive Pulmonary disease. HIV=Human Immunodeficiency Virus.

### Section-IV

#### Supplementary figures

#### (Forest charts, funnel plots and bubble plots)

**eFigure 1.** Pooled odds ratio for all-cause mortality in male patients compared to female patients. (Unadjusted)

1a) Forest plot

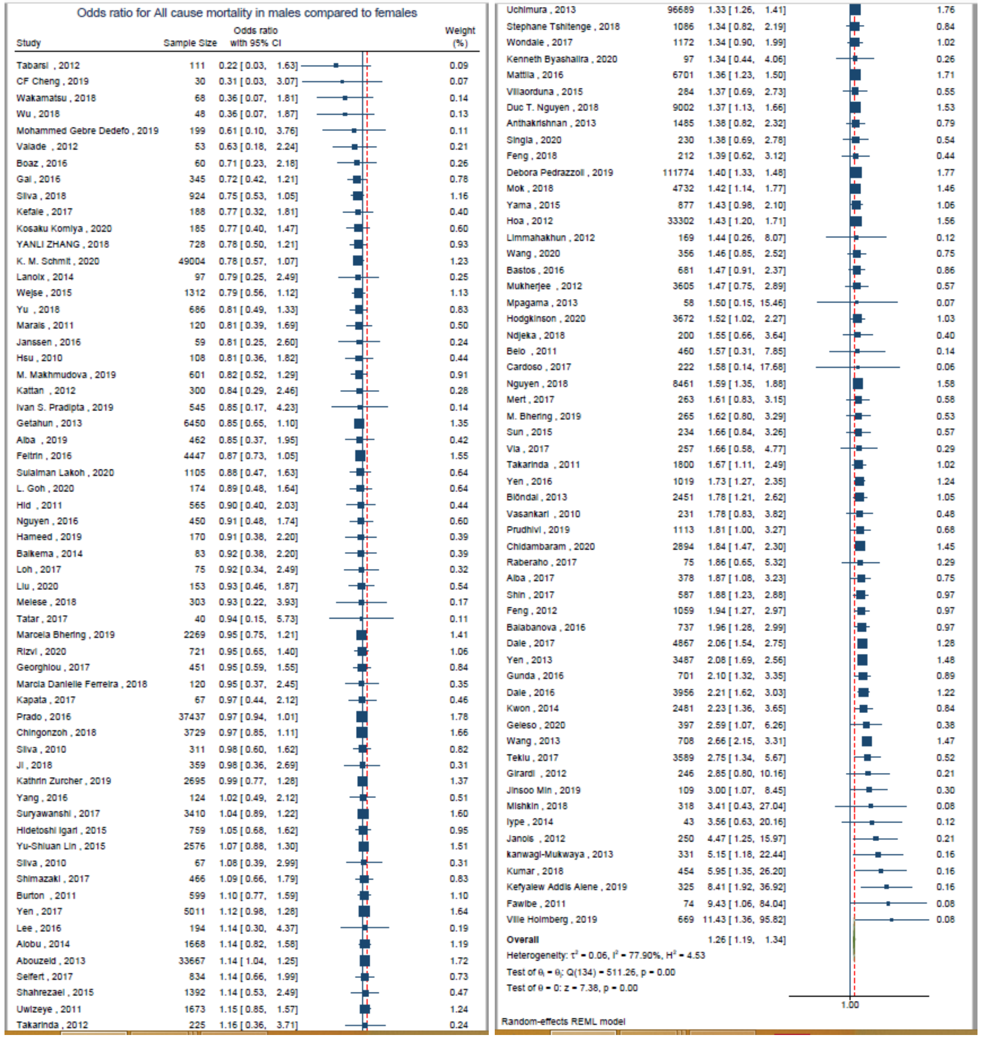

1b) Subgroup analysis based on the time of outcome assessment. (During Treatment and During Follow up)

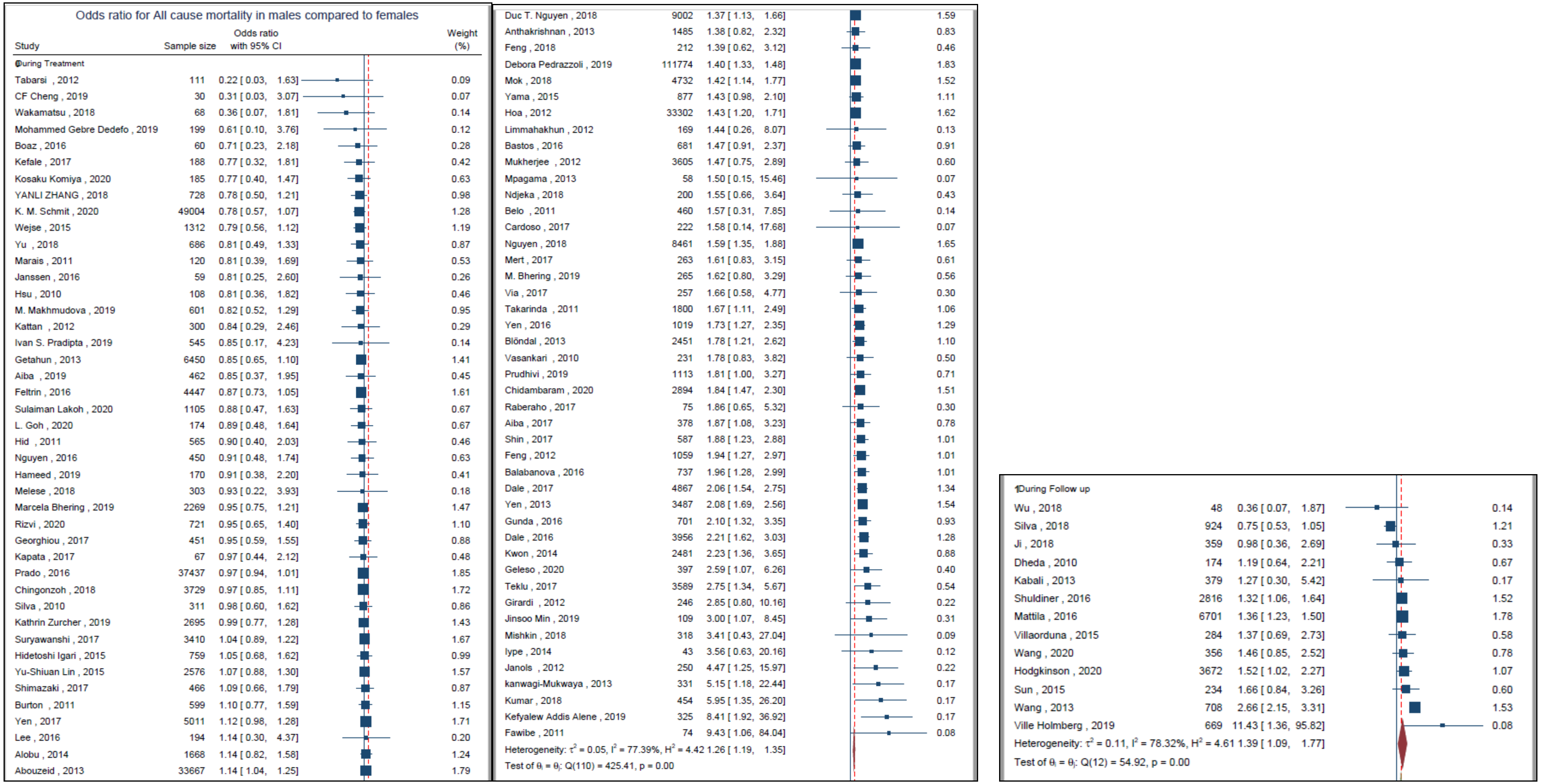

1c) Subgroup analysis based on the World Bank income status classification.

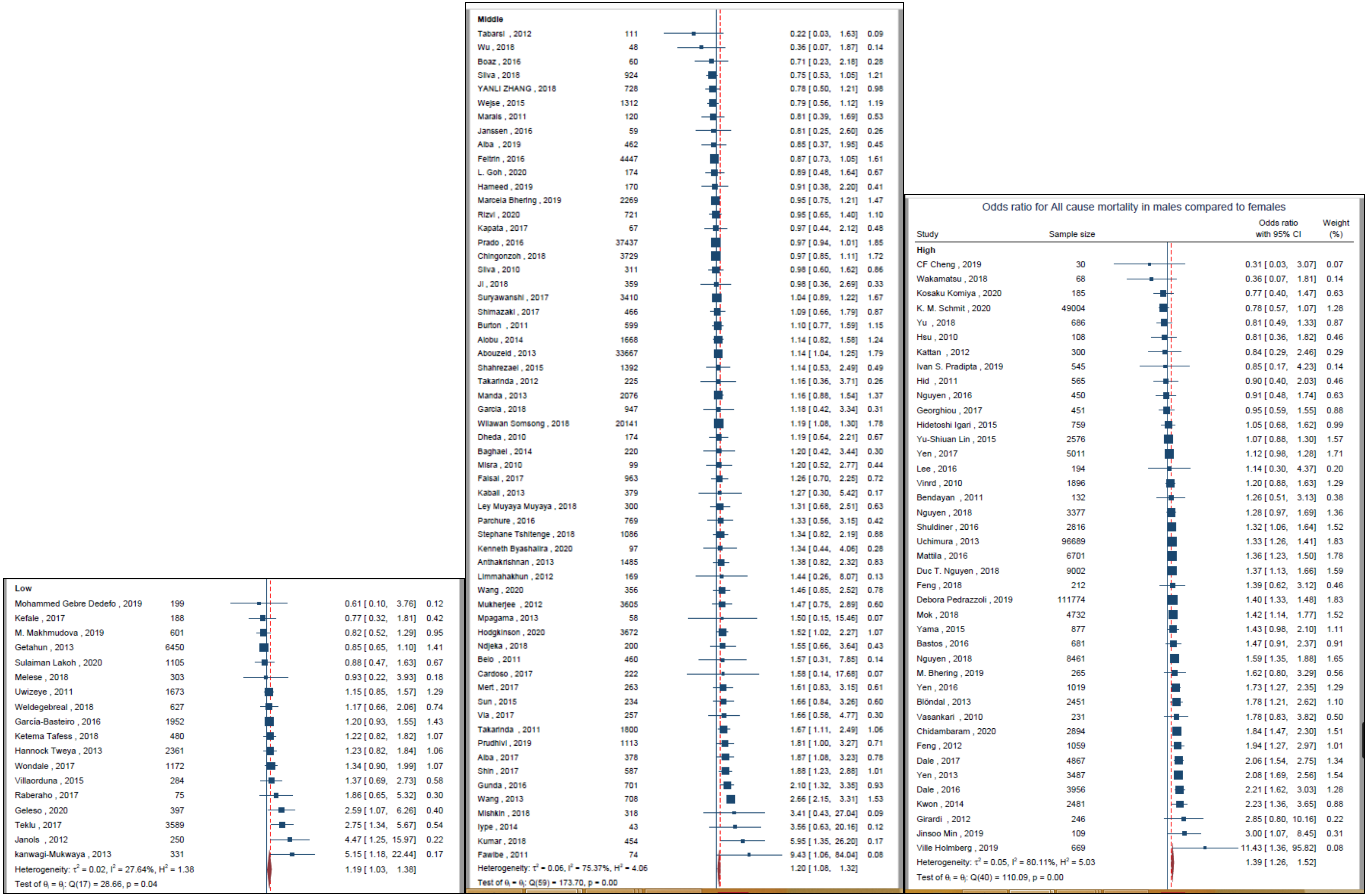

1d) Subgroup analysis based on the TB incidence of the study country.

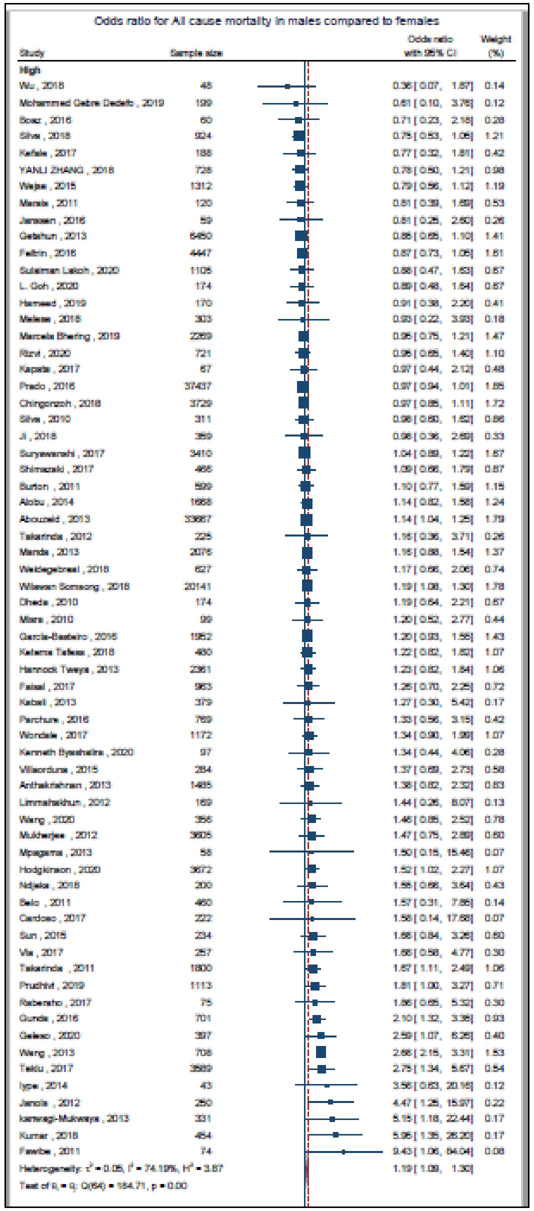

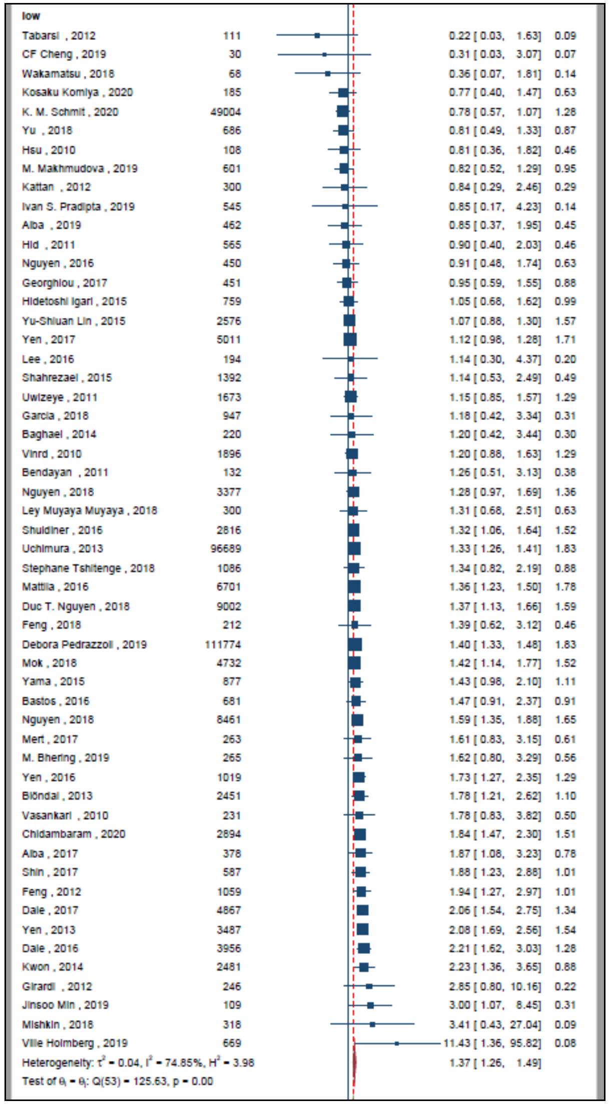

1e) Subgroup analysis based on the TB-HIV coinfection incidence of the study country.

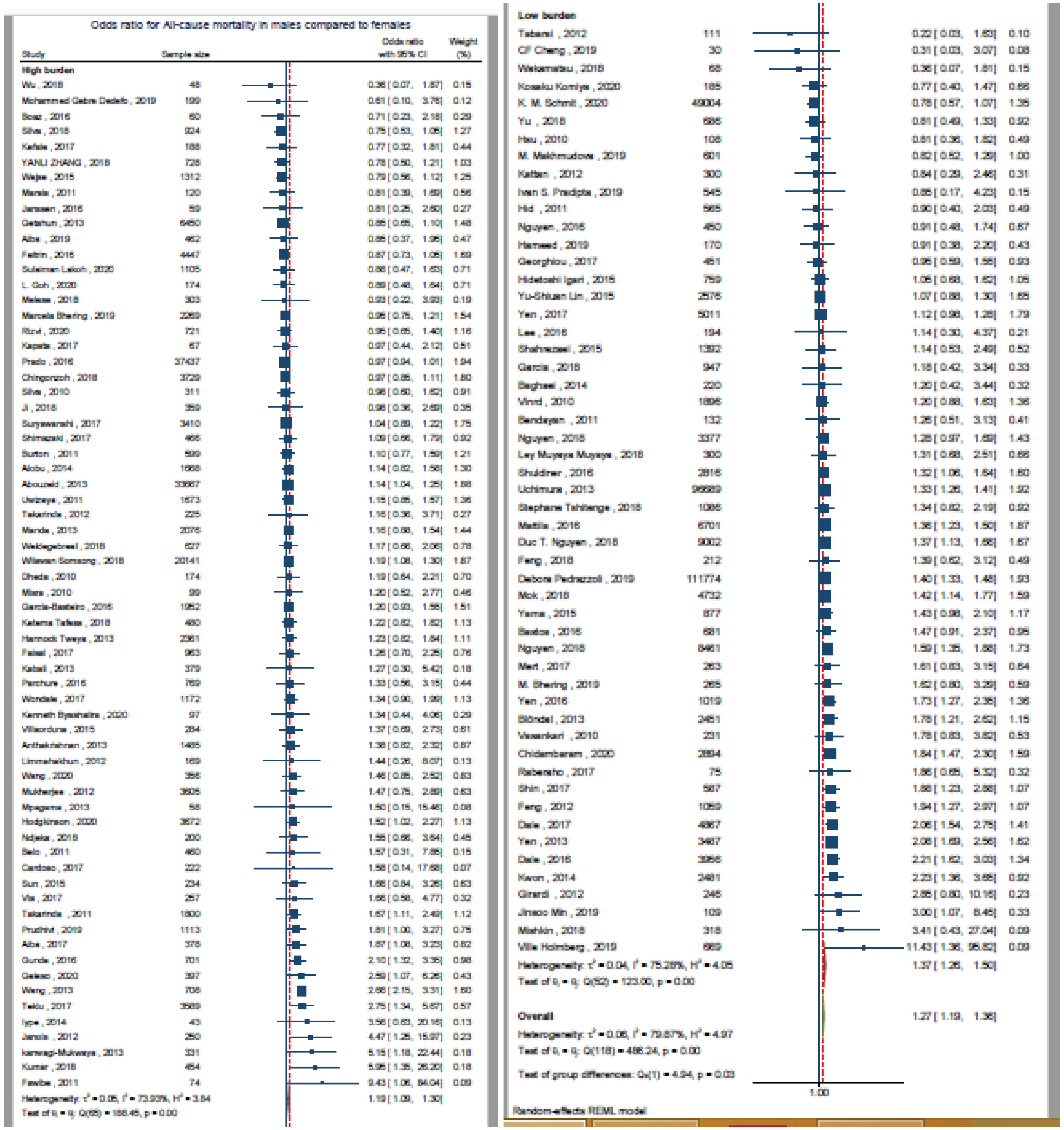

1f) Subgroup analysis based on the drug sensitivity of the study participants.

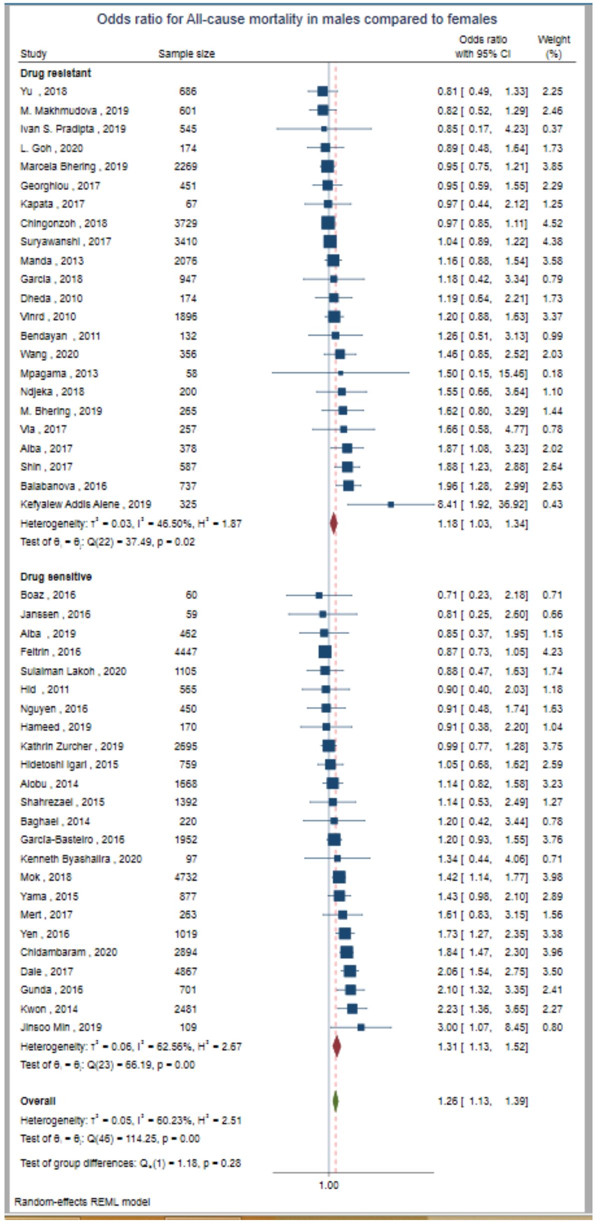

1g) Subgroup analysis based on the site of TB.

Odds ratio for All-cause mortality in males compared to females

| Study | Sample size | Odds ratio with 95% CI | Weight (%) |
|---|---|---|---|
| **Extra-Pulmonary** | | | |
| Wakamatsu , 2018 | 68 | 0.36 [ 0.07, 1.81] | 0.58 |
| Boaz , 2016 | 60 | 0.71 [ 0.23, 2.18] | 1.06 |
| Marais , 2011 | 120 | 0.81 [ 0.39, 1.69] | 1.87 |
| Hsu , 2010 | 108 | 0.81 [ 0.36, 1.82] | 1.68 |
| Rizvi , 2020 | 721 | 0.95 [ 0.65, 1.40] | 3.30 |
| Misra , 2010 | 99 | 1.20 [ 0.52, 2.77] | 1.60 |
| Vinrd , 2010 | 1896 | 1.20 [ 0.88, 1.63] | 3.67 |
| Mert , 2017 | 263 | 1.61 [ 0.83, 3.15] | 2.10 |
| Vasankari , 2010 | 231 | 1.78 [ 0.83, 3.82] | 1.80 |
| Raberaho , 2017 | 75 | 1.86 [ 0.65, 5.32] | 1.16 |
| Iype , 2014 | 43 | 3.56 [ 0.63, 20.16] | 0.51 |
| Heterogeneity: $\tau^2$ = 0.00, $I^2$ = 0.00%, $H^2$ = 1.00 | | 1.13 [ 0.94, 1.36] | |
| Test of $\theta_i = \theta_j$: Q(10) = 9.95, p = 0.45 | | | |
| **Pulmonary** | | | |
| Kosaku Komiya , 2020 | 185 | 0.77 [ 0.40, 1.47] | 2.16 |
| Wejse , 2015 | 1312 | 0.79 [ 0.56, 1.12] | 3.47 |
| Yu , 2018 | 686 | 0.81 [ 0.49, 1.33] | 2.78 |
| M. Makhmudova , 2019 | 601 | 0.82 [ 0.52, 1.29] | 2.96 |
| Aiba , 2019 | 462 | 0.85 [ 0.37, 1.95] | 1.62 |
| Hid , 2011 | 565 | 0.90 [ 0.40, 2.03] | 1.67 |
| Hameed , 2019 | 170 | 0.91 [ 0.38, 2.20] | 1.50 |
| Georghiou , 2017 | 451 | 0.95 [ 0.59, 1.55] | 2.81 |
| Chingonzoh , 2018 | 3729 | 0.97 [ 0.85, 1.11] | 4.40 |
| Ji , 2018 | 359 | 0.98 [ 0.36, 2.69] | 1.24 |
| Hidetoshi Igari , 2015 | 759 | 1.05 [ 0.68, 1.62] | 3.07 |
| Yu-Shiuan Lin , 2015 | 2576 | 1.07 [ 0.88, 1.30] | 4.17 |
| Shimazaki , 2017 | 466 | 1.09 [ 0.66, 1.79] | 2.79 |
| Garcia , 2018 | 947 | 1.18 [ 0.42, 3.34] | 1.18 |
| Wilawan Somsong , 2018 | 20141 | 1.19 [ 1.08, 1.30] | 4.50 |
| Hannock Tweya , 2013 | 2361 | 1.23 [ 0.82, 1.84] | 3.20 |
| Mattila , 2016 | 6701 | 1.36 [ 1.23, 1.50] | 4.49 |
| Yama , 2015 | 877 | 1.43 [ 0.98, 2.10] | 3.32 |
| Mpagama , 2013 | 58 | 1.50 [ 0.15, 15.46] | 0.30 |
| Ndjeka , 2018 | 200 | 1.55 [ 0.66, 3.64] | 1.56 |
| Prudhivi , 2019 | 1113 | 1.81 [ 1.00, 3.27] | 2.39 |
| Chidambaram , 2020 | 2894 | 1.84 [ 1.47, 2.30] | 4.07 |
| Feng , 2012 | 1059 | 1.94 [ 1.27, 2.97] | 3.11 |
| Balabanova , 2016 | 737 | 1.96 [ 1.28, 2.99] | 3.11 |
| Yen , 2013 | 3487 | 2.08 [ 1.69, 2.56] | 4.12 |
| Kwon , 2014 | 2481 | 2.23 [ 1.36, 3.65] | 2.80 |
| Wang , 2013 | 708 | 2.66 [ 2.15, 3.31] | 4.09 |
| Teklu , 2017 | 3589 | 2.75 [ 1.34, 5.67] | 1.92 |
| Janols , 2012 | 250 | 4.47 [ 1.25, 15.97] | 0.86 |
| Kefyalew Addis Alene , 2019 | 325 | 8.41 [ 1.92, 36.92] | 0.67 |
| Fawibe , 2011 | 74 | 9.43 [ 1.06, 84.04] | 0.33 |
| Heterogeneity: $\tau^2$ = 0.11, $I^2$ = 84.44%, $H^2$ = 6.43 | | 1.36 [ 1.17, 1.59] | |
| Test of $\theta_i = \theta_j$: Q(30) = 151.21, p = 0.00 | | | |
| **Overall** | | 1.32 [ 1.15, 1.50] | |
| Heterogeneity: $\tau^2$ = 0.10, $I^2$ = 78.65%, $H^2$ = 4.68 | | | |
| Test of $\theta_i = \theta_j$: Q(41) = 163.34, p = 0.00 | | | |
| Test of group differences: $Q_b(1)$ = 2.29, p = 0.13 | | | |

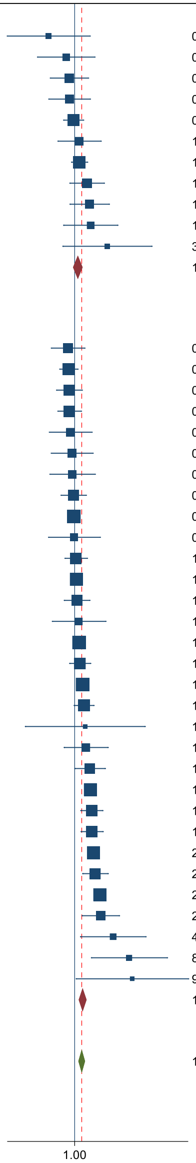

Random-effects REML model

**eFigure 2.** Pooled odds ratio for all-cause mortality in male patients compared to female patients. (Adjusted)

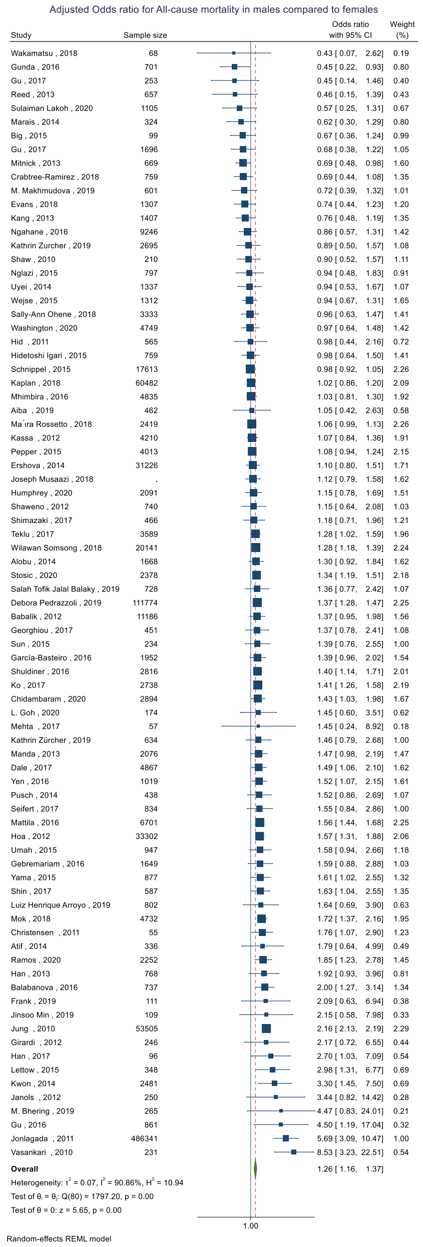

**eFigure 3.** Pooled hazard ratio for all-cause mortality in male patients compared to female patients. (Unadjusted)

3a) Forest plot

Hazards ratio for All-cause mortality in males compared to females

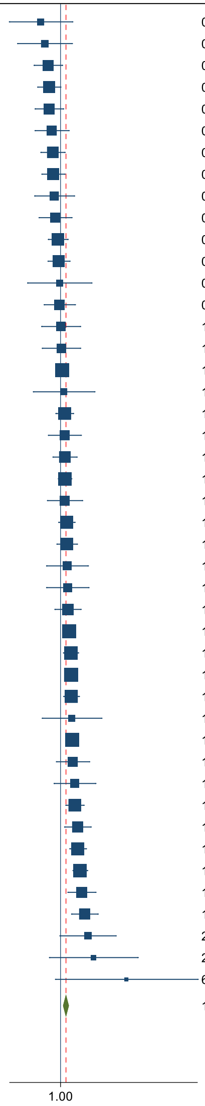

| Study | Sample size | Hazards ratio with 95% CI | Weight (%) |
|---|---|---|---|
| Geleso , 2020 | 397 | 0.57 [ 0.23, 1.40] | 0.73 |
| Reed , 2013 | 657 | 0.64 [ 0.29, 1.39] | 0.92 |
| Crabtree-Ramirez , 2018 | 759 | 0.70 [ 0.47, 1.05] | 2.19 |
| Blanc , 2011 | 661 | 0.72 [ 0.52, 1.00] | 2.64 |
| YANLI ZHANG , 2018 | 728 | 0.72 [ 0.48, 1.08] | 2.18 |
| Evans , 2018 | 1307 | 0.78 [ 0.48, 1.26] | 1.80 |
| Mitnick , 2013 | 669 | 0.80 [ 0.57, 1.12] | 2.55 |
| Wejse , 2015 | 1312 | 0.81 [ 0.59, 1.12] | 2.65 |
| Kosaku Komiya , 2020 | 185 | 0.84 [ 0.48, 1.48] | 1.47 |
| Pietersen , 2014 | 107 | 0.86 [ 0.54, 1.37] | 1.87 |
| Villaorduna , 2015 | 284 | 0.93 [ 0.70, 1.23] | 2.96 |
| Thao , 2017 | 1699 | 0.95 [ 0.70, 1.29] | 2.75 |
| Ji , 2018 | 359 | 0.97 [ 0.39, 2.42] | 0.71 |
| Shaweno , 2012 | 740 | 0.97 [ 0.62, 1.51] | 1.99 |
| Salah Tofik Jalal Balaky , 2019 | 728 | 1.01 [ 0.58, 1.75] | 1.52 |
| Gu , 2017 | 1696 | 1.02 [ 0.59, 1.76] | 1.55 |
| Pepper , 2015 | 4013 | 1.05 [ 0.91, 1.21] | 3.91 |
| Palacios , 2012 | 52 | 1.10 [ 0.46, 2.64] | 0.77 |
| Farley , 2011 | 757 | 1.12 [ 0.87, 1.44] | 3.17 |
| Horita , 2013 | 432 | 1.12 [ 0.71, 1.79] | 1.88 |
| Joseph Musaazi , 2018 | . | 1.13 [ 0.80, 1.59] | 2.55 |
| Shuldiner , 2016 | 2816 | 1.13 [ 0.94, 1.36] | 3.59 |
| Shaw , 2010 | 210 | 1.13 [ 0.69, 1.86] | 1.73 |
| Kassa , 2012 | 4210 | 1.19 [ 0.95, 1.50] | 3.31 |
| Ershova , 2014 | 31226 | 1.20 [ 0.90, 1.60] | 2.90 |
| Tara Catherine Bouton , 2019 | 394 | 1.21 [ 0.67, 2.19] | 1.38 |
| Ley Muyaya Muyaya , 2018 | 300 | 1.22 [ 0.67, 2.23] | 1.36 |
| Humphrey , 2020 | 2091 | 1.23 [ 0.85, 1.78] | 2.37 |
| Kaplan , 2018 | 60482 | 1.28 [ 1.09, 1.50] | 3.78 |
| Teklu , 2017 | 3589 | 1.34 [ 1.08, 1.65] | 3.43 |
| Van'tHoog , 2012 | 10875 | 1.35 [ 1.18, 1.55] | 3.93 |
| Kim , 2010 | 75 | 1.36 [ 1.09, 1.69] | 3.37 |
| Parchure , 2016 | 769 | 1.38 [ 0.59, 3.22] | 0.81 |
| Wickett , 2018 | 560 | 1.40 [ 1.20, 1.64] | 3.79 |
| Wang , 2020 | 356 | 1.42 [ 0.89, 2.27] | 1.85 |
| Sun , 2015 | 234 | 1.50 [ 0.83, 2.71] | 1.39 |
| Horne , 2010 | 3451 | 1.50 [ 1.16, 1.94] | 3.12 |
| Ba´ez-Saldaña , 2016 | 804 | 1.63 [ 1.12, 2.37] | 2.34 |
| Kang , 2013 | 1407 | 1.64 [ 1.30, 2.07] | 3.28 |
| Chidambaram , 2020 | 2894 | 1.75 [ 1.43, 2.15] | 3.47 |
| Feng , 2012 | 1059 | 1.83 [ 1.23, 2.72] | 2.23 |
| Balabanova , 2016 | 737 | 1.99 [ 1.37, 2.90] | 2.34 |
| Jeon , 2011 | 202 | 2.18 [ 0.98, 4.84] | 0.89 |
| Calligaro , 2015 | 341 | 2.57 [ 0.73, 9.08] | 0.40 |
| Ville Holmberg , 2019 | 669 | 6.60 [ 0.86, 50.46] | 0.17 |
| **Overall** | | 1.17 [ 1.07, 1.27] | |

Heterogeneity: $\tau^2$ = 0.04, $I^2$ = 64.53%, $H^2$ = 2.82

Test of $\theta_i = \theta_j$: Q(44) = 110.62, p = 0.00

Test of $\theta$ = 0: z = 3.59, p = 0.00

Random-effects REML model

3b) Subgroup analysis based on the time of outcome assessment.

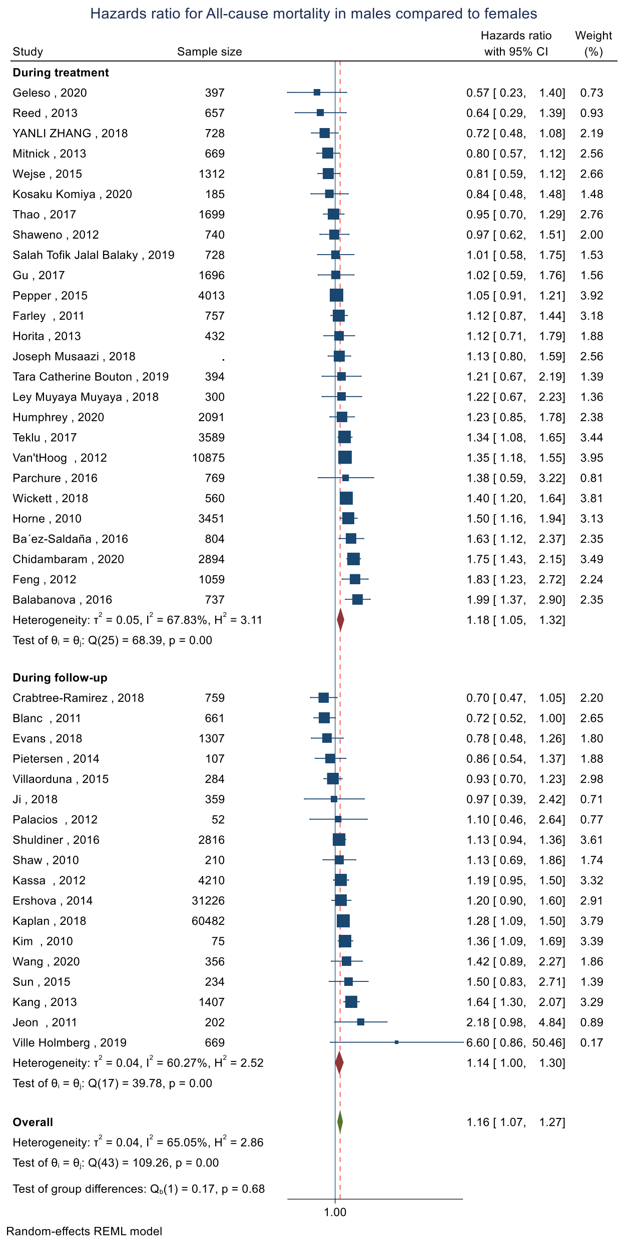

3c) Subgroup analysis based on the World Bank income status classification.

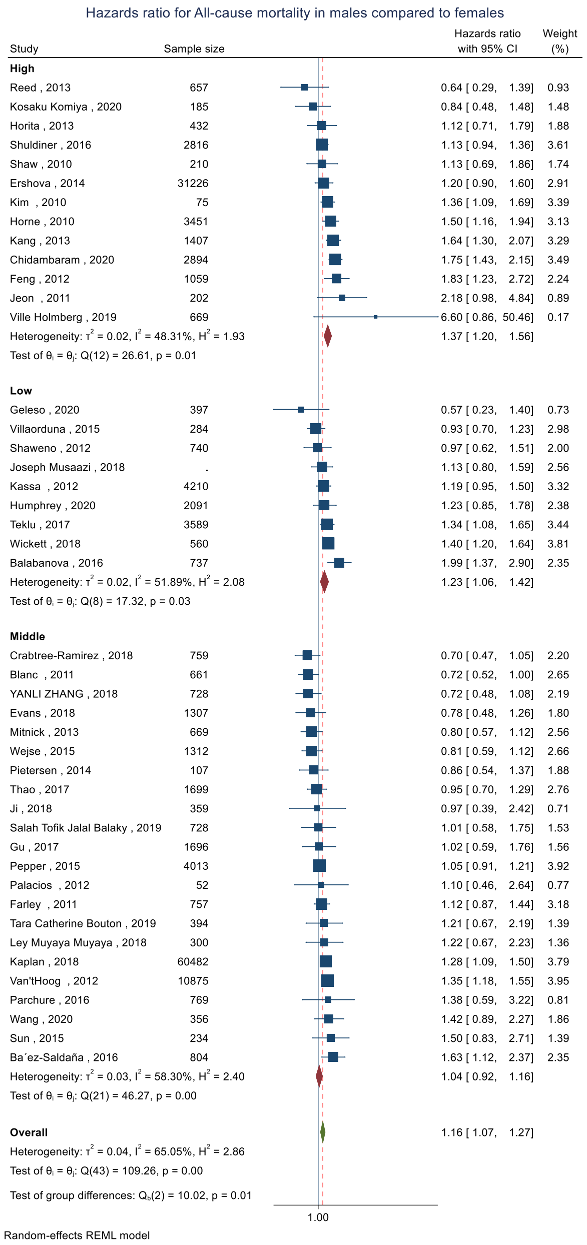

3d) Subgroup analysis based on the TB incidence of the study country.

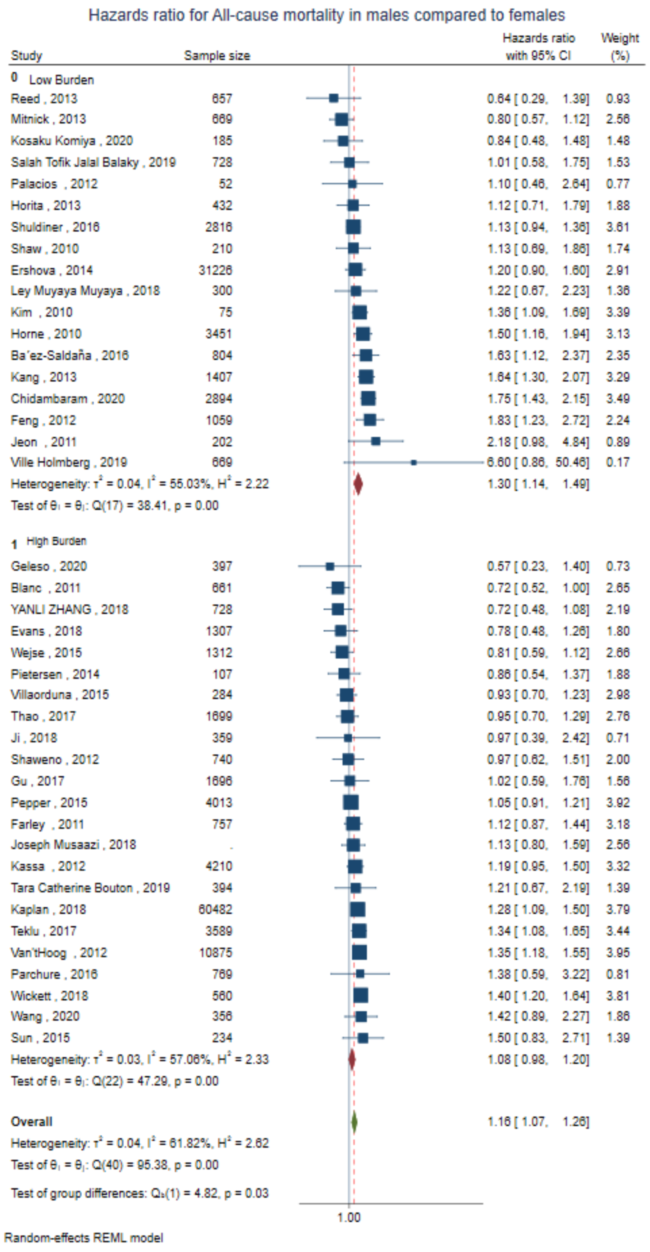

3e) Subgroup analysis based on the TB-HIV coinfection incidence of the study country.

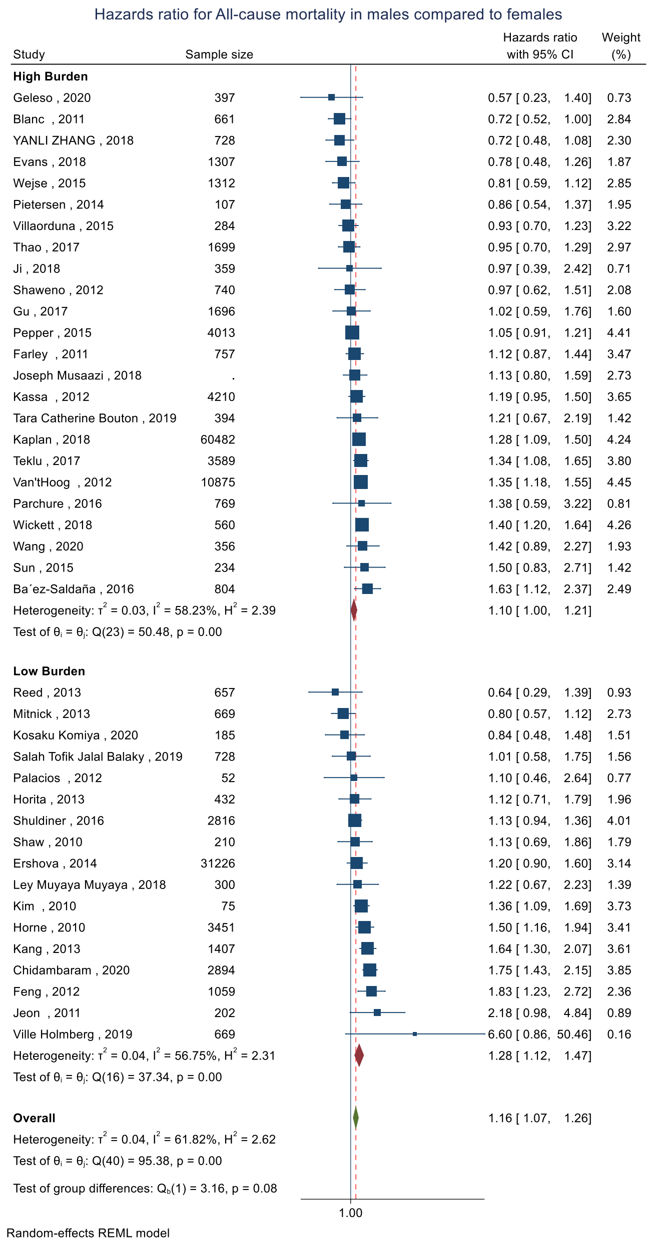

3f) Subgroup analysis based on the drug sensitivity of the study participants.

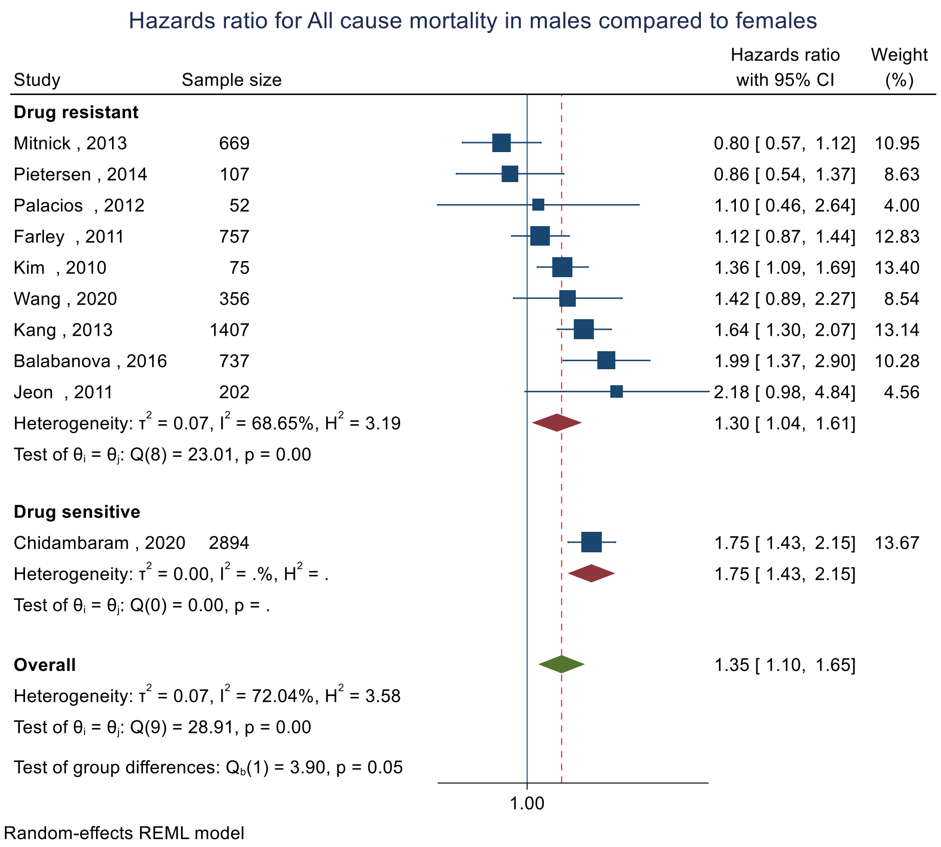

3g) Subgroup analysis based on the site of TB

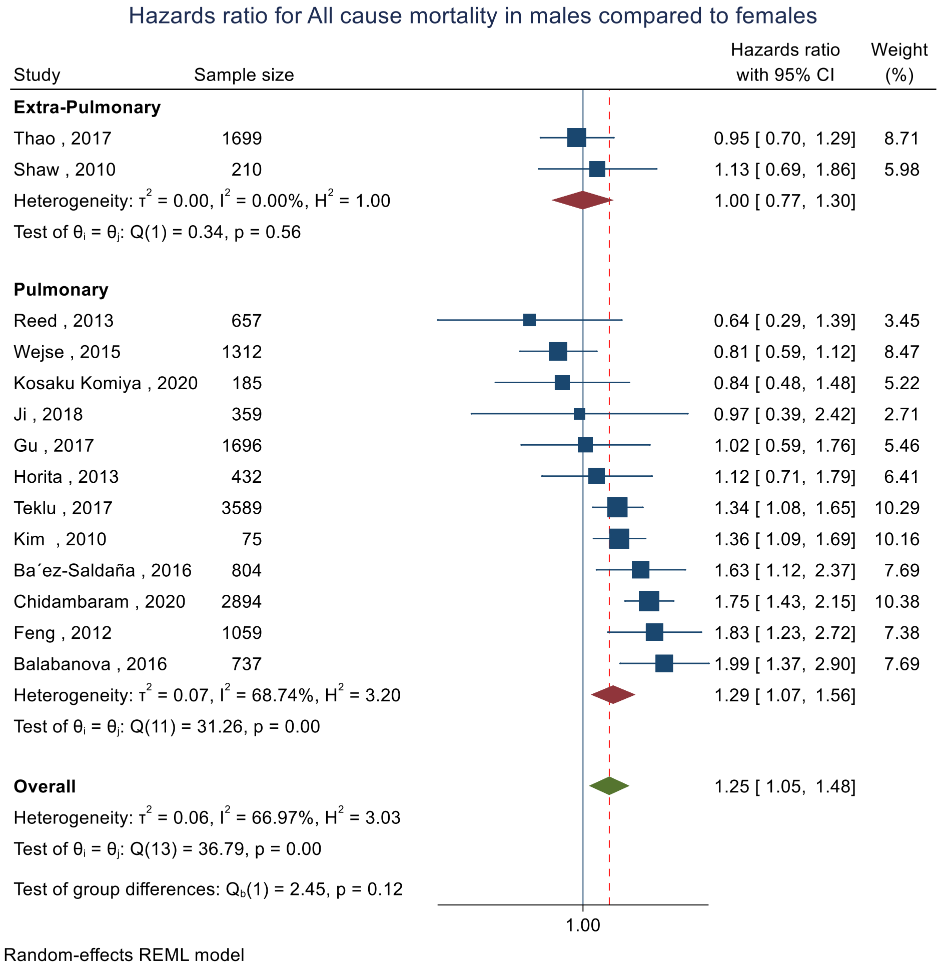

**eFigure 4.** Pooled hazard ratio for all-cause mortality in male patients compared to female patients. (Adjusted)

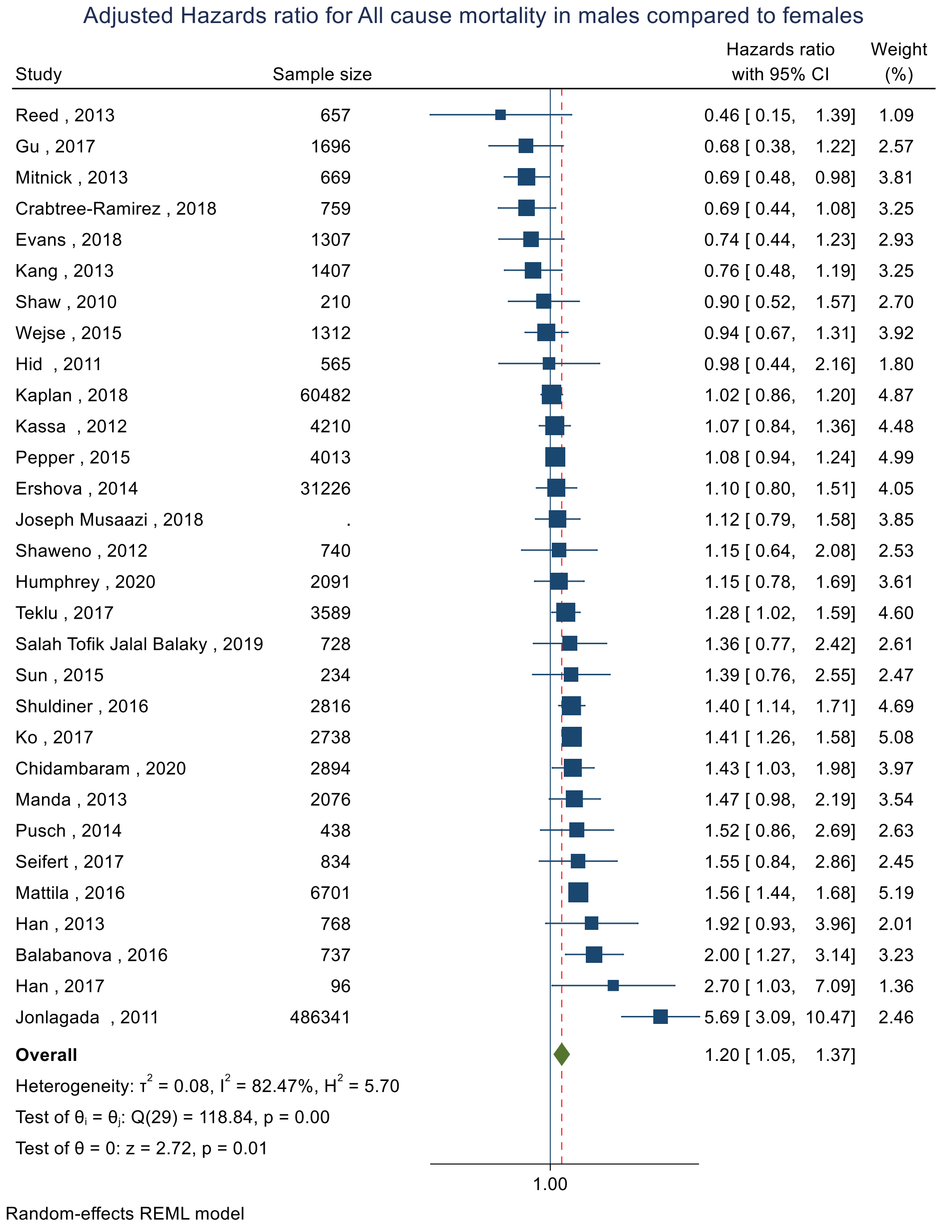

eFigure 5 Change in log-odds for mortality in males compared to females with change in default between males and females.

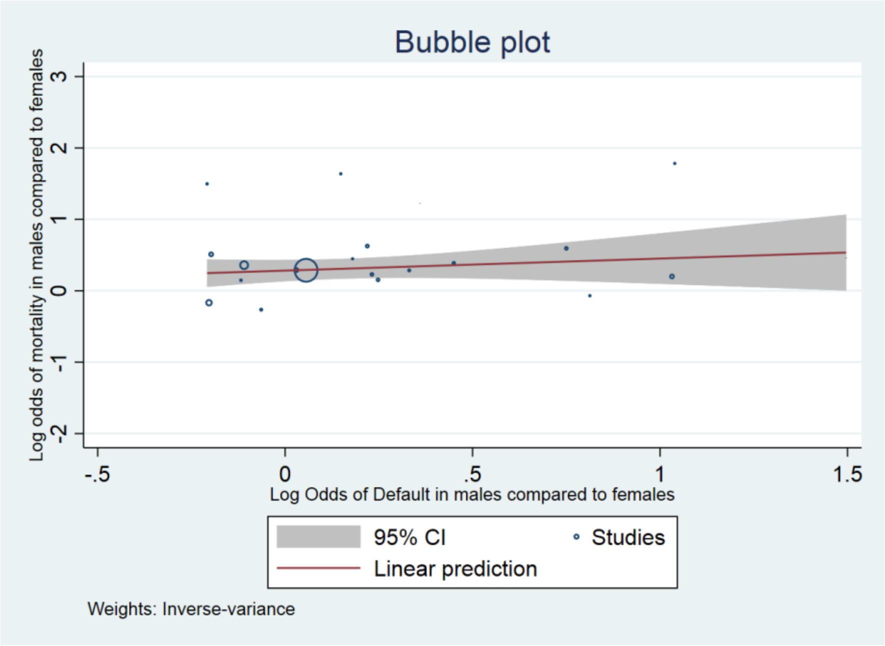

eFigure 6 Change in log-odds for mortality in males compared to females with change in LTFU between males and females.

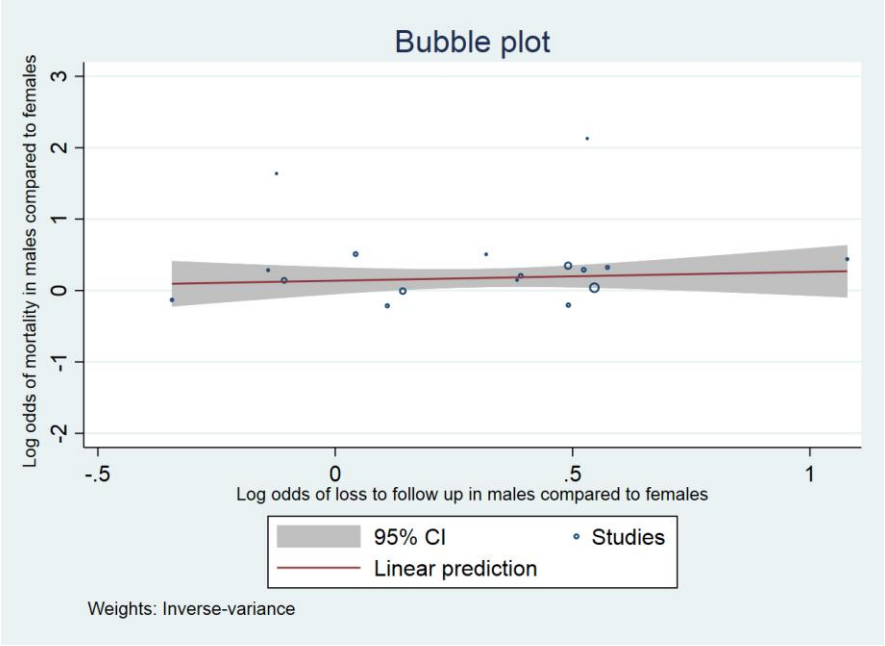

**eFigure 7.** Change in log-odds for mortality in males compared to females with change in treatment completion between males and females.

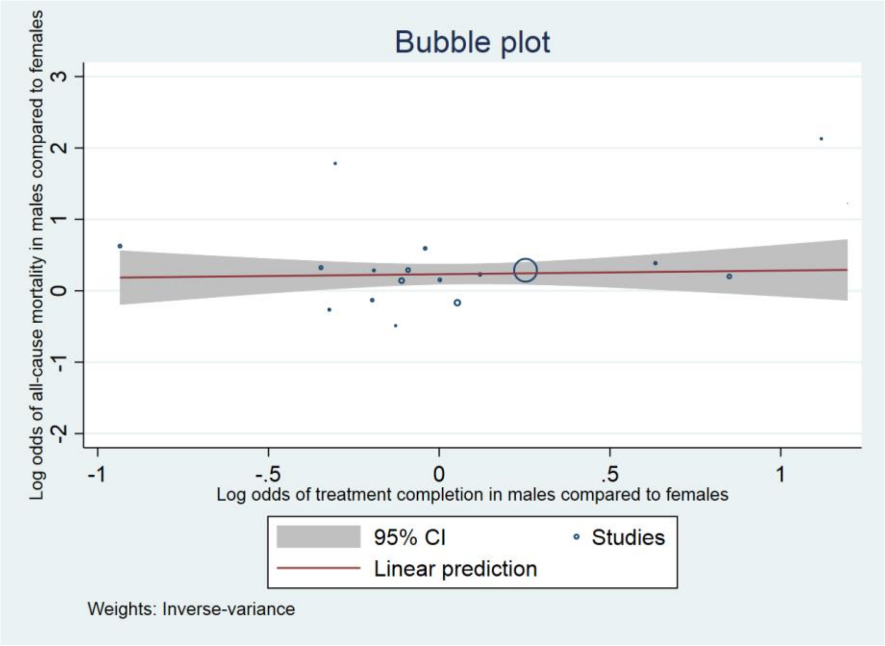

**eFigure 8.** Change in log-odds for mortality in males compared to females with change in HIV between males and females.

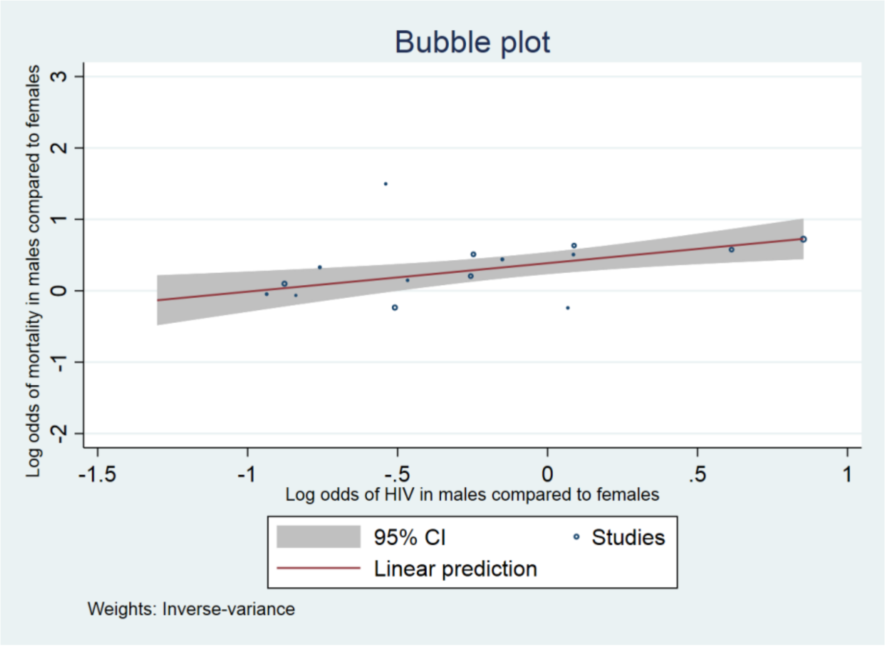

**eFigure 9.** Pooled odds ratio for sputum smear AFB positivity after treatment initiation in male patients compared to female patients. (Unadjusted)

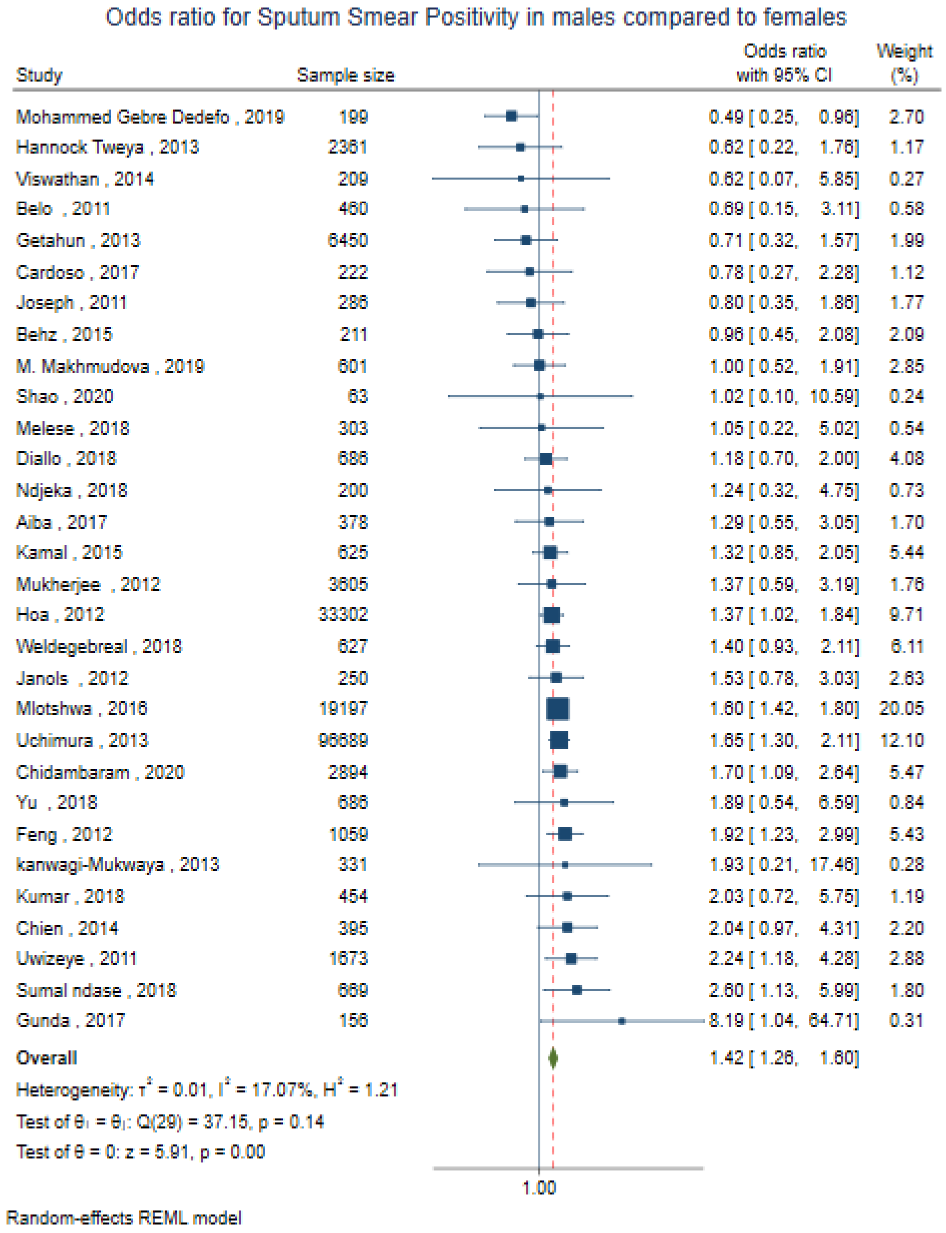

**eFigure 10.** Pooled odds ratio for sputum smear AFB positivity after treatment initiation in male patients compared to female patients. (Adjusted)

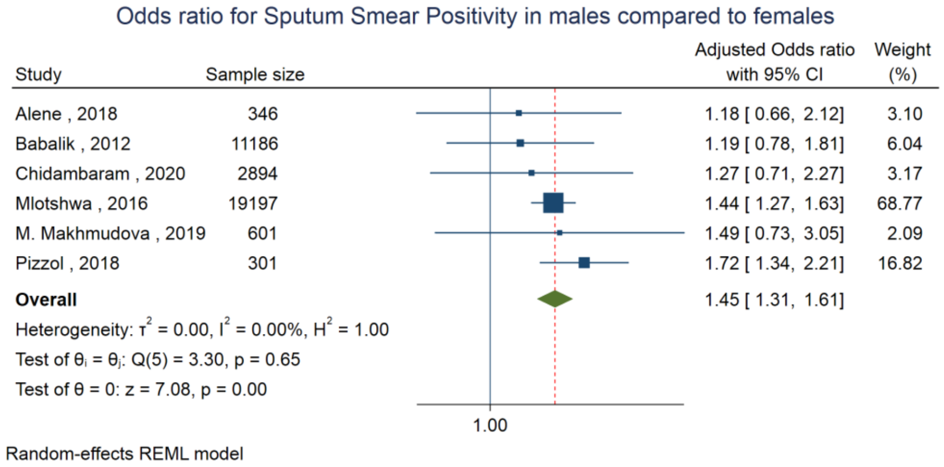

**eFigure 11.** Pooled odds ratio for sputum culture positivity after treatment initiation in male patients compared to female patients. (Unadjusted)

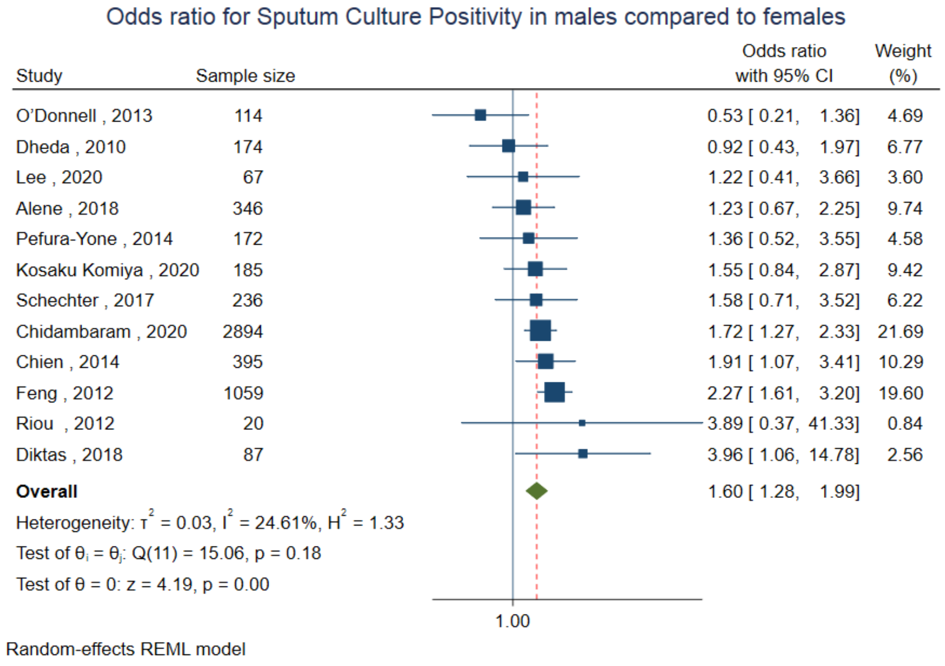

**eFigure 12.** Pooled odds ratio for sputum culture positivity after treatment initiation in male patients compared to female patients. (Adjusted)

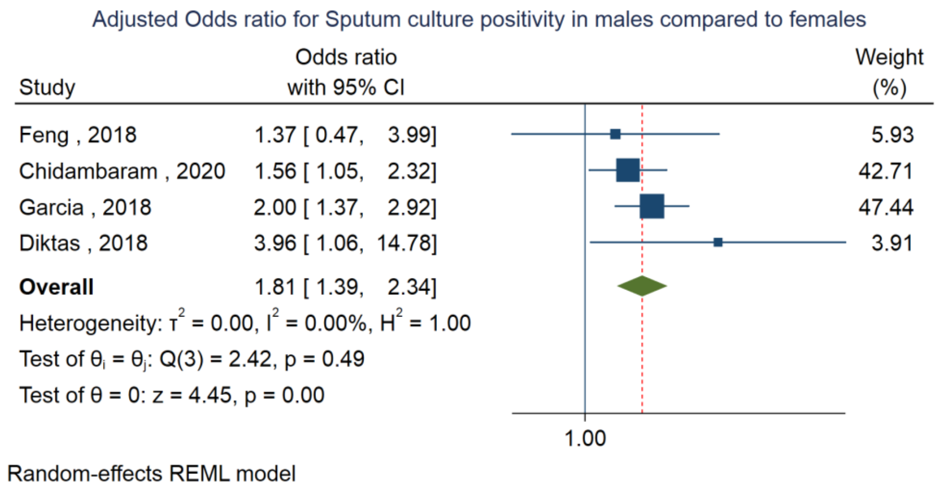

**eFigure 13.** Pooled hazard ratio for sputum culture positivity after treatment initiation in male patients compared to female patients. (Unadjusted)

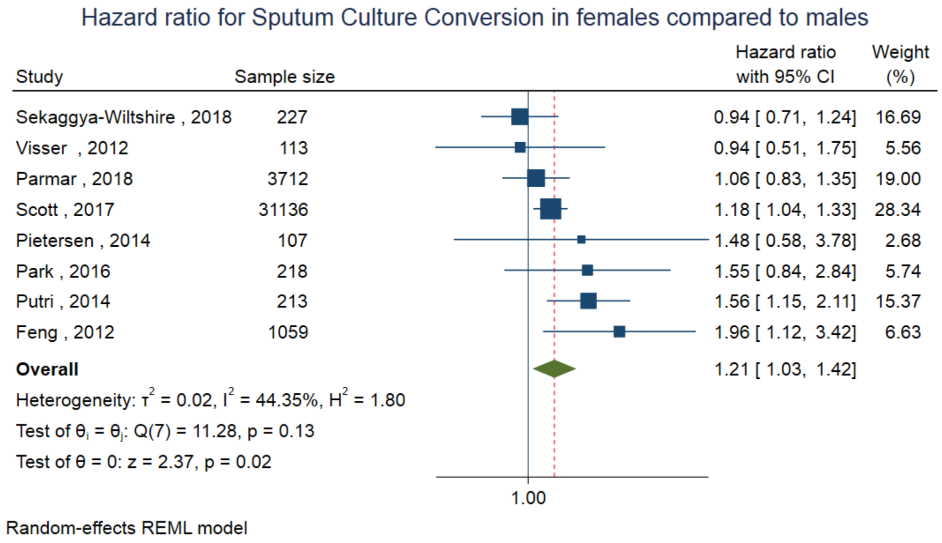

**eFigure 14.** Pooled hazard ratio for sputum culture positivity after treatment initiation in male patients compared to female patients. (Adjusted)

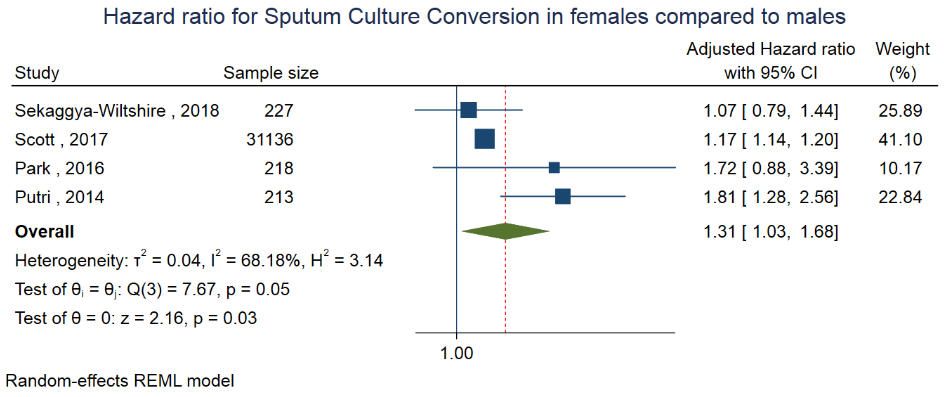

**eFigure 15.** Pooled odds ratio for treatment success positivity after treatment initiation in male patients compared to female patients. (Unadjusted)

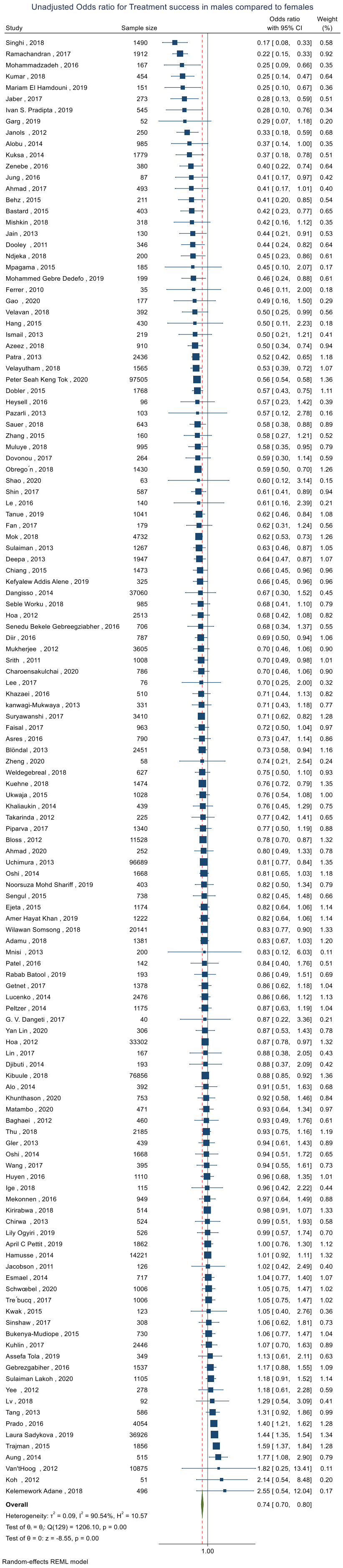

**eFigure 16.** Pooled odds ratio for treatment success positivity after treatment initiation in male patients compared to female patients. (Adjusted)

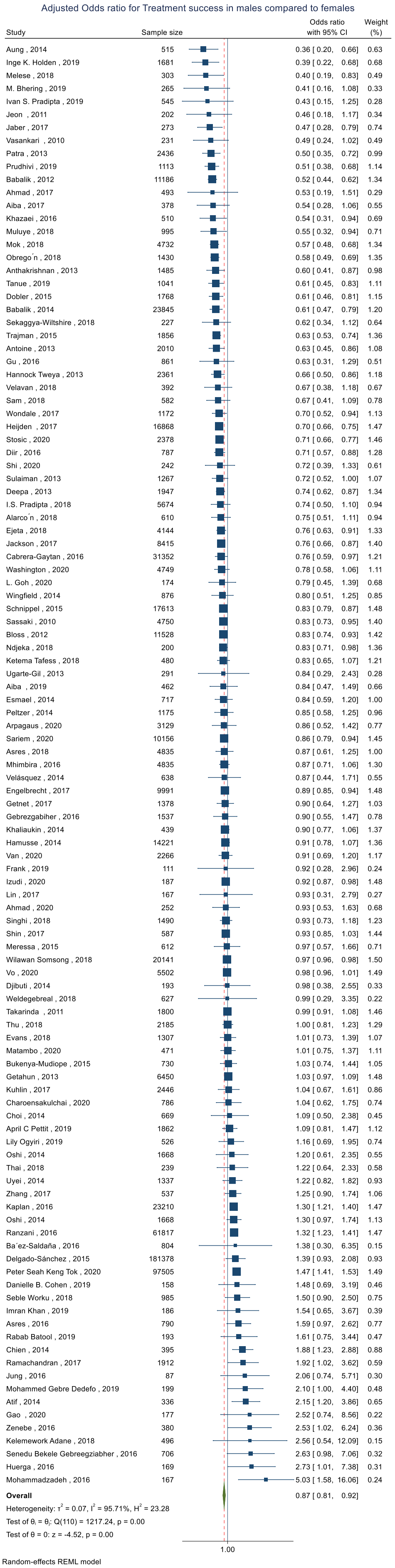

**eFigure 17.** Funnel Plot for all-cause mortality a) Odds ratio b) Hazard ratio

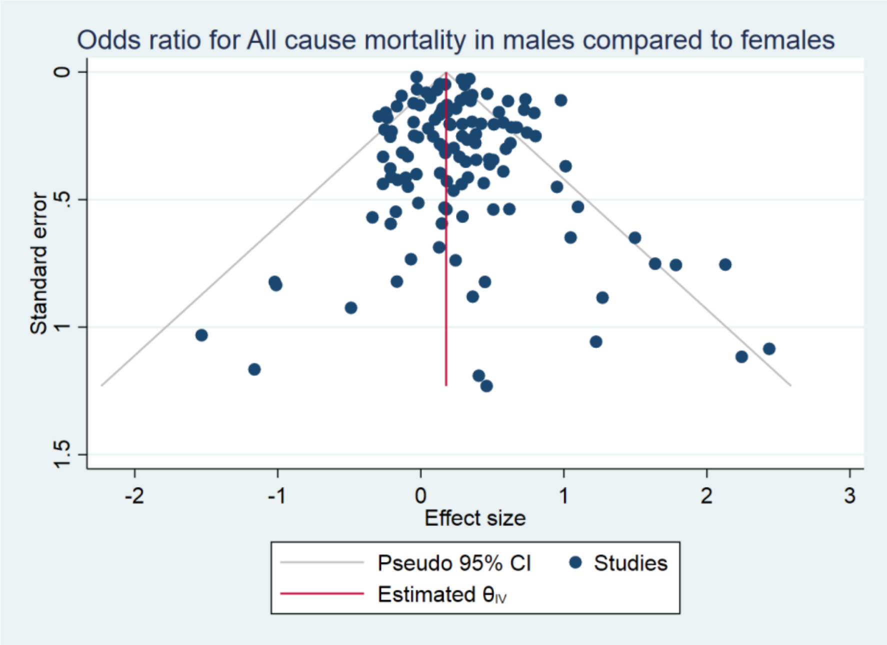

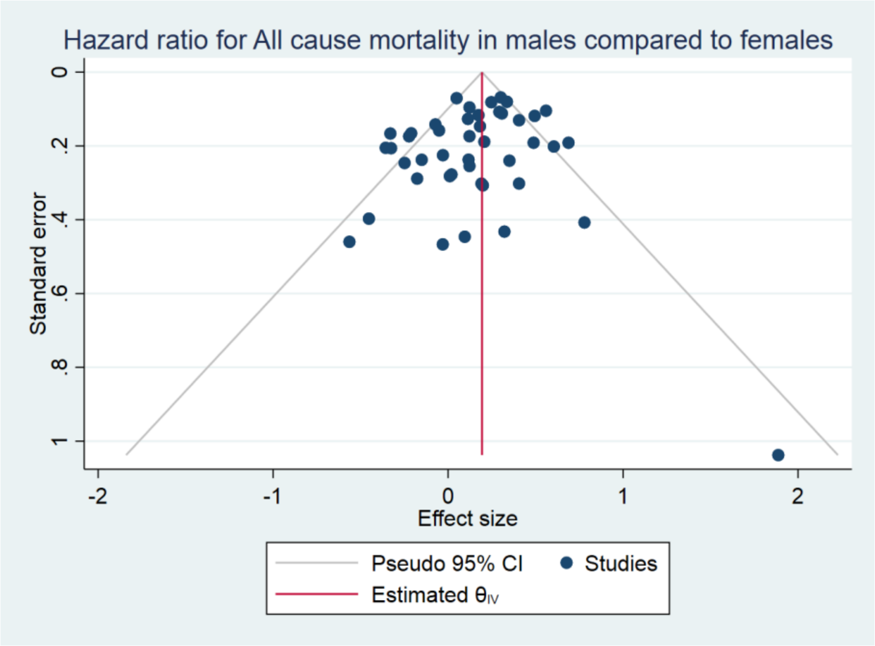

**eFigure 18.** Funnel Plot for a) Sputum culture Positivity b) Sputum smear positivity

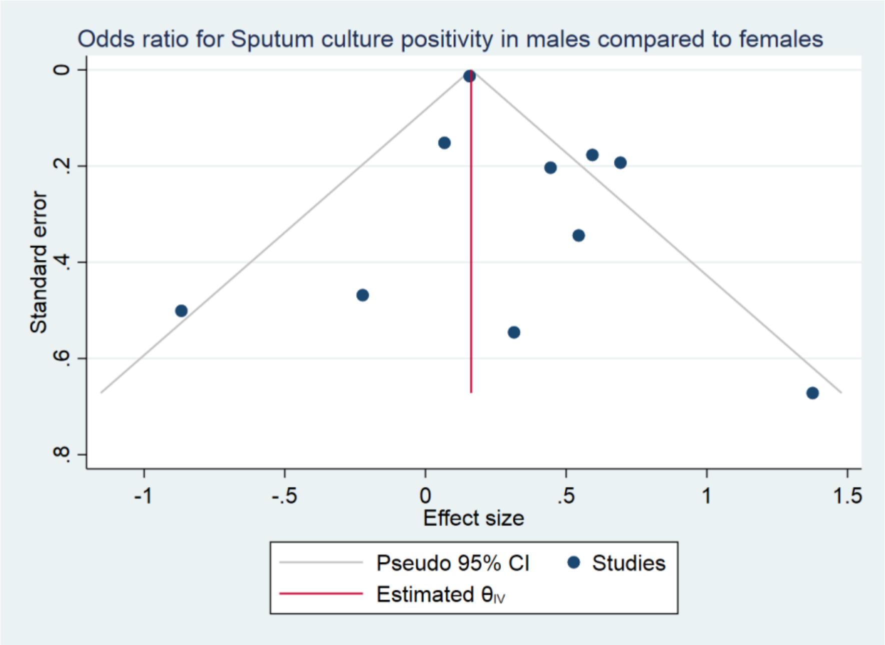

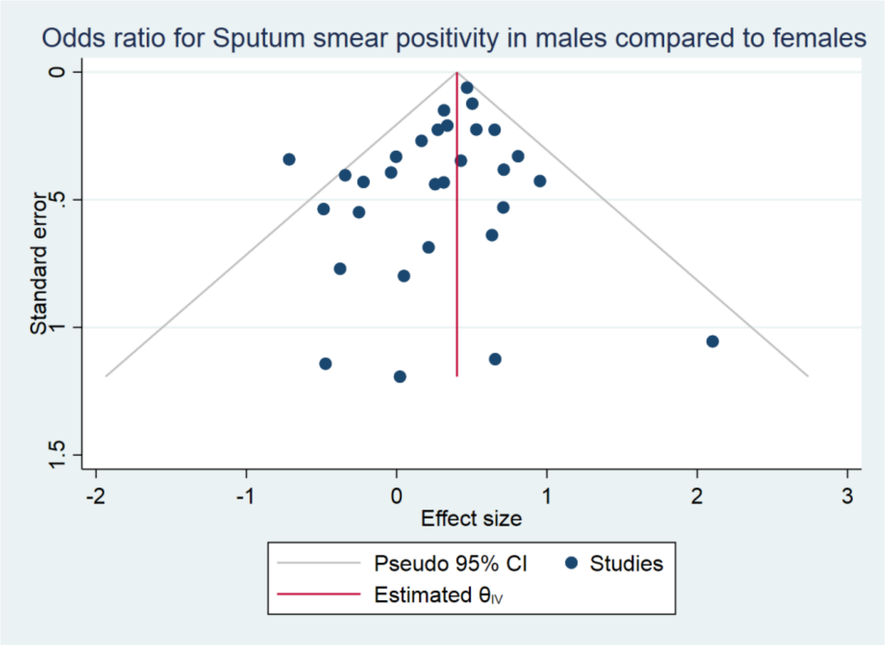

**eFigure 19.** Funnel Plot for treatment success

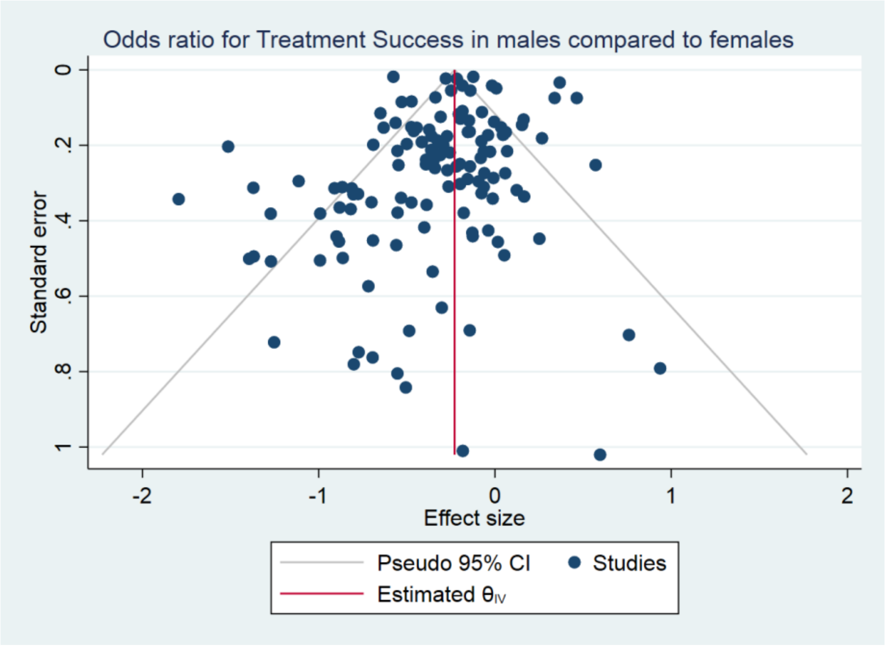

### Section-V

16 Ramos JM, Comeche B, Tesfamariam A, *et al.* Sex differences and HIV status of tuberculosis in adults at a rural

hospital in southern Ethiopia: an 18-year retrospective cross-sectional study. *Afri Heal Sci* 2020; **20**: 605.

17 Van LH, Phu PT, Vinh DN, *et al.* Risk factors for poor treatment outcomes of 2266 multidrug-resistant tuberculosis cases in Ho Chi Minh City: A retrospective study. *BMC Infect Dis* 2020; **20**. DOI:10.1186/s12879-020-4887-1.

18 Singla R, Raghu B, Gupta A, *et al.* Risk factors for early mortality in patients with pulmonary tuberculosis admitted to the emergency room. *Pulmonology* 2020. DOI:10.1016/j.pulmoe.2020.02.002.

19 Shao Y, Li Y, Song H, *et al.* A retrospective cohort study of isoniazid-resistant tuberculosis treatment outcomes and isoniazid resistance-associated mutations in eastern China from 2013 to 2018. *J Glob Antimicrob Resist* 2020. DOI:10.1016/j.jgar.2020.07.012.

20 Olayanju O, Esmail A, Limberis J, Dheda K. A regimen containing bedaquiline and delamanid compared to bedaquiline in patients with drug-resistant tuberculosis. *Eur Respir J* 2020; **55**. DOI:10.1183/13993003.01181-2019.

21 Lee H, Sohn JW, Sim YS, Shin TR, Kim DG, Choi H. Outcomes of extended duration therapy for drug-susceptible cavitary pulmonary tuberculosis. *Ann Transl Med* 2020; **8**. DOI:10.21037/atm.2020.02.104.

22 Schwœbel V, Trébucq A, Kashongwe Z, *et al.* Outcomes of a nine-month regimen for rifampicin-resistant tuberculosis up to 24 months after treatment completion in nine African countries. *EClinicalMedicine* 2020; **20**. DOI:10.1016/j.eclinm.2020.100268.

23 Humphrey JM, Mpofu P, Pettit AC, *et al.* Mortality among people with HIV treated for tuberculosis based on positive, negative, or no bacteriologic test results for tuberculosis: The IEDEA consortium. *Open Forum Infect Dis* 2020; **7**. DOI:10.1093/ofid/ofaa006.

24 Piubello A, Souleymane MB, Hassane-Harouna S, *et al.* Management of multidrug-resistant tuberculosis with shorter treatment regimen in Niger: Nationwide programmatic achievements. *Respir Med* 2020; **161**. DOI:10.1016/j.rmed.2019.105844.

25 Shi W, Davies Forsman L, Hu Y, *et al.* Improved treatment outcome of multidrug-resistant tuberculosis with the use of a rapid molecular test to detect drug resistance in China. *Int J Infect Dis* 2020; **96**: 390–7.

26 Gao M, Gao J, Xie L, *et al.* Early outcome and safety of bedaquiline-containing regimens for treatment of MDR- and XDR-TB in China: a multicentre study. *Clin Microbiol Infect* 2020. DOI:10.1016/j.cmi.2020.06.004.

27 Rizvi I, Malhotra HS, Garg RK, Kumar N. Derivation of a bedside score (MASH-P) to predict 6-month mortality in tuberculous meningitis. *J Neurol Sci* 2020; **415**. DOI:10.1016/j.jns.2020.116877.

28 Charoensakulchai S, Limsakul M, Saengungsumalee I, *et al.* Characteristics of poor tuberculosis treatment outcomes among patients with pulmonary tuberculosis in community hospitals of Thailand. *Am J Trop Med Hyg* 2020; **102**: 553–61.

29 Lakoh S, Jiba DF, Adekanmbi O, *et al.* Diagnosis and treatment outcomes of adult tuberculosis in an urban setting with high HIV prevalence in Sierra Leone: A retrospective study. *Int J Infect Dis* 2020; **96**: 112–8.

30 Alene KA, Viney K, Yi H, *et al.* Comparison of the validity of smear and culture conversion as a prognostic marker of treatment outcome in patients with multidrug-resistant tuberculosis. *PLoS One* 2018; **13**: e0197880.

31 Tok PSK, Liew SM, Wong LP, *et al.* Determinants of unsuccessful treatment outcomes and mortality among tuberculosis patients in Malaysia: A registry-based cohort study. *PLoS One* 2020; **15**: e0231986.

32 Lin Y, Bai Y, Zhang T, *et al.* Unfavourable treatment outcomes in tuberculosis patients with different vitamin D status and blood glucose levels in a programme setting in China. *Trop Med Int Heal* 2020; **25**: 373–9.

33 Schmit KM, Shah N, Kammerer S, Bamrah Morris S, Marks SM. Tuberculosis Transmission or Mortality Among

Persons Living with HIV, USA, 2011-2016. *J racial Ethn Heal disparities* 2020. DOI:10.1007/s40615-020-00709-7.

34 Gonah L, Maphosa TM. Association of MDR-TB treatment outcomes and HIV status in Zimbabwe: A retrospective study. *Cogent Med* 2020; **7**. DOI:10.1080/2331205X.2020.1783129.

35 Tanue EA, Nsagha DS, Njamen TN, Assob NJC. Tuberculosis treatment outcome and its associated factors among people living with HIV and AIDS in Fako Division of Cameroon. *PLoS One* 2019; **14**: e0218800.

36 Makhmudova M, Maxsumova Z, Rajabzoda A, Makhmadov A, van den Hof S, Mirtskhulava V. Risk factors for unfavourable treatment outcomes among rifampicin-resistant tuberculosis patients in Tajikistan. *Int J Tuberc Lung Dis* 2019; **23**: 331–6.

37 Cheng CF, Huang YM, Lu CH, Hsieh SC, Li KJ. Prednisolone dose during treatment of tuberculosis might be a risk factor for mortality in patients with systemic lupus erythematosus: a hospital-based cohort study. *Lupus* 2019; **28**: 1699–704.

38 Pradipta IS, Van't Boveneind-Vrubleuskaya N, Akkerman OW, Alffenaar JWC, Hak E. Predictors for treatment outcomes among patients with drug-susceptible tuberculosis in the Netherlands: a retrospective cohort study. *Clin Microbiol Infect* 2019; **25**: 761.e1-761.e7.

39 Holden IK, Lillebaek T, Seersholm N, Andersen PH, Wejse C, Johansen IS. Predictors for Pulmonary Tuberculosis Treatment Outcome in Denmark 2009-2014. *Sci Rep* 2019; **9**: 12995.

40 Arroyo LH, Ramos AC V, Yamamura M, *et al.* Predictive model of unfavorable outcomes for multidrug-resistant tuberculosis. *Rev Saude Publica* 2019; **53**: 77.

41 Bhering M, Duarte R, Kritski A. Predictive factors for unfavourable treatment in MDR-TB and XDR-TB patients in Rio de Janeiro State, Brazil, 2000-2016. *PLoS One* 2019; **14**: e0218299.

42 Cohen DB, Davies G, Malwafu W, *et al.* Poor outcomes in recurrent tuberculosis: More than just drug resistance? *PLoS One* 2019; **14**: e0215855.

43 Bhering M, Kritski A, Nunes C, Duarte R. Multidrug-resistant tuberculosis in Lisbon: unfavourable treatment and associated factors, 2000-2014. *Int J Tuberc Lung Dis* 2019; **23**: 1075–81.

44 Komiya K, Goto A, Kan T, *et al.* A high C-reactive protein level and poor performance status are associated with delayed sputum conversion in elderly patients with pulmonary tuberculosis in Japan. *Clin Respir J* 2020; **14**: 291–8.

45 Mahwire TC, Zunza M, Marukutira TC, Naidoo P. Impact of Xpert MTB/RIF assay on multidrug-resistant tuberculosis treatment outcomes in a health district in South Africa. *S Afr Med J* 2019; **109**: 259–63.

46 Shariff NM, Shah SA, Kamaludin F. Impact of ethnic disparities on the treatment outcomes of HIV-negative drug-resistant tuberculosis patients in Kuala Lumpur, Malaysia: A call for a culturally-sensitive community intervention approach. *J Glob Antimicrob Resist* 2019; **19**: 274–9.

47 Bouton TC, Forson A, Kudzawu S, *et al.* High mortality during tuberculosis retreatment at a Ghanaian tertiary center: a retrospective cohort study. *Pan Afr Med J* 2019; **33**: 111.

48 Ogyiri L, Lartey M, Ojewale O, *et al.* Effect of HIV infection on TB treatment outcomes and time to mortality in two urban hospitals in Ghana-a retrospective cohort study. *Pan Afr Med J* 2019; **32**: 206.

49 Zürcher K, Ballif M, Fenner L, *et al.* Drug susceptibility testing and mortality in patients treated for tuberculosis in high-burden countries: a multicentre cohort study. *Lancet Infect Dis* 2019; **19**: 298–307.

50 Pettit AC, Jenkins CA, Blevins Peratikos M, *et al.* Directly observed therapy and risk of unfavourable tuberculosis treatment outcomes among an international cohort of people living with HIV in low- and middle-income countries. *J Int AIDS Soc* 2019; **22**: e25423.

51 Zürcher K, Ballif M, Kiertiburanakul S, *et al.* Diagnosis and clinical outcomes of extrapulmonary tuberculosis in antiretroviral therapy programmes in low- and middle-income countries: a multicohort study. *J Int AIDS Soc* 2019; **22**: e25392.

52 Golub JE, Mok Y, Hong S, Jung KJ, Jee SH, Samet JM. Diabetes mellitus and tuberculosis in Korean adults: impact on tuberculosis incidence, recurrence and mortality. *Int J Tuberc Lung Dis* 2019; **23**: 507–13.

53 Byashalira K, Mbelele P, Semvua H, *et al.* Clinical outcomes of new algorithm for diagnosis and treatment of Tuberculosis sepsis in HIV patients. *Int J Mycobacteriol* 2019; **8**: 313–9.

54 Min J, Mi Shin Y, Lee WJ, *et al.* Clinical features of octogenarian patients with tuberculosis at a tertiary hospital in South Korea. *J Int Med Res* 2019; **47**: 271–80.

55 Batool R, Khan SW, Imran M, Barry Z, Ali SZ. Treatment outcomes of the drug resistant tuberculosis cases previously exposed to second line anti Tuberculosis drugs in Pakistan: A multi-center cross-sectional study. *J Pak Med Assoc* 2019; **69**: 4–10.

56 El Hamdouni M, Bourkadi JE, Benamor J, Hassar M, Cherrah Y, Ahid S. Treatment outcomes of drug resistant tuberculosis patients in Morocco: multi-centric prospective study. *BMC Infect Dis* 2019; **19**: 316.

57 Khan AH, Sulaiman SAS, Laghari M, *et al.* Treatment outcomes and risk factors of extra-pulmonary tuberculosis in patients with co-morbidities. *BMC Infect Dis* 2019; **19**: 691.

58 Tola A, Mishore KM, Ayele Y, Mekuria AN, Legese N. Treatment Outcome of Tuberculosis and Associated Factors among TB-HIV Co-Infected Patients at Public Hospitals of Harar Town, Eastern Ethiopia. A five-year retrospective study. *BMC Public Health* 2019; **19**: 1658.

59 Musaazi J, Sekaggya-Wiltshire C, Kiragga AN, *et al.* Sustained positive impact on tuberculosis treatment outcomes of TB-HIV integrated care in Uganda. *Int J Tuberc Lung Dis* 2019; **23**: 514–21.

60 Balaky STJ, Mawlood AH, Shabila NP. Survival analysis of patients with tuberculosis in Erbil, Iraqi Kurdistan region. *BMC Infect Dis* 2019; **19**: 865.

61 Aguilar JP, Arriaga MB, Rodas MN, Martins Netto E. Smoking and pulmonary tuberculosis treatment failure: a case-control study. *J Bras Pneumol* 2019; **45**: e20180359.

62 Ali MH, Alrasheedy AA, Kibuule D, Godman B, Hassali MA, Ali HMH. Assessment of multidrug-resistant tuberculosis (MDR-TB) treatment outcomes in Sudan; findings and implications. *Expert Rev Anti Infect Ther* 2019; **17**: 927–37.

63 Pedrazzoli D, Kranzer K, Thomas HL, Lalor MK. Trends and risk factors for death and excess all-cause mortality among notified tuberculosis patients in the uk: An analysis of surveillance data. *ERJ Open Res* 2019; **5**. DOI:10.1183/23120541.00125-2019.

64 Dedefo MG, Sirata MT, Ejeta BM, Wakjira GB, Fekadu G, Labata BG. Treatment outcomes of tuberculosis retreatment case and its determinants in west ethiopia. *Open Respir Med J* 2019; **13**: 58–64.

65 Pradipta IS, Van't Boveneind-Vrubleuskaya N, Akkerman OW, Alffenaar JWC, Hak E. Predictors for treatment outcomes among patients with drug-susceptible tuberculosis in the Netherlands: a retrospective cohort study. *Clin Microbiol Infect* 2019; **25**: 761.e1-761.e7.

66 Somsong W, Lawpoolsri S, Kasetjaroen Y, Manosuthi W, Kaewkungwal J. Treatment outcomes for elderly patients in Thailand with pulmonary tuberculosis. *Asian Biomed* 2019; **12**: 75–82.

67 Nguyen DT, Jenkins HE, Graviss EA. Prognostic score to predict mortality during TB treatment in TB/HIV co-infected patients. *PLoS One* 2018; **13**: e0196022.

68 Nandasena S, Senavirathna C, Munasinghe C, Wijesena C, Sucharitharathna R. Characteristics and sputum conversion of tuberculosis (TB) patients in Kalutara, Sri Lanka. *Indian J Tuberc* 2019; **66**: 76–80.

69 Prudhivi R, Challa SR, Basaveswara Rao M V, Veena G, Bhaskara Rao N, Narne HM. Assessment of success rate of directly observed treatment short-course (DOTS) in tuberculosis patients of South India. *J Young Pharm* 2019; **11**: 67–72.

70 Zhang Y, Wu L, Li F. Analysis of survival time and factors that influence HIV/ mycobacterium tuberculosis infection. *Acta Medica Mediterr* 2019; **35**: 23–7.

71 Rossetto M, Brand É M, Rodrigues RM, Serrant L, Teixeira LB. Factors associated with hospitalization and death among TB/HIV co-infected persons in Porto Alegre, Brazil. *PLoS One* 2019; **14**: e0209174.

72 Ohene SA, Bakker MI, Ojo J, Toonstra A, Awudi D, Klatser P. Extra-pulmonary tuberculosis: A retrospective study of patients in Accra, Ghana. *PLoS One* 2019; **14**: e0209650.

73 Khan I, Ahmad N, Khan S, *et al.* Evaluation of treatment outcomes and factors associated with unsuccessful outcomes in multidrug resistant tuberculosis patients in Baluchistan province of Pakistan. *J Infect Public Heal* 2019; **12**: 809–15.

74 Holmberg V, Soini H, Kivelä P, Ollgren J, Ristola M. Epidemiology and outcome of HIV patients in Finland co-infected with tuberculosis 1998-2015. *BMC Infect Dis* 2019; **19**: 264.

75 Sadykova L, Maimakov T, Berikova E, Kurakbayev K, Carr NT. Laura Sadykova, MD, PhD. 2020; : 1–14.

76 Dangeti G V, Mailankody S, Neeradi C, *et al.* Vitamin D deficiency in patients with tuberculous meningitis and its relationship with treatment outcome. *Int J Tuberc Lung Dis* 2018; **22**: 93–9.

77 Javaid A, Ahmad N, Afridi AK, *et al.* Validity of Time to Sputum Culture Conversion to Predict Cure in Patients with Multidrug-Resistant Tuberculosis: A Retrospective Single-Center Study. *Am J Trop Med Hyg* 2018; **98**: 1629–36.

78 Ambaw F, Mayston R, Hanlon C, Medhin G, Alem A. Untreated depression and tuberculosis treatment outcomes, quality of life and disability, Ethiopia. *Bull World Heal Organ* 2018; **96**: 243–55.

79 Parmar MM, Sachdeva KS, Dewan PK, *et al.* Unacceptable treatment outcomes and associated factors among India's initial cohorts of multidrug-resistant tuberculosis (MDR-TB) patients under the revised national TB control programme (2007-2011): Evidence leading to policy enhancement. *PLoS One* 2018; **13**: e0193903.

80 Adane K, Spigt M, Dinant GJ. Tuberculosis treatment outcome and predictors in northern Ethiopian prisons: a five-year retrospective analysis. *BMC Pulm Med* 2018; **18**: 37.

81 Worku S, Derbie A, Mekonnen D, Biadglegne F. Treatment outcomes of tuberculosis patients under directly observed treatment short-course at Debre Tabor General Hospital, northwest Ethiopia: nine-years retrospective study. *Infect Dis Poverty* 2018; **7**: 16.

82 Tafess K, Beyen TK, Abera A, *et al.* Treatment Outcomes of Tuberculosis at Asella Teaching Hospital, Ethiopia: Ten Years' Retrospective Aggregated Data. *Front Med* 2018; **5**: 38.

83 Javaid A, Ullah I, Masud H, *et al.* Predictors of poor treatment outcomes in multidrug-resistant tuberculosis patients: a retrospective cohort study. *Clin Microbiol Infect* 2018; **24**: 612–7.

84 Muyaya LM, Young T, Loveday M. Predictors of mortality in adults on treatment for human immunodeficiency virus-associated tuberculosis in Botswana: A retrospective cohort study. *Med* 2018; **97**: e0486.

85 Ferreira MD, Neves CPD, Souza AB, *et al.* Predictors of mortality among intensive care unit patients coinfected with tuberculosis and HIV. *J Bras Pneumol* 2018; **44**: 118–24.

86 Muluye AB, Kebamo S, Teklie T, Alemkere G. Poor treatment outcomes and its determinants among tuberculosis patients in selected health facilities in East Wollega, Western Ethiopia. *PLoS One* 2018; **13**: e0206227.

87 Tshitenge S, Ogunbanjo GA, Citeya A. A mortality review of tuberculosis and HIV co-infected patients in Mahalapye, Botswana: Does cotrimoxazole preventive therapy and/or antiretroviral therapy protect against death? *Afr J Prim Heal Care Fam Med* 2018; **10**: e1–5.

88 Evans D, Sineke T, Schnippel K, *et al.* Impact of Xpert MTB/RIF and decentralized care on linkage to care and drug-resistant tuberculosis treatment outcomes in Johannesburg, South Africa. *BMC Heal Serv Res* 2018; **18**: 973.

89 Adamu AL, Aliyu MH, Galadanci NA, *et al.* The impact of rural residence and HIV infection on poor tuberculosis treatment outcomes in a large urban hospital: a retrospective cohort analysis. *Int J Equity Heal* 2018; **17**: 4.

90 Kaplan R, Hermans S, Caldwell J, Jennings K, Bekker LG, Wood R. HIV and TB co-infection in the ART era: CD4 count distributions and TB case fatality in Cape Town. *BMC Infect Dis* 2018; **18**: 356.

91 Bulabula ANH, Nelson JA, Musafiri EM, *et al.* Prevalence, Predictors, and Successful Treatment Outcomes of Xpert MTB/RIF-identified Rifampicin-resistant Tuberculosis in Post-conflict Eastern Democratic Republic of the Congo, 2012-2017: A Retrospective Province-Wide Cohort Study. *Clin Infect Dis* 2019; **69**: 1278–87.

92 Frank M, Adamashvili N, Lomtadze N, *et al.* Long-term follow-up reveals high posttreatment mortality rate among patients with extensively drug-resistant tuberculosis in the Country of Georgia. *Open Forum Infect Dis* 2019; **6**. DOI:10.1093/ofid/ofz152.

93 Garg K, Saini V, Dhillon R, Agarwal P. Isoniazid mono-resistant tuberculosis: Time to take it seriously. *Indian J Tuberc* 2019; **66**: 247–52.

94 Lee EH, Yong SH, Leem AY, *et al.* Improved Fluoroquinolone-Resistant and Extensively Drug-Resistant Tuberculosis Treatment Outcomes. *Open Forum Infect Dis* 2019; **6**. DOI:10.1093/ofid/ofz118.

95 Crabtree-Ramírez B, Jenkins C, Jayathilake K, *et al.* HIV-related tuberculosis: Mortality risk in persons without vs. With culture-confirmed disease. *Int J Tuberc Lung Dis* 2019; **23**: 306–14.

96 Han Y, Kim SJ, Lee SH, *et al.* High blood neutrophil-lymphocyte ratio associated with poor outcomes in miliary tuberculosis. *J Thorac Dis* 2018; **10**: 339–46.

97 Ejeta E, Beyene G, Balay G, Bonsa Z, Abebe G. Factors associated with unsuccessful treatment outcome in tuberculosis patients among refugees and their surrounding communities in Gambella Regional State, Ethiopia. *PLoS One* 2018; **13**: e0205468.

98 Diallo A, Dahourou DL, Dah TTE, Tassembedo S, Sawadogo R, Meda N. Factors associated with tuberculosis treatment failure in the Central East Health region of Burkina Faso. *Pan Afr Med J* 2018; **30**: 293.

99 Melese A, Zeleke B. Factors associated with poor treatment outcome of tuberculosis in Debre Tabor, northwest Ethiopia. *BMC Res Notes* 2018; **11**: 25.

100 Viana PVS, Redner P, Ramos JP. Factors associated with loss to follow-up and death in cases of drug-resistant tuberculosis (DR-TB) treated at a reference center in Rio de Janeiro, Brazil. *Cad Saude Publica* 2018; **34**: e00048217.

101 Wu W, Yang M, Xu M, *et al.* Diagnostic delay and mortality of active tuberculosis in patients after kidney transplantation in a tertiary care hospital in China. *PLoS One* 2018; **13**: e0195695.

102 Feng JY, Pan SW, Huang SF, Chen YY, Lin YY, Su WJ. Depressed Gamma Interferon Responses and Treatment Outcomes in Tuberculosis Patients: a Prospective Cohort Study. *J Clin Microbiol* 2018; **56**. DOI:10.1128/jcm.00664-18.

103 Sekaggya-Wiltshire C, von Braun A, Lamorde M, *et al.* Delayed Sputum Culture Conversion in Tuberculosis-Human Immunodeficiency Virus-Coinfected Patients With Low Isoniazid and Rifampicin Concentrations. *Clin Infect Dis* 2018; **67**: 708–16.

104 Thai PVK, Ha DTM, Hanh NT, *et al.* Bacterial risk factors for treatment failure and relapse among patients with isoniazid resistant tuberculosis. *BMC Infect Dis* 2018; **18**: 112.

105 Shimazaki T, Taniguchi T, Saludar NRD, *et al.* Bacterial co-infection and early mortality among pulmonary tuberculosis patients in Manila, The Philippines. *Int J Tuberc Lung Dis* 2018; **22**: 65–72.

106 Nguyen CH, Pascopella L, Barry PM. Association between diabetes mellitus and mortality among patients with tuberculosis in California, 2010-2014. *Int J Tuberc Lung Dis* 2018; **22**: 1269–76.

107 Mishkin K, Alaei K, Alikeyeva E, Paynter C, Aringazina A, Alaei A. Association between antiretroviral therapy and antitubercular drug resistance in TB treatment outcome among Kazakh TB/HIV co-infected patients. *J Glob Antimicrob Resist* 2018; **14**: 104–8.

108 Azeez A, Ndege J, Mutambayi R. Associated factors with unsuccessful tuberculosis treatment outcomes among tuberculosis/HIV coinfected patients with drug-resistant tuberculosis. *Int J Mycobacteriol* 2018; **7**: 347–54.

109 Velavan A, Purty AJ, Shringarpure K, *et al.* Tuberculosis retreatment outcomes and associated factors: A mixed-methods study from Puducherry, India. *Public Heal Action* 2018; **8**: 187–93.

110 Yu MC, Chiang CY, Lee JJ, *et al.* Treatment outcomes of multidrug-resistant tuberculosis in Taiwan: Tackling loss to follow-up. *Clin Infect Dis* 2018; **67**: 202–10.

111 Lin CB, Sun HC, Chiang CY, *et al.* Treatment outcomes for multidrug-resistant tuberculosis in Eastern Taiwan. *Tzu Chi Med J* 2018; **31**: 35–9.

112 Lv L, Li T, Xu K, *et al.* Sputum bacteriology conversion and treatment outcome of patients with multidrug-resistant tuberculosis. *Infect Drug Resist* 2018; **11**: 147–54.

113 Kuehne A, Hauer B, Brodhun B, Haas W, Fiebig L. Screening and prevention of infectious diseases in newly arrived migrants. Find and treat or find and lose? Tuberculosis treatment outcomes among screened newly arrived asylum seekers in Germany 2002 to 2014. *Eurosurveillance* 2018; **23**. DOI:10.2807/1560-7917.ES.2018.23.11.17-00042.

114 Hameed S, Zuberi FF, Hussain S, Ali SK. Risk factors for mortality among inpatients with smear positive pulmonary tuberculosis. *Pakistan J Med Sci* 2019; **35**: 1361–5.

115 Wickett E, Peralta-Santos A, Beste J, *et al.* Treatment outcomes of TB-infected individuals attending public sector primary care clinics in rural Liberia from 2015 to 2017: a retrospective cohort study. *Trop Med Int Heal* 2018; **23**: 549–57.

116 Zhang L, Meng Q, Chen S, *et al.* Treatment outcomes of multidrug-resistant tuberculosis patients in Zhejiang, China, 2009-2013. *Clin Microbiol Infect* 2018; **24**: 381–8.

117 Cornejo Garcia JG, Alarcón Guizado VA, Mendoza Ticona A, Alarcon E, Heldal E, Moore DAJ. Treatment outcomes for isoniazid-monoresistant tuberculosis in Peru, 2012-2014. *PLoS One* 2018; **13**: e0206658.

118 Mok J, An D, Kim S, Lee M, Kim C, Son H. Treatment outcomes and factors affecting treatment outcomes of new patients with tuberculosis in Busan, South Korea: a retrospective study of a citywide registry, 2014-2015. *BMC Infect Dis* 2018; **18**: 655.

119 Trébucq A, Schwoebel V, Kashongwe Z, *et al.* Treatment outcome with a short multidrug-resistant tuberculosis regimen in nine African countries. *Int J Tuberc Lung Dis* 2018; **22**: 17–25.

120 Weldegebreal F, Mitiku H, Teklemariam Z. Treatment outcome of tuberculosis among Human Immunodeficiency Virus positive patients in Eastern Ethiopia: a retrospective study. *Pan Afr Med J* 2018; **30**: 32.

121 Ige OM, Oladokun RE. Time to sputum culture conversion and treatment outcome among the first cohort of

multidrug resistant tuberculosis patients in a high burden country. *Indian J Tuberc* 2018; **65**: 322–8.

122 Chingonzoh R, Manesen MR, Madlavu MJ, *et al.* Risk factors for mortality among adults registered on the routine drug resistant tuberculosis reporting database in the Eastern Cape Province, South Africa, 2011 to 2013. *PLoS One* 2018; **13**: e0202469.

123 Ji YJ, Liang PP, Shen JY, *et al.* Risk factors affecting the mortality of HIV-infected patients with pulmonary tuberculosis in the cART era: a retrospective cohort study in China. *Infect Dis Poverty* 2018; **7**: 25.

124 Velayutham B, Chadha VK, Singla N, *et al.* Recurrence of tuberculosis among newly diagnosed sputum positive pulmonary tuberculosis patients treated under the Revised National Tuberculosis Control Programme, India: A multi-centric prospective study. *PLoS One* 2018; **13**: e0200150.

125 Obregón G, Zevallos K, Alarcón V, *et al.* Rapid drug susceptibility testing and treatment outcomes for multidrug-resistant tuberculosis in Peru. *Int J Tuberc Lung Dis* 2018; **22**: 1350–7.

126 Alarcón V, Alarcón-Arrascue E, Mendoza-Ticona A, *et al.* Programmatic management of patients with pre-extensively drug-resistant tuberculosis in Peru, 2011-2014. *Int J Tuberc Lung Dis* 2018; **22**: 1220–6.

127 Nguyen DT, Graviss EA. Development and validation of a prognostic score to predict tuberculosis mortality. *J Infect* 2018; **77**: 283–90.

128 Thao LTP, Heemskerk AD, Geskus RB, *et al.* Prognostic Models for 9-Month Mortality in Tuberculous Meningitis. *Clin Infect Dis* 2018; **66**: 523–32.

129 Fan YM, Ding SP, Bao ZJ, *et al.* Prognostic factors for treatment success in patients with multidrug-resistant tuberculosis in China. *Int J Tuberc Lung Dis* 2018; **22**: 300–5.

130 Kibuule D, Verbeeck RK, Nunurai R, *et al.* Predictors of tuberculosis treatment success under the DOTS program in Namibia. *Expert Rev Respir Med* 2018; **12**: 979–87.

131 Pizzol D, Veronese N, Marotta C, *et al.* Predictors of therapy failure in newly diagnosed pulmonary tuberculosis cases in Beira, Mozambique. *BMC Res Notes* 2018; **11**: 99.

132 Kirirabwa NS, Kimuli D, DeJene S, *et al.* Response to anti-tuberculosis treatment by people over age 60 in Kampala, Uganda. *PLoS One* 2018; **13**. DOI:10.1371/journal.pone.0208390.

133 Wakamatsu K, Nagata N, Kumazoe H, *et al.* Prognostic factors in patients with miliary tuberculosis. *J Clin Tuberc Other Mycobact Dis* 2018; **12**: 66–72.

134 Silva DI, Ceccato MDGB, Silveira MR, *et al.* Predictors of mortality among individuals with tuberculosis and human immunodeficiency virus coinfection at a reference center in southeastern Brazil: A retrospective cohort study. *J Young Pharm* 2018; **10**: 476–80.

135 Aibana O, Slavuckij A, Bachmaha M, *et al.* Patient predictors of poor drug sensitive tuberculosis treatment outcomes in Kyiv Oblast, Ukraine [version 2; peer review: 2 approved, 1 approved with reservations]. *F1000Research* 2018; **6**. DOI:10.12688/f1000research.12687.2.

136 Singhi L, Sagili KD, Sharath BN, *et al.* Non-response to first-line anti-tuberculosis treatment in Sikkim, India: A risk-factor analysis study. *Public Heal Action* 2018; **8**: 162–8.

137 Tan W, Soodeen-Lalloo AK, Chu Y, *et al.* Sex influences the association between haemostasis and the extent of lung lesions in tuberculosis. *Biol Sex Differ* 2018; **9**: 44.

138 Kumar R, Ahirwar RK, Dave L, *et al.* TREATMENT OUTCOME AND EFFICACY OF ANTI-TUBERCULOSIS TREATMENT IN TUBERCULOSIS PATIENTS PUT ON DOTS IN RNTCP IN CENTRAL INDIA. *J Evol Med Dent Sci* 2018; **7**: 1840–4.

139 Engelbrecht MC, Kigozi NG, Chikobvu P, Botha S, van Rensburg HCJ. Unsuccessful TB treatment outcomes

with a focus on HIV co-infected cases: a cross-sectional retrospective record review in a high-burdened province of South Africa. *BMC Heal Serv Res* 2017; **17**: 470.

140 Suryawanshi SL, Shewade HD, Nagaraja SB, Nair SA, Parmar M. Unfavourable outcomes among patients with MDR-TB on the standard 24-month regimen in Maharashtra, India. *Public Heal Action* 2017; **7**: 116–22.

141 Mattila T, Heliövaara M, Rissanen H, Knekt P, Puukka P, Vasankari T. Tuberculosis, Airway Obstruction and Mortality in a Finnish Population. *COPD* 2017; **14**: 143–9.

142 Cardoso MA, do Brasil P, Schmaltz CAS, Sant'Anna FM, Rolla VC. Tuberculosis Treatment Outcomes and Factors Associated with Each of Them in a Cohort Followed Up between 2010 and 2014. *Biomed Res Int* 2017; **2017**: 3974651.

143 Tatar D, Senol G, Kirakli C, Edipoglu O, Cimen P. Contributing factors to mortality rates of pulmonary tuberculosis in intensive care units. *J Chin Med Assoc* 2018; **81**: 605–10.

144 Alene KA, Viney K, Gray DJ, McBryde ES, Xu Z, Clements ACA. Development of a risk score for prediction of poor treatment outcomes among patients with multidrug-resistant tuberculosis. *PLoS One* 2020; **15**: e0227100.

145 Nagu TJ, Aboud S, Mwiru R, *et al.* Tuberculosis associated mortality in a prospective cohort in Sub Saharan Africa: Association with HIV and antiretroviral therapy. *Int J Infect Dis* 2017; **56**: 39–44.

146 Nagu T, Ray R, Munseri P, *et al.* Tuberculosis among the elderly in Tanzania: disease presentation and initial response to treatment. *Int J Tuberc Lung Dis* 2017; **21**: 1251–7.

147 Lee H, Jeong BH, Park HY, *et al.* Treatment Outcomes with Fluoroquinolone-Containing Regimens for Isoniazid-Resistant Pulmonary Tuberculosis. *Antimicrob Agents Chemother* 2016; **60**: 471–7.

148 Bastos ML, Cosme LB, Fregona G, *et al.* Treatment outcomes of MDR-tuberculosis patients in Brazil: a retrospective cohort analysis. *BMC Infect Dis* 2017; **17**: 718.

149 Alene KA, Viney K, McBryde ES, Tsegaye AT, Clements AC. Treatment outcomes in patients with multidrug-resistant tuberculosis in north-west Ethiopia. *Trop Med Int Heal* 2017; **22**: 351–62.

150 Ahmad T, Khan M, Khan MM, Ejeta E, Karami M, Ohia C. Treatment outcome of tuberculosis patients under directly observed treatment short course and its determinants in Shangla, Khyber-Pakhtunkhwa, Pakistan: A retrospective study. *Int J Mycobacteriol* 2017; **6**: 360–4.

151 Schechter MC, Bizune D, Kagei M, *et al.* Time to Sputum Culture Conversion and Treatment Outcomes Among Patients with Isoniazid-Resistant Tuberculosis in Atlanta, Georgia. *Clin Infect Dis* 2017; **65**: 1862–71.

152 Sinshaw Y, Alemu S, Fekadu A, Gizachew M. Successful TB treatment outcome and its associated factors among TB/HIV co-infected patients attending Gondar University Referral Hospital, Northwest Ethiopia: an institution based cross-sectional study. *BMC Infect Dis* 2017; **17**: 132.

153 Aibana O, Bachmaha M, Krasiuk V, *et al.* Risk factors for poor multidrug-resistant tuberculosis treatment outcomes in Kyiv Oblast, Ukraine. *BMC Infect Dis* 2017; **17**: 129.

154 Loh WJ, Yu Y, Loo CM, Low SY. Factors associated with mortality among patients with active pulmonary tuberculosis requiring intensive care. *Singapore Med J* 2017; **58**: 656–9.

155 Nagu TJ, Aboud S, Matee MI, Maeurer MJ, Fawzi WW, Mugusi F. Effects of isoniazid resistance on TB treatment outcomes under programmatic conditions in a high-TB and -HIV setting: a prospective multicentre study. *J Antimicrob Chemother* 2017; **72**: 876–81.

156 Getnet F, Sileshi H, Seifu W, Yirga S, Alemu AS. Do retreatment tuberculosis patients need special treatment response follow-up beyond the standard regimen? Finding of five-year retrospective study in pastoralist setting. *BMC Infect Dis* 2017; **17**: 762.

157 Ko PY, Lin SD, Hsieh MC, Chen YC. Diabetes mellitus increased all-cause mortality rate among newly-diagnosed tuberculosis patients in an Asian population: A nationwide population-based study. *Diabetes Res Clin Pr* 2017; **133**: 115–23.

158 Yen YF, Feng JY, Pan SW, Chuang PH, Su VY, Su WJ. Determinants of mortality in elderly patients with tuberculosis: a population-based follow-up study. *Epidemiol Infect* 2017; **145**: 1374–81.

159 Raberahona M, Rakotoarivelo RA, Razafinambinintsoa T, Andrianasolo RL, Randria MJ. Clinical Features and Outcome in Adult Cases of Tuberculous Meningitis in Tertiary Care Hospital in Antananarivo, Madagascar. *Biomed Res Int* 2017; **2017**: 9316589.

160 Prado TN, Rajan J V, Miranda AE, *et al.* Clinical and epidemiological characteristics associated with unfavorable tuberculosis treatment outcomes in TB-HIV co-infected patients in Brazil: a hierarchical polytomous analysis. *Braz J Infect Dis* 2017; **21**: 162–70.

161 Sam S, Shapiro AE, Sok T, *et al.* Initiation, scale-up and outcomes of the Cambodian National MDR-TB programme 2006-2016: Hospital and community-based treatment through an NGO-NTP partnership. *BMJ Open Respir Res* 2018; **5**. DOI:10.1136/bmjresp-2017-000256.

162 Kuhlin J, Smith C, Khaemraev A, *et al.* Impact of pyrazinamide resistance on multidrug-resistant tuberculosis in Karakalpakstan, Uzbekistan. *Int J Tuberc Lung Dis* 2018; **22**: 544–50.

163 Ndjeka N, Schnippel K, Master I, *et al.* High treatment success rate for multidrug-resistant and extensively drug-resistant tuberculosis using a bedaquiline-containing treatment regimen. *Eur Respir J* 2018; **52**. DOI:10.1183/13993003.01528-2018.

164 Sauer CM, Sasson D, Paik KE, *et al.* Feature selection and prediction of treatment failure in tuberculosis. *PLoS One* 2018; **13**. DOI:10.1371/journal.pone.0207491.

165 Diktanas S, Vasiliauskiene E, Polubenko K, *et al.* Factors associated with persistent sputum positivity at the end of the second month of tuberculosis treatment in Lithuania. *Tuberc Respir Dis (Seoul)* 2018; **81**: 233–40.

166 Nguyen CH, Pascopella L, Barry PM. Association between diabetes mellitus and mortality among patients with tuberculosis in California, 2010-2014. *Int J Tuberc Lung Dis* 2018; **22**: 1269–76.

167 Mundra A, Deshmukh P, Dawale A. Determinants of adverse treatment outcomes among patients treated under Revised National Tuberculosis Control Program in Wardha, India: Case–control study. *Med J Armed Forces India* 2018; **74**: 241–9.

168 Asres A, Jerene D, Deressa W. Delays to treatment initiation is associated with tuberculosis treatment outcomes among patients on directly observed treatment short course in Southwest Ethiopia: A follow-up study. *BMC Pulm Med* 2018; **18**. DOI:10.1186/s12890-018-0628-2.

169 Gaborit BJ, Revest M, Roblot F, *et al.* Characteristics and outcome of multidrug-resistant tuberculosis in a low-incidence area. *Med Mal Infect* 2018; **48**: 457–64.

170 Jackson C, Stagg HR, Doshi A, *et al.* Tuberculosis treatment outcomes among disadvantaged patients in India. *Public Heal Action* 2017; **7**: 134–40.

171 Piparva KG. Treatment outcome of tuberculosis patients on dots therapy for category 1 and 2 at district tuberculosis centre. *Int J Pharm Sci Res* 2017; **8**: 207–12.

172 Wang XM, Yin SH, Du J, *et al.* Risk factors for the treatment outcome of retreated pulmonary tuberculosis patients in China: An optimized prediction model. *Epidemiol Infect* 2017; **145**: 1805–14.

173 Wondale B, Medihn G, Teklu T, Mersha W, Tamirat M, Ameni G. A retrospective study on tuberculosis treatment outcomes at Jinka General Hospital, southern Ethiopia. *BMC Res Notes* 2017; **10**: 680.

174 Kefale AT, Anagaw YK. Outcome of tuberculosis treatment and its predictors among HIV infected patients in

southwest Ethiopia. *Int J Gen Med* 2017; **10**: 161–9.

175 Gunda DW, Nkandala I, Kavishe GA, Kilonzo SB, Kabangila R, Mpondo BC. Prevalence and Risk Factors of Delayed Sputum Conversion among Patients Treated for Smear Positive PTB in Northwestern Rural Tanzania: A Retrospective Cohort Study. *J Trop Med* 2017; **2017**: 5352906.

176 Nair D, Velayutham B, Kannan T, *et al.* Predictors of unfavourable treatment outcome in patients with multidrug-resistant tuberculosis in India. *Public Heal Action* 2017; **7**: 32–8.

177 Kapata N, Grobusch MP, Chongwe G, *et al.* Outcomes of multidrug-resistant tuberculosis in Zambia: a cohort analysis. *Infection* 2017; **45**: 831–9.

178 Huerga H, Bastard M, Kamene M, *et al.* Outcomes from the first multidrug-resistant tuberculosis programme in Kenya. *Int J Tuberc Lung Dis* 2017; **21**: 314–9.

179 Mehta S. Ocular Inflammatory Disease as a Predictor for In-Hospital Mortality in Patients Hospitalized with Disseminated Tuberculosis. *Cureus* 2017; **9**: e956.

180 Mert A, Arslan F, Kuyucu T, *et al.* Miliary tuberculosis: Epidemiologicaland clinical analysis of large-case series from moderate to low tuberculosis endemic Country. *Med* 2017; **96**: e5875.

181 van der Heijden YF, Karim F, Mufamadi G, *et al.* Isoniazid-monoresistant tuberculosis is associated with poor treatment outcomes in Durban, South Africa. *Int J Tuberc Lung Dis* 2017; **21**: 670–6.

182 Georghiou SB, Seifert M, Catanzaro DG, Garfein RS, Rodwell TC. Increased Tuberculosis Patient Mortality Associated with Mycobacterium tuberculosis Mutations Conferring Resistance to Second-Line Antituberculous Drugs. *J Clin Microbiol* 2017; **55**: 1928–37.

183 Seifert M, Georghiou SB, Garfein RS, Catanzaro D, Rodwell TC. Impact of Fluoroquinolone Use on Mortality Among a Cohort of Patients With Suspected Drug-Resistant Tuberculosis. *Clin Infect Dis* 2017; **65**: 772–8.

184 Shin SS, Modongo C, Boyd R, *et al.* High Treatment Success Rates Among HIV-Infected Multidrug-Resistant Tuberculosis Patients After Expansion of Antiretroviral Therapy in Botswana, 2006-2013. *J Acquir Immune Defic Syndr* 2017; **74**: 65–71.

185 Thu MK, Kumar AM V, Soe KT, *et al.* High treatment success rate among multidrug-resistant tuberculosis patients in Myanmar, 2012-2014: a retrospective cohort study. *Trans R Soc Trop Med Hyg* 2017; **111**: 410–7.

186 Janssen S, Schutz C, Ward AM, *et al.* Hemostatic Changes Associated With Increased Mortality Rates in Hospitalized Patients With HIV-Associated Tuberculosis: A Prospective Cohort Study. *J Infect Dis* 2017; **215**: 247–58.

187 Teklu AM, Nega A, Mamuye AT, *et al.* Factors Associated with Mortality of TB/HIV Co-infected Patients in Ethiopia. *Ethiop J Heal Sci* 2017; **27**: 29–38.

188 Dale K, Tay E, Trauer JM, Trevan P, Denholm J. Gender differences in tuberculosis diagnosis, treatment and outcomes in Victoria, Australia, 2002–2015. *Int J Tuberc Lung Dis* 2017; **21**: 1264–71.

189 Ramachandran G, Kupparam HKA, Vedhachalam C, *et al.* Factors influencing tuberculosis treatment outcome in adult patients treated with thrice-weekly regimens in India. *Antimicrob Agents Chemother* 2017; **61**. DOI:10.1128/AAC.02464-16.

190 Faisal M, Alauddin CABM, Md. Imdadul H, Moniruddin C. An evaluation of treatment outcome in tuberculosis directly observed treatment short course facilities in Jigawa state, Nigeria (2010–2014). *J Heal Transl Med* 2017; **20**: 1–7.

191 El-Shabrawy M, El-Shafei DA. Evaluation of treatment failure outcome and its predictors among pulmonary tuberculosis patients in Sharkia Governorate, 2013–2014. *Egypt J Chest Dis Tuberc* 2017; **66**: 145–52.

192 Jaber AAS, Khan AH, Sulaiman SAS. Evaluating treatment outcomes and durations among cases of smear-positive pulmonary tuberculosis in Yemen: A prospective follow-up study. *J Pharm Policy Pract* 2017; **10**. DOI:10.1186/s40545-017-0124-8.

193 Masjedi MR, Hosseini M, Aryanpur M, *et al.* The effects of smoking on treatment outcome in patients newly diagnosed with pulmonary tuberculosis. *Int J Tuberc Lung Dis* 2017; **21**: 351–6.

194 Heysell SK, Ogarkov OB, Zhdanova S, *et al.* Undertreated HIV and drug-resistant tuberculosis at a referral hospital in Irkutsk, Siberia. *Int J Tuberc Lung Dis* 2016; **20**: 187–92.

195 Lee H, Ahn S, Hwang NY, *et al.* Treatment outcomes of rifabutin-containing regimens for rifabutin-sensitive multidrug-resistant pulmonary tuberculosis. *Int J Infect Dis* 2017; **65**: 135–41.

196 Parchure R, Kulkarni V, Gangakhedkar R, Swaminathan S. Treatment outcomes of daily anti-tuberculosis treatment in HIV-infected patients seeking care at a private clinic in India. *Int J Tuberc Lung Dis* 2016; **20**: 1348–53.

197 Patel S V, Nimavat KB, Alpesh PB, *et al.* Treatment outcome among cases of multidrug-resistant tuberculosis (MDR TB) in Western India: A prospective study. *J Infect Public Heal* 2016; **9**: 478–84.

198 Dovonou AC, Kpangon AA, Amidou SA, *et al.* Risk factors for pulmonary tuberculosis treatment failure in rural settings in Benin, West Africa: A cohort study. *African J Respir Med* 2017; **12**: 8–11.

199 Gebreegziabher SB, Bjune GA, Yimer SA. Total Delay Is Associated with Unfavorable Treatment Outcome among Pulmonary Tuberculosis Patients in West Gojjam Zone, Northwest Ethiopia: A Prospective Cohort Study. *PLoS One* 2016; **11**: e0159579.

200 Balabanova Y, Ignatyeva O, Fiebig L, *et al.* Survival of patients with multidrug-resistant TB in Eastern Europe: what makes a difference? *Thorax* 2016; **71**: 854–61.

201 Mlotshwa M, Abraham N, Beery M, *et al.* Risk factors for tuberculosis smear non-conversion in Eden district, Western Cape, South Africa, 2007-2013: a retrospective cohort study. *BMC Infect Dis* 2016; **16**: 365.

202 Snyder RE, Marlow MA, Phuphanich ME, Riley LW, Maciel EL. Risk factors for differential outcome following directly observed treatment (DOT) of slum and non-slum tuberculosis patients: a retrospective cohort study. *BMC Infect Dis* 2016; **16**: 494.

203 Mbatchou Ngahane BH, Dahirou F, Tchieche C, *et al.* Clinical characteristics and outcomes of tuberculosis in Douala, Cameroon: a 7-year retrospective cohort study. *Int J Tuberc Lung Dis* 2016; **20**: 1609–14.

204 Ade S, Adjibodé O, Wachinou P, *et al.* Characteristics and Treatment Outcomes of Retreatment Tuberculosis Patients in Benin. *Tuberc Res Treat* 2016; **2016**: 1468631.

205 Cabrera-Gaytán DA, Niebla-Fuentes MD, Padilla-Velázquez R, *et al.* Association of Pulmonary Tuberculosis and HIV in the Mexican Institute of Social Security, 2006-2014. *PLoS One* 2016; **11**: e0168559.

206 Yen YF, Chuang PH, Yen MY, *et al.* Association of Body Mass Index With Tuberculosis Mortality: A Population-Based Follow-Up Study. *Med* 2016; **95**: e2300.

207 Nagai K, Horita N, Sato T, Yamamoto M, Nagakura H, Kaneko T. Age, Dehydration, Respiratory Failure, Orientation Disturbance, and Blood Pressure Score Predicts In-hospital Mortality in HIV-negative Non-multidrug-resistant Smear-positive Pulmonary Tuberculosis in Japan. *Sci Rep* 2016; **6**: 21610.

208 Asres A, Jerene D, Deressa W. Tuberculosis treatment outcomes of six and eight month treatment regimens in districts of Southwestern Ethiopia: A comparative cross-sectional study. *BMC Infect Dis* 2016; **16**. DOI:10.1186/s12879-016-1917-0.

209 Zenebe T, Tefera E. Tuberculosis treatment outcome and associated factors among smear-positive pulmonary tuberculosis patients in Afar, Eastern Ethiopia: a retrospective study. *Brazilian J Infect Dis* 2016; **20**: 635–6.

210 Gebreegziabher SB, Bjune GA, Yimer SA. Total Delay Is Associated with Unfavorable Treatment Outcome among Pulmonary Tuberculosis Patients in West Gojjam Zone, Northwest Ethiopia: A Prospective Cohort Study. *PLoS One* 2016; **11**: e0159579.

211 Khazaei S, Hassanzadeh J, Rezaeian S, *et al.* Treatment outcome of new smear positive pulmonary tuberculosis patients in Hamadan, Iran: A registry-based cross-sectional study. *Egypt J Chest Dis Tuberc* 2016; **65**: 825–30.

212 Scott C, Cavanaugh JS, Silk BJ, *et al.* Comparison of sputum-culture conversion for Mycobacterium bovis and M. Tuberculosis. *Emerg Infect Dis* 2017; **23**: 456–62.

213 Gunda DW, Kilonzo SB, Bulegesi SM, Mpondo BCT, Shao ER. Risk factors for mortality among tuberculosis patients on treatment at Bugando Medical Centre in North-Western Tanzania: A retrospective cross-sectional study. *Tanzan J Health Res* 2016; **18**. DOI:10.4314/thrb.v18i4.1.

214 Mekonnen D, Derbie A, Mekonnen H, Zenebe Y. Profile and treatment outcomes of patients with tuberculosis in Northeastern Ethiopia: A cross sectional study. *Afr Health Sci* 2016; **16**: 663–70.

215 Boaz MM, Kalluvya S, Downs JA, Mpondo BCT, Mshana SE. Pattern, Clinical Characteristics, and Outcome of Meningitis among HIV-Infected Adults Admitted in a Tertiary Hospital in North Western Tanzania: A Cross-Sectional Study. *J Trop Med* 2016; **2016**. DOI:10.1155/2016/6573672.

216 Schnippel K, Shearer K, Evans D, Berhanu R, Dlamini S, Ndjeka N. Predictors of mortality and treatment success during treatment for rifampicin-resistant tuberculosis within the South African National TB Programme, 2009 to 2011: a cohort analysis of the national case register. *Int J Infect Dis* 2015; **39**: 89–94.

217 Kerkhoff AD, Wood R, Cobelens FG, Gupta-Wright A, Bekker LG, Lawn SD. The predictive value of current haemoglobin levels for incident tuberculosis and/or mortality during long-term antiretroviral therapy in South Africa: a cohort study. *BMC Med* 2015; **13**: 70.

218 van Lettow M, Bedell R, Maosa S, *et al.* Outcomes and Diagnostic Processes in Outpatients with Presumptive Tuberculosis in Zomba District, Malawi. *PLoS One* 2015; **10**: e0141414.

219 Ejeta E, Chala M, Arega G, *et al.* Outcome of tuberculosis patients under directly observed short course treatment in western Ethiopia. *J Infect Dev Ctries* 2015; **9**: 752–9.

220 Mpagama SG, Lekule IA, Mbuya AW, Kisonga RM, Heysell SK. The Influence of Mining and Human Immunodeficiency Virus Infection Among Patients Admitted for Retreatment of Tuberculosis in Northern Tanzania. *Am J Trop Med Hyg* 2015; **93**: 212–5.

221 Pepper DJ, Schomaker M, Wilkinson RJ, de Azevedo V, Maartens G. Independent predictors of tuberculosis mortality in a high HIV prevalence setting: a retrospective cohort study. *AIDS Res Ther* 2015; **12**: 35.

222 Trajman A, Durovni B, Saraceni V, *et al.* Impact on Patients' Treatment Outcomes of XpertMTB/RIF Implementation for the Diagnosis of Tuberculosis: Follow-Up of a Stepped-Wedge Randomized Clinical Trial. *PLoS One* 2015; **10**: e0123252.

223 Wejse C, Patsche CB, Kühle A, *et al.* Impact of HIV-1, HIV-2, and HIV-1+2 dual infection on the outcome of tuberculosis. *Int J Infect Dis* 2015; **32**: 128–34.

224 Nglazi MD, Bekker LG, Wood R, Kaplan R. The impact of HIV status and antiretroviral treatment on TB treatment outcomes of new tuberculosis patients attending co-located TB and ART services in South Africa: a retrospective cohort study. *BMC Infect Dis* 2015; **15**: 536.

225 Acuña-Villaorduña C, Ayakaka I, Dryden-Peterson S, *et al.* High mortality associated with retreatment of tuberculosis in a clinic in Kampala, Uganda: a retrospective study. *Am J Trop Med Hyg* 2015; **93**: 73–5.

226 Jung IY, Song YG, Choi JY, *et al.* Predictive factors for unfavorable outcomes of tuberculous pericarditis in human immunodeficiency virus-uninfected patients in an intermediate tuberculosis burden country. *BMC*

*Infect Dis* 2016; **16**: 719.

227 Bastos HN, Osório NS, Castro AG, *et al.* A Prediction Rule to Stratify Mortality Risk of Patients with Pulmonary Tuberculosis. *PLoS One* 2016; **11**: e0162797.

228 García-Basteiro AL, Respeito D, Augusto OJ, *et al.* Poor tuberculosis treatment outcomes in Southern Mozambique (2011-2012). *BMC Infect Dis* 2016; **16**: 214.

229 Dale K, Tay E, Trevan P, Denholm JT. Mortality among tuberculosis cases in Victoria, 2002-2013: case fatality and factors associated with death. *Int J Tuberc Lung Dis* 2016; **20**: 515–23.

230 Shuldiner J, Leventhal A, Chemtob D, Mor Z. Mortality after anti-tuberculosis treatment completion: results of long-term follow-up. *Int J Tuberc Lung Dis* 2016; **20**: 43–8.

231 Báez-Saldaña R, Delgado-Sánchez G, García-García L, *et al.* Isoniazid Mono-Resistant Tuberculosis: Impact on Treatment Outcome and Survival of Pulmonary Tuberculosis Patients in Southern Mexico 1995-2010. *PLoS One* 2016; **11**: e0168955.

232 Kaplan R, Caldwell J, Hermans S, *et al.* An integrated community TB-HIV adherence model provides an alternative to DOT for tuberculosis patients in Cape Town. *Int J Tuberc Lung Dis* 2016; **20**: 1185–91.

233 Gebremariam G, Asmamaw G, Hussen M, *et al.* Impact of HIV Status on Treatment Outcome of Tuberculosis Patients Registered at Arsi Negele Health Center, Southern Ethiopia: A Six Year Retrospective Study. *PLoS One* 2016; **11**: e0153239.

234 Lee JY, Lee HJ, Kim YK, *et al.* Impact of Fluoroquinolone Exposure Prior to Tuberculosis Diagnosis on Clinical Outcomes in Immunocompromised Patients. *Antimicrob Agents Chemother* 2016; **60**: 4005–12.

235 Ranzani OT, Carvalho CR, Waldman EA, Rodrigues LC. The impact of being homeless on the unsuccessful outcome of treatment of pulmonary TB in São Paulo State, Brazil. *BMC Med* 2016; **14**: 41.

236 Mhimbira F, Hella J, Maroa T, *et al.* Home-Based and Facility-Based Directly Observed Therapy of Tuberculosis Treatment under Programmatic Conditions in Urban Tanzania. *PLoS One* 2016; **11**: e0161171.

237 Mohammadzadeh Kh A, Ghayoomi A, Maghsoudloo D. Evaluation of factors associated with failure of tuberculosis treatment under DOTS in northern Islamic Republic of Iran. *East Mediterr Heal J* 2016; **22**: 87–94.

238 Yang JY, Han M, Koh Y, *et al.* Effects of Corticosteroids on Critically Ill Pulmonary Tuberculosis Patients With Acute Respiratory Failure: A Propensity Analysis of Mortality. *Clin Infect Dis* 2016; **63**: 1449–55.

239 Berhanu R, Schnippel K, Mohr E, *et al.* Early Outcomes Of Decentralized Care for Rifampicin-Resistant Tuberculosis in Johannesburg, South Africa: An Observational Cohort Study. *PLoS One* 2016; **11**: e0164974.

240 Dos Santos Feltrin AF, Vendramini SH, Neto FC, *et al.* Death in patients with tuberculosis and diabetes: Associated factors. *Diabetes Res Clin Pr* 2016; **120**: 111–6.

241 Prado Junior JC, Virgilio TC, Medronho Rde A. Cure rates for tuberculosis in the municipality of Rio de Janeiro, Brazil, in 2012 compared with coverage by, and time of establishment of, Family Health units, and socio-economic and demographic factors. *Cien Saude Colet* 2016; **21**: 1491–8.

242 Behnaz F, Mohammadzadeh M, Mohammadzade G. Five-year assessment of time of sputum smears conversion and outcome and risk factors of tuberculosis patients in central iran. *Tuberc Res Treat* 2015; **2015**: 609083.

243 Milanov V, Falzon D, Zamfirova M, *et al.* Factors associated with treatment success and death in cases with multidrug-resistant tuberculosis in Bulgaria, 2009-2010. *Int J Mycobacteriol* 2015; **4**: 131–7.

244 Shahrezaei M, Maracy MR, Farid F. Factors Affecting Mortality and Treatment Completion of Tuberculosis Patients in Isfahan Province from 2006 to 2011. *Int J Prev Med* 2015; **6**: 91.

245 Bigna JJ, Noubiap JJ, Agbor AA, *et al.* Early Mortality during Initial Treatment of Tuberculosis in Patients Co-Infected with HIV at the Yaoundé Central Hospital, Cameroon: An 8-Year Retrospective Cohort Study (2006-2013). *PLoS One* 2015; **10**: e0132394.

246 Lin YS, Yen YF. Determinants of mortality before start of and during tuberculosis treatment among elderly patients: a population-based retrospective cohort study. *Age Ageing* 2015; **44**: 490–6.

247 Djouma FN, Noubom M, Ateudjieu J, Donfack H. Delay in sputum smear conversion and outcomes of smear-positive tuberculosis patients: a retrospective cohort study in Bafoussam, Cameroon. *BMC Infect Dis* 2015; **15**: 139.

248 Kwak N, Kim HR, Yoo CG, Kim YW, Han SK, Yim JJ. Changes in treatment outcomes of multidrug-resistant tuberculosis. *Int J Tuberc Lung Dis* 2015; **19**: 525–30.

249 Calligaro GL, Theron G, Khalfey H, *et al.* Burden of tuberculosis in intensive care units in Cape Town, South Africa, and assessment of the accuracy and effect on patient outcomes of the Xpert MTB/RIF test on tracheal aspirate samples for diagnosis of pulmonary tuberculosis: a prospective burden o. *Lancet Respir Med* 2015; **3**: 621–30.

250 Igari H, Imasawa T, Noguchi N, *et al.* Advanced stage of chronic kidney disease is risk of poor treatment outcome for smear-positive pulmonary tuberculosis. *J Infect Chemother* 2015; **21**: 559–63.

251 Meressa D, Hurtado RM, Andrews JR, *et al.* Achieving high treatment success for multidrug-resistant TB in Africa: initiation and scale-up of MDR TB care in Ethiopia--an observational cohort study. *Thorax* 2015; **70**: 1181–8.

252 Hang NTL, Maeda S, Keicho N, Thuong PH, Endo H. Sublineages of Mycobacterium tuberculosis Beijing genotype strains and unfavorable outcomes of anti-tuberculosis treatment. *Tuberculosis* 2015; **95**: 336–42.

253 Ukwaja KN, Oshi SN, Alobu I, Oshi DC. Six-vs. eight-month anti-tuberculosis regimen for pulmonary tuberculosis under programme conditions. *Int J Tuberc Lung Dis* 2015; **19**: 295–301.

254 Zhang Q, Wu Z, Zhang Z, Sha W, Shen X, Xiao H. Efficacy and effect of free treatment on multidrug-resistant tuberculosis. *Exp Ther Med* 2016; **11**: 777–82.

255 Huyen MN, Buu TN, Tiemersma E, *et al.* Tuberculosis relapse in Vietnam is significantly associated with Mycobacterium tuberculosis Beijing genotype infections. *J Infect Dis* 2013; **207**: 1516–24.

256 Park HO, Kim SH, Moon SH, *et al.* Association between body mass index and sputum culture conversion among South Korean patients with multidrug resistant tuberculosis in a tuberculosis referral hospital. *Infect Chemother* 2016; **48**: 317–23.

257 Nabukenya-Mudiope MG, Kawuma HJ, Brouwer M, Mudiope P, Vassall A. Tuberculosis retreatment 'others' in comparison with classical retreatment cases; a retrospective cohort review. *BMC Public Health* 2015; **15**: 840.

258 Umanah T, Ncayiyana J, Padanilam X, Nyasulu PS. Treatment outcomes in multidrug resistant tuberculosis-human immunodeficiency virus Co-infected patients on anti-retroviral therapy at Sizwe Tropical Disease Hospital Johannesburg, South Africa. *BMC Infect Dis* 2015; **15**: 478.

259 Chien JY, Chen YT, Wu SG, Lee JJ, Wang JY, Yu CJ. Treatment outcome of patients with isoniazid mono-resistant tuberculosis. *Clin Microbiol Infect* 2015; **21**: 59–68.

260 Yamana H, Matsui H, Fushimi K, Yasunaga H. Treatment options and outcomes of hospitalised tuberculosis patients: a nationwide study. *Int J Tuberc Lung Dis* 2015; **19**: 120–6.

261 Dobler CC, Korver S, Batbayar O, *et al.* Success of community-based directly observed anti-tuberculosis treatment in Mongolia. *Int J Tuberc Lung Dis* 2015; **19**: 657–62.

262 Sawadogo B, Tint KS, Tshimanga M, Kuonza L, Ouedraogo L. Risk factors for tuberculosis treatment failure

among pulmonary tuberculosis patients in four health regions of Burkina Faso, 2009: case control study. *Pan Afr Med J* 2015; **21**: 152.

263 Oshi SN, Alobu I, Ukwaja KN, Oshi DC. Investigating gender disparities in the profile and treatment outcomes of tuberculosis in Ebonyi state, Nigeria. *Epidemiol Infect* 2015; **143**: 932–42.

264 Chiang CY, Bai KJ, Lin HH, *et al.* The influence of diabetes, glycemic control, and diabetes-related comorbidities on pulmonary tuberculosis. *PLoS One* 2015; **10**. DOI:10.1371/journal.pone.0121698.

265 Kosgei RJ, Sitienei JK, Kipruto H, *et al.* Gender differences in treatment outcomes among 15-49 year olds with smear-positive pulmonary tuberculosis in Kenya. *Int J Tuberc Lung Dis* 2015; **19**: 1176–81.

266 Yilmaz S, Taylan M, Sen HS, *et al.* Factors associated with mortality among tuberculosis patients in southeast Turkey. *Acta Medica Mediterr* 2015; **31**: 1241–6.

267 Sengul A, Akturk UA, Aydemir Y, Kaya N, Kocak ND, Tasolar FT. Factors affecting successful treatment outcomes in pulmonary tuberculosis: A single-center experience in Turkey, 2005-2011. *J Infect Dev Ctries* 2015; **9**: 821–8.

268 Delgado-Sánchez G, García-García L, Castellanos-Joya M, *et al.* Association of pulmonary tuberculosis and diabetes in Mexico: Analysis of the National Tuberculosis Registry 2000-2012. *PLoS One* 2015; **10**. DOI:10.1371/journal.pone.0129312.

269 Sun Y, Harley D, Vally H, Sleigh A. Comparison of characteristics and mortality in multidrug resistant (MDR) and non-MDR tuberculosis patients in China. *BMC Public Health* 2015; **15**: 1027.

270 Wu YC, Lo HY, Yang SL, Chu DC, Chou P. Comparing the factors correlated with tuberculosis-specific and non-tuberculosis-specific deaths in different age groups among tuberculosis-related deaths in Taiwan. *PLoS One* 2015; **10**: e0118929.

271 Balkema CA, Irusen EM, Taljaard JJ, Koegelenberg CF. Tuberculosis in the intensive care unit: a prospective observational study. *Int J Tuberc Lung Dis* 2014; **18**: 824–30.

272 Lanoix JP, Gaudry S, Flicoteaux R, Ruimy R, Wolff M. Tuberculosis in the intensive care unit: a descriptive analysis in a low-burden country. *Int J Tuberc Lung Dis* 2014; **18**: 581–7.

273 Dangisso MH, Woldesemayat EM, Datiko DG, Lindtjørn B. Long-term outcome of smear-positive tuberculosis patients after initiation and completion of treatment: A ten-year retrospective cohort study. *PLoS One* 2018; **13**: e0193396.

274 Marais E, Mlambo CK, Lewis JJ, *et al.* Treatment outcomes of multidrug-resistant tuberculosis patients in Gauteng, South Africa. *Infection* 2014; **42**: 405–13.

275 Lucenko I, Riekstina V, Perevoscikovs J, *et al.* Treatment outcomes among drug-susceptible tuberculosis patients in Latvia, 2006-2010. *Public Heal Action* 2014; **4**: S54-8.

276 Jain K, Desai M, Solanki R, Dikshit RK. Treatment outcome of standardized regimen in patients with multidrug resistant tuberculosis. *J Pharmacol Pharmacother* 2014; **5**: 145–9.

277 Atif M, Sulaiman SA, Shafie AA, Ali I, Asif M, Babar ZU. Treatment outcome of new smear positive pulmonary tuberculosis patients in Penang, Malaysia. *BMC Infect Dis* 2014; **14**: 399.

278 Pusch T, Pasipanodya JG, Hall 2nd RG, Gumbo T. Therapy duration and long-term outcomes in extra-pulmonary tuberculosis. *BMC Infect Dis* 2014; **14**: 115.

279 Aung KJ, Van Deun A, Declercq E, *et al.* Successful '9-month Bangladesh regimen' for multidrug-resistant tuberculosis among over 500 consecutive patients. *Int J Tuberc Lung Dis* 2014; **18**: 1180–7.

280 Han SH, Zhou J, Lee MP, *et al.* Prognostic significance of the interval between the initiation of antiretroviral

therapy and the initiation of anti-tuberculosis treatment in HIV/tuberculosis-coinfected patients: results from the TREAT Asia HIV Observational Database. *HIV Med* 2014; **15**: 77–85.

281 Oshi DC, Oshi SN, Alobu I, Ukwaja KN. Profile and treatment outcomes of tuberculosis in the elderly in southeastern Nigeria, 2011-2012. *PLoS One* 2014; **9**: e111910.

282 Peltzer K, Louw JS. Prevalence and factors associated with tuberculosis treatment outcome among hazardous or harmful alcohol users in public primary health care in South Africa. *Afr Heal Sci* 2014; **14**: 157–66.

283 Choi H, Lee M, Chen RY, *et al.* Predictors of pulmonary tuberculosis treatment outcomes in South Korea: a prospective cohort study, 2005-2012. *BMC Infect Dis* 2014; **14**: 360.

284 Khaliaukin A, Kumar AM, Skrahina A, *et al.* Poor treatment outcomes among multidrug-resistant tuberculosis patients in Gomel Region, Republic of Belarus. *Public Heal Action* 2014; **4**: S24-8.

285 Kuksa L, Riekstina V, Leimane V, *et al.* Multi- and extensively drug-resistant tuberculosis in Latvia: trends, characteristics and treatment outcomes. *Public Heal Action* 2014; **4**: S47-53.

286 Shuldiner J, Leventhal A, Chemtob D, Mor Z. Mortality of tuberculosis patients during treatment in Israel, 2000-2010. *Int J Tuberc Lung Dis* 2014; **18**: 818–23.

287 Pietersen E, Ignatius E, Streicher EM, *et al.* Long-term outcomes of patients with extensively drug-resistant tuberculosis in South Africa: a cohort study. *Lancet* 2014; **383**: 1230–9.

288 Babalik A, Kiziltas S, Gencer S, Kilicaslan Z. An investigation into the relationship between region specific quality of life and adverse tuberculosis treatment outcomes in Istanbul, Turkey. *Rev Port Pneumol* 2014; **20**: 248–53.

289 Uyei J, Coetzee D, Macinko J, Weinberg SL, Guttmacher S. The influence of integrated tuberculosis and human immunodeficiency virus service delivery on patient outcomes. *Int J Tuberc Lung Dis* 2014; **18**: 315–21.

290 Velásquez GE, Becerra MC, Gelmanova IY, *et al.* Improving outcomes for multidrug-resistant tuberculosis: aggressive regimens prevent treatment failure and death. *Clin Infect Dis* 2014; **59**: 9–15.

291 Baghaei P, Marjani M, Tabarsi P, *et al.* Impact of chronic renal failure on anti-tuberculosis treatment outcomes. *Int J Tuberc Lung Dis* 2014; **18**: 352–6.

292 Djibuti M, Mirvelashvili E, Makharashvili N, Magee MJ. Household income and poor treatment outcome among patients with tuberculosis in Georgia: a cohort study. *BMC Public Health* 2014; **14**: 88.

293 Bastard M, Sanchez-Padilla E, Hewison C, *et al.* Effects of treatment interruption patterns on treatment success among patients with multidrug-resistant tuberculosis in Armenia and Abkhazia. *J Infect Dis* 2015; **211**: 1607–15.

294 Hamusse SD, Demissie M, Teshome D, Lindtjørn B. Fifteen-year trend in treatment outcomes among patients with pulmonary smear-positive tuberculosis and its determinants in Arsi Zone, Central Ethiopia. *Glob Heal Action* 2014; **7**: 25382.

295 Wingfield T, Boccia D, Tovar M, *et al.* Defining catastrophic costs and comparing their importance for adverse tuberculosis outcome with multi-drug resistance: a prospective cohort study, Peru. *PLoS Med* 2014; **11**: e1001675.

296 Putri FA, Burhan E, Nawas A, *et al.* Body mass index predictive of sputum culture conversion among MDR-TB patients in Indonesia. *Int J Tuberc Lung Dis* 2014; **18**: 564–70.

297 Esmael A, Tsegaye G, Wubie M, Abera H, Endris M. Treatment outcomes of tuberculosis patients in Debre Markos Referral Hospital, north west Ethiopia (June 2008-August 2013): A five year retrospective study. *Int J Pharm Sci Res* 2014; **5**: 1500–5.

298 Alobu I, Oshi SN, Oshi DC, Ukwaja KN. Risk factors of treatment default and death among tuberculosis patients in a resource-limited setting. *Asian Pac J Trop Med* 2014; **7**: 977–84.

299 Kwon YS, Kim YH, Song JU, *et al.* Risk factors for death during pulmonary tuberculosis treatment in Korea: a multicenter retrospective cohort study. *J Korean Med Sci* 2014; **29**: 1226–31.

300 Alobu I, Oshi DC, Oshi SN, Ukwaja KN. Profile and determinants of treatment failure among smear-positive pulmonary tuberculosis patients in Ebonyi, Southeastern Nigeria. *Int J Mycobacteriology* 2014; **3**: 127–31.

301 Pefura-Yone EW, Kengne AP, Kuaban C. Non-conversion of sputum culture among patients with smear positive pulmonary tuberculosis in Cameroon: A prospective cohort study. *BMC Infect Dis* 2014; **14**. DOI:10.1186/1471-2334-14-138.

302 Ershova J V, Kurbatova E V, Moonan PK, Cegielski JP. Mortality among tuberculosis patients with acquired resistance to second-line antituberculosis drugs - United States, 1993-2008. *Clin Infect Dis* 2014; **59**: 465–72.

303 Iype T, Pillai AK, Cherian A, *et al.* Major outcomes of patients with tuberculous meningitis on directly observed thrice a week regime. *Ann Indian Acad Neurol* 2014; **17**: 281–6.

304 Viswanathan AA, Gawde NC. Effect of type II diabetes mellitus on treatment outcomes of tuberculosis. *Lung India* 2014; **31**: 244–8.

305 Alo A, Gounder S, Graham SM, Graham SM. Clinical characteristics and treatment outcomes of tuberculosis cases hospitalised in the intensive phase in Fiji. *Public Heal Action* 2014; **4**: 164–8.

306 Przybylski G, Dabrowska A, Trzcińska H. Alcoholism and other socio-demographic risk factors for adverse tb-drug reactions and unsuccessful tuberculosis treatment - Data from ten years' observation at the regional centre of pulmonology, bydgoszcz, Poland. *Med Sci Monit* 2014; **20**: 444–53.

307 O'Donnell MR, Padayatchi N, Kvasnovsky C, Werner L, Master I, Horsburgh CR. Treatment outcomes for extensively drug-resistant tuberculosis and HIV Co-infection. *Emerg Infect Dis* 2013; **19**: 416–24.

308 Getahun B, Ameni G, Medhin G, Biadgilign S. Treatment outcome of tuberculosis patients under directly observed treatment in Addis Ababa, Ethiopia. *Braz J Infect Dis* 2013; **17**: 521–8.

309 Anderson LF, Tamne S, Watson JP, *et al.* Treatment outcome of multi-drug resistant tuberculosis in the United Kingdom: retrospective-prospective cohort study from 2004 to 2007. *Euro Surveill* 2013; **18**. DOI:10.2807/1560-7917.es2013.18.40.20601.

310 Manda SO, Masenyetse LJ, Lancaster JL, van der Walt ML. Risk of Death among HIV Co-Infected Multidrug Resistant Tuberculosis Patients, Compared To Mortality in the General Population of South Africa. *J AIDS Clin Res* 2013; **Suppl 3**: 7.

311 Babalik A, Kilicaslan Z, Caner SS, *et al.* A registry-based cohort study of pulmonary tuberculosis treatment outcomes in Istanbul, Turkey. *Jpn J Infect Dis* 2013; **66**: 115–20.

312 Patra S, Lukhmana S, Tayler Smith K, *et al.* Profile and treatment outcomes of elderly patients with tuberculosis in Delhi, India: implications for their management. *Trans R Soc Trop Med Hyg* 2013; **107**: 763–8.

313 Horita N, Miyazawa N, Yoshiyama T, *et al.* Poor performance status is a strong predictor for death in patients with smear-positive pulmonary TB admitted to two Japanese hospitals. *Trans R Soc Trop Med Hyg* 2013; **107**: 451–6.

314 Blöndal K, Rahu K, Altraja A, Viiklepp P, Rahu M. Overall and cause-specific mortality among patients with tuberculosis and multidrug-resistant tuberculosis. *Int J Tuberc Lung Dis* 2013; **17**: 961–8.

315 Schwartz AB, Tamuhla N, Steenhoff AP, *et al.* Outcomes in HIV-infected adults with tuberculosis at clinics with and without co-located HIV clinics in Botswana. *Int J Tuberc Lung Dis* 2013; **17**: 1298–303.

316 Abouzeid MS, Al RF, Memish ZA. Mortality among tuberculosis patients in Saudi Arabia (2001-2010). *Ann Saudi Med* 2013; **33**: 247–52.

317 Kabali C, Mtei L, Brooks DR, *et al.* Increased mortality associated with treated active tuberculosis in HIV-infected adults in Tanzania. *Tuberc* 2013; **93**: 461–6.

318 Deepa D, Achanta S, Jaju J, *et al.* The impact of isoniazid resistance on the treatment outcomes of smear positive re-treatment tuberculosis patients in the state of Andhra Pradesh, India. *PLoS One* 2013; **8**: e76189.

319 Kang YA, Kim SY, Jo KW, *et al.* Impact of diabetes on treatment outcomes and long-term survival in multidrug-resistant tuberculosis. *Respiration* 2013; **86**: 472–8.

320 Sulaiman SAS, Khan AH, Ahmad N, Iqubal MS, Muttalif AR, Hassali MA. Impact of diabetes mellitus on treatment outcomes of tuberculosis patients in tertiary care setup. *Am J Med Sci* 2013; **345**: 321–5.

321 Reed GW, Choi H, Lee SY, *et al.* Impact of diabetes and smoking on mortality in tuberculosis. *PLoS One* 2013; **8**: e58044.

322 Nakanwagi-Mukwaya A, Reid AJ, Fujiwara PI, *et al.* Characteristics and treatment outcomes of tuberculosis retreatment cases in three regional hospitals, Uganda. *Public Heal Action* 2013; **3**: 149–55.

323 Uchimura K, Ngamvithayapong-Yanai J, Kawatsu L, *et al.* Characteristics and treatment outcomes of tuberculosis cases by risk groups, Japan, 2007-2010. *West Pac Surveill Response J* 2013; **4**: 11–8.

324 Hoa NB, Lauritsen JM, Rieder HL. Changes in body weight and tuberculosis treatment outcome in Viet Nam. *Int J Tuberc Lung Dis* 2013; **17**: 61–6.

325 Ugarte-Gil C, Ruiz P, Zamudio C, *et al.* Association of major depressive episode with negative outcomes of tuberculosis treatment. *PLoS One* 2013; **8**: e69514.

326 Mitnick CD, Franke MF, Rich ML, *et al.* Aggressive regimens for multidrug-resistant tuberculosis decrease all-cause mortality. *PLoS One* 2013; **8**: e58664.

327 Gler MT, Guilatco R, Caoili JC, Ershova J, Cegielski P, Johnson JL. Weight Gain and response to treatment for multidrug-resistant tuberculosis. *Am J Trop Med Hyg* 2013; **89**: 943–9.

328 Kenangalem E, Waramori G, Pontororing GJ, *et al.* Tuberculosis Outcomes in Papua, Indonesia: The Relationship with Different Body Mass Index Characteristics between Papuan and Non-Papuan Ethnic Groups. *PLoS One* 2013; **8**. DOI:10.1371/journal.pone.0076077.

329 Ribeiro Macedo L, Reis-Santos B, Riley LW, Maciel EL. Treatment outcomes of tuberculosis patients in Brazilian prisons: A polytomous regression analysis. *Int J Tuberc Lung Dis* 2013; **17**: 1427–34.

330 Antoine D, Che D. Treatment outcome monitoring of pulmonary tuberculosis cases notified in France in 2009. *Eurosurveillance* 2013; **18**.

331 Tang S, Tan S, Yao L, *et al.* Risk factors for poor treatment outcomes in patients with MDR-TB and XDR-TB in China: Retrospective multi-center investigation. *PLoS One* 2013; **8**. DOI:10.1371/journal.pone.0082943.

332 Ananthakrishnan R, Kumar K, Ganesh M, *et al.* The Profile and Treatment Outcomes of the Older (Aged 60 Years and Above) Tuberculosis Patients in Tamilnadu, South India. *PLoS One* 2013; **8**. DOI:10.1371/journal.pone.0067288.

333 Pazarli P, Duman DY, Moçin Ö Y, Karagöz T. The effect of smoking on treatment outcome of multidrug-resistant tuberculosis. *Turk Toraks Derg* 2013; **14**: 93–7.

334 Tweya H, Feldacker C, Phiri S, *et al.* Comparison of treatment outcomes of new smear-positive pulmonary tuberculosis patients by HIV and antiretroviral status in a TB/HIV clinic, Malawi. *PLoS One* 2013; **8**. DOI:10.1371/journal.pone.0056248.

335 Yen YF, Yen MY, Lin YS, *et al.* Smoking increases risk of recurrence after successful anti-tuberculosis treatment: a population-based study. *Int J Tuberc Lung Dis* 2014; **18**: 492–8.

336 Mpagama SG, Heysell SK, Ndusilo ND, *et al.* Diagnosis and interim treatment outcomes from the first cohort of multidrug-resistant tuberculosis patients in Tanzania. *PLoS One* 2013; **8**: e62034.

337 Ismail I, Bulgiba A. Determinants of unsuccessful tuberculosis treatment outcomes in Malaysian HIV-infected patients. *Prev Med* 2013; **57 Suppl**: S27-30.

338 Wang WB, Zhao Q, Yuan ZA, Jiang WL, Liu ML, Xu B. Deaths of tuberculosis patients in urban China: a retrospective cohort study. *Int J Tuberc Lung Dis* 2013; **17**: 493–8.

339 Shaweno D, Worku A. Tuberculosis treatment survival of HIV positive TB patients on directly observed treatment short-course in Southern Ethiopia: a retrospective cohort study. *BMC Res Notes* 2012; **5**: 682.

340 Limmahakhun S, Chaiwarith R, Nuntachit N, Sirisanthana T, Supparatpinyo K. Treatment outcomes of patients co-infected with tuberculosis and HIV at Chiang Mai University Hospital, Thailand. *Int J STD AIDS* 2012; **23**: 414–8.

341 Takarinda KC, Harries AD, Srinath S, Mutasa-Apollo T, Sandy C, Mugurungi O. Treatment outcomes of adult patients with recurrent tuberculosis in relation to HIV status in Zimbabwe: a retrospective record review. *BMC Public Health* 2012; **12**: 124.

342 Hoa NB, Sokun C, Wei C, Lauritsen JM, Rieder HL. Time to unsuccessful tuberculosis treatment outcome, Cambodia, China, and Viet Nam. *Public Heal Action* 2012; **2**: 15–20.

343 Cavanaugh JS, Kazennyy BY, Nguyen ML, *et al.* Outcomes and follow-up of patients treated for multidrug-resistant tuberculosis in Orel, Russia, 2002-2005. *Int J Tuberc Lung Dis* 2012; **16**: 1069–74.

344 Bloss E, Chan PC, Cheng NW, Wang KF, Yang SL, Cegielski P. Increasing directly observed therapy related to improved tuberculosis treatment outcomes in Taiwan. *Int J Tuberc Lung Dis* 2012; **16**: 462–7.

345 Kassa A, Teka A, Shewaamare A, Jerene D. Incidence of tuberculosis and early mortality in a large cohort of HIV infected patients receiving antiretroviral therapy in a tertiary hospital in Addis Ababa, Ethiopia. *Trans R Soc Trop Med Hyg* 2012; **106**: 363–70.

346 Girardi E, Palmieri F, Angeletti C, *et al.* Impact of previous ART and of ART initiation on outcome of HIV-associated tuberculosis. *Clin Dev Immunol* 2012; **2012**: 931325.

347 Palacios E, Franke M, Muñoz M, *et al.* HIV-positive patients treated for multidrug-resistant tuberculosis: clinical outcomes in the HAART era. *Int J Tuberc Lung Dis* 2012; **16**: 348–54.

348 Feng JY, Huang SF, Ting WY, *et al.* Gender differences in treatment outcomes of tuberculosis patients in Taiwan: a prospective observational study. *Clin Microbiol Infect* 2012; **18**: E331-7.

349 Mukherjee A, Saha I, Sarkar A, Chowdhury R. Gender differences in notification rates, clinical forms and treatment outcome of tuberculosis patients under the RNTCP. *Lung India* 2012; **29**: 120–2.

350 Chirwa T, Nyasulu P, Chirwa E, *et al.* Levels of Tuberculosis Treatment Adherence among Sputum Smear Positive Pulmonary Tuberculosis Patients Attending Care at Zomba Central Hospital, Southern Malawi, 2007-2008. *PLoS One* 2013; **8**. DOI:10.1371/journal.pone.0063050.

351 Mnisi T, Tumbo J, Govender I. Factors associated with pulmonary tuberculosis outcomes among inmates in Potchefstroom Prison in North West province. *South African J Epidemiol Infect* 2013; **28**: 96–101.

352 Riou C, Perez Peixoto B, Roberts L, *et al.* Effect of standard tuberculosis treatment on plasma cytokine levels in patients with active pulmonary tuberculosis. *PLoS One* 2012; **7**: e36886.

353 Janols H, Abate E, Idh J, *et al.* Early treatment response evaluated by a clinical scoring system correlates with

the prognosis of pulmonary tuberculosis patients in Ethiopia: a prospective follow-up study. *Scand J Infect Dis* 2012; **44**: 828–34.

354 Koh WJ, Kang YR, Jeon K, *et al.* Daily 300 mg dose of linezolid for multidrug-resistant and extensively drug-resistant tuberculosis: updated analysis of 51 patients. *J Antimicrob Chemother* 2012; **67**: 1503–7.

355 Yee DP, Menzies D, Brassard P. Clinical outcomes of pyrazinamide-monoresistant Mycobacterium tuberculosis in Quebec. *Int J Tuberc Lung Dis* 2012; **16**: 604–9.

356 Kattan JA, Sosa LE, Lobato MN. Tuberculosis mortality: Death from a curable disease, Connecticut, 2007-2009. *Int J Tuberc Lung Dis* 2012; **16**: 1657–62.

357 Valade S, Raskine L, Aout M, *et al.* Tuberculosis in the intensive care unit: A retrospective descriptive cohort study with determination of a predictive fatality score. *Can J Infect Dis Med Microbiol* 2012; **23**: 173–8.

358 Tabarsi P, Chitsaz E, Moradi A, *et al.* Treatment outcome, mortality and their predictors among HIV-associated tuberculosis patients. *Int J STD AIDS* 2012; **23**: e1–4.

359 Van't Hoog AH, Williamson J, Sewe M, *et al.* Risk factors for excess mortality and death in adults with tuberculosis in Western Kenya. *Int J Tuberc Lung Dis* 2012; **16**: 1649–56.

360 Gegia M, Cohen T, Kalandadze I, Vashakidze L, Furin J. Outcomes among tuberculosis patients with isoniazid resistance in Georgia, 2007-2009. *Int J Tuberc Lung Dis* 2012; **16**: 812–6.

361 Tabarsi P, Chitsaz E, Moradi A, *et al.* Treatment outcome, mortality and their predictors among HIV-associated tuberculosis patients. *Int J STD AIDS* 2012; **23**: e1–4.

362 Belo MT, Luiz RR, Teixeira EG, Hanson C, Trajman A. Tuberculosis treatment outcomes and socio-economic status: a prospective study in Duque de Caxias, Brazil. *Int J Tuberc Lung Dis* 2011; **15**: 978–81.

363 Srinath S, Sharath B, Santosha K, *et al.* Tuberculosis 'retreatment others': profile and treatment outcomes in the state of Andhra Pradesh, India. *Int J Tuberc Lung Dis* 2011; **15**: 105–9.

364 Takarinda KC, Harries AD, Srinath S, Mutasa-Apollo T, Sandy C, Mugurungi O. Treatment outcomes of new adult tuberculosis patients in relation to HIV status in Zimbabwe. *Public Heal Action* 2011; **1**: 34–9.

365 Jeon DS, Shin DO, Park SK, *et al.* Treatment outcome and mortality among patients with multidrug-resistant tuberculosis in tuberculosis hospitals of the public sector. *J Korean Med Sci* 2011; **26**: 33–41.

366 Jonnalagada S, Harries AD, Zachariah R, *et al.* The timing of death in patients with tuberculosis who die during anti-tuberculosis treatment in Andhra Pradesh, South India. *BMC Public Health* 2011; **11**: 921.

367 Dooley KE, Lahlou O, Ghali I, *et al.* Risk factors for tuberculosis treatment failure, default, or relapse and outcomes of retreatment in Morocco. *BMC Public Health* 2011; **11**: 140.

368 Farley JE, Ram M, Pan W, *et al.* Outcomes of multi-drug resistant tuberculosis (MDR-TB) among a cohort of South African patients with high HIV prevalence. *PLoS One* 2011; **6**: e20436.

369 Bendayan D, Hendler A, Polansky V, Weinberger M. Outcome of hospitalized MDR-TB patients: Israel 2000-2005. *Eur J Clin Microbiol Infect Dis* 2011; **30**: 375–9.

370 Christensen AS, Roed C, Omland LH, Andersen PH, Obel N, Andersen Å B. Long-term mortality in patients with tuberculous meningitis: a Danish nationwide cohort study. *PLoS One* 2011; **6**: e27900.

371 Visser ME, Stead MC, Walzl G, *et al.* Baseline predictors of sputum culture conversion in pulmonary tuberculosis: importance of cavities, smoking, time to detection and W-Beijing genotype. *PLoS One* 2012; **7**: e29588.

372 Arentz M, Narita M, Sangaré L, *et al.* Impact of smear microscopy results and observed therapy on tuberculosis treatment in Mombasa, Kenya. *Int J Tuberc Lung Dis* 2011; **15**: 1656–63.

373 Nahid P, Jarlsberg LG, Rudoy I, *et al.* Factors associated with mortality in patients with drug-susceptible pulmonary tuberculosis. *BMC Infect Dis* 2011; **11**: 1.

374 Burton NT, Forson A, Lurie MN, Kudzawu S, Kwarteng E, Kwara A. Factors associated with mortality and default among patients with tuberculosis attending a teaching hospital clinic in Accra, Ghana. *Trans R Soc Trop Med Hyg* 2011; **105**: 675–82.

375 Blanc FX, Sok T, Laureillard D, *et al.* Earlier versus later start of antiretroviral therapy in HIV-infected adults with tuberculosis. *N Engl J Med* 2011; **365**: 1471–81.

376 Jung RS, Bennion JR, Sorvillo F, Bellomy A. Trends in tuberculosis mortality in the United States, 1990-2006: a population-based case-control study. *Public Heal Rep* 2010; **125**: 389–97.

377 Kim DH, Kim HJ, Park SK, *et al.* Treatment outcomes and survival based on drug resistance patterns in multidrug-resistant tuberculosis. *Am J Respir Crit Care Med* 2010; **182**: 113–9.

378 Vasankari T, Holmström P, Ollgren J, Liippo K, Ruutu P. Treatment outcome of extra-pulmonary tuberculosis in Finland: a cohort study. *BMC Public Health* 2010; **10**: 399.

379 Misra UK, Kalita J, Nair PP. Role of aspirin in tuberculous meningitis: a randomized open label placebo controlled trial. *J Neurol Sci* 2010; **293**: 12–7.

380 Wen CP, Chan TC, Chan HT, Tsai MK, Cheng TY, Tsai SP. The reduction of tuberculosis risks by smoking cessation. *BMC Infect Dis* 2010; **10**: 156.

381 Hsu PC, Yang CC, Ye JJ, Huang PY, Chiang PC, Lee MH. Prognostic factors of tuberculous meningitis in adults: a 6-year retrospective study at a tertiary hospital in northern Taiwan. *J Microbiol Immunol Infect* 2010; **43**: 111–8.

382 Sassaki CM, Scatena LM, Gonzales RI, Ruffino-Netto A, Hinos P, Villa TC. Predictors of favorable results in pulmonary tuberculosis treatment (Recife, Pernambuco, Brazil, 2001-2004). *Rev Esc Enferm USP* 2010; **44**: 504–10.

383 Ferrer G, Acuna-Villaorduna C, Escobedo M, Vlasich E, Rivera M. Outcomes of multidrug-resistant tuberculosis among binational cases in El Paso, Texas. *Am J Trop Med Hyg* 2010; **83**: 1056–8.

384 Buu TN, Huyen MN, van Soolingen D, *et al.* The Mycobacterium tuberculosis Beijing genotype does not affect tuberculosis treatment failure in Vietnam. *Clin Infect Dis* 2010; **51**: 879–86.

385 Silva DR, Menegotto DM, Schulz LF, Gazzana MB, Dalcin PT. Mortality among patients with tuberculosis requiring intensive care: a retrospective cohort study. *BMC Infect Dis* 2010; **10**: 54.

386 Shaw JE, Pasipanodya JG, Gumbo T. Meningeal tuberculosis: high long-term mortality despite standard therapy. *Med* 2010; **89**: 189–95.

387 Vinnard C, Winston CA, Wileyto EP, Macgregor RR, Bisson GP. Isoniazid resistance and death in patients with tuberculous meningitis: retrospective cohort study. *Bmj* 2010; **341**: c4451.

388 Uwizeye CB, De Serres G, Gilca R, Schwartzman K, Gasana M. Tuberculosis may be underestimated in Rwandan women. *Int J Tuberc Lung Dis* 2011; **15**: 776-781+i.

389 Jacobson KR, Theron D, Victor TC, Streicher EM, Warren RM, Murray MB. Treatment outcomes of isoniazid-resistant tuberculosis patients, Western Cape Province, South Africa. *Clin Infect Dis* 2011; **53**: 369–72.

390 Joseph N, Nagaraj K, Bhat J, *et al.* Treatment outcomes among new smear positive and retreatment cases of tuberculosis in Mangalore, South India - a descriptive study. *Australas Med J* 2011; **4**: 162–7.

391 Fawibe AE, Salami AK, Oluboyo PO, Desalu OO, Odeigha LO. Profile and outcome of unilateral tuberculous lung destruction in Ilorin, Nigeria. *West Afr J Med* 2011; **30**: 130–5.

392 Marais S, Pepper DJ, Schutz C, Wilkinson RJ, Meintjes G. Presentation and outcome of tuberculous meningitis in a high HIV prevalence setting. *PLoS One* 2011; **6**. DOI:10.1371/journal.pone.0020077.

393 Horne DJ, Hubbard R, Narita M, Exarchos A, Park DR, Goss CH. Factors associated with mortality in patients with tuberculosis. *BMC Infect Dis* 2010; **10**: 258.

394 Silva DR, Menegotto DM, Schulz LF, Gazzana MB, Dalcin Pde T. Factors associated with mortality in hospitalized patients with newly diagnosed tuberculosis. *Lung* 2010; **188**: 33–41.

395 Leimane V, Dravniece G, Riekstina V, *et al.* Treatment outcome of multidrug/extensively drug-resistant tuberculosis in Latvia, 2000-2004. *Eur Respir J* 2010; **36**: 584–93.

396 Dheda K, Shean K, Zumla A, *et al.* Early treatment outcomes and HIV status of patients with extensively drug-resistant tuberculosis in South Africa: a retrospective cohort study. *Lancet* 2010; **375**: 1798–807.

397 Senkoro M, Mfinanga SG, Mørkve O. Smear microscopy and culture conversion rates among smear positive pulmonary tuberculosis patients by HIV status in Dar es Salaam, Tanzania. *BMC Infect Dis* 2010; **10**: 210.

398 Chidambaram V, Gupte A, Wang J-Y, Golub JE, Karakousis PC. The Impact of Hypertension and Use of Calcium Channel Blockers on Tuberculosis Treatment Outcomes. *Clin Infect Dis* 2020; published online Sept 24. DOI:10.1093/cid/ciaa1446.

399 Moher D, Shamseer L, Clarke M, *et al.* Preferred reporting items for systematic review and meta-analysis protocols (PRISMA-P) 2015 statement. *Rev Esp Nutr Humana y Diet* 2016; **20**: 148–60.

400 Definitions and reporting framework for tuberculosis-2013 revision. .

401 Covidence systematic review software, Veritas Health Innovation, Melbourne AA at, Www.covidence.org. No Title. .

402 2020 Q (2020) Q com. A at: http://www. qualtrics. com. (accessed 24 march. No Title. .

403 Chapter 6: Choosing effect measures and computing estimates of effect | Cochrane Training. https://training.cochrane.org/handbook/current/chapter-06 (accessed Jan 27, 2021).

404 Ottawa Hospital Research Institute. http://www.ohri.ca/programs/clinical_epidemiology/oxford.asp (accessed Jan 28, 2021).

405 World Bank Country and Lending Groups – World Bank Data Help Desk. https://datahelpdesk.worldbank.org/knowledgebase/articles/906519-world-bank-country-and-lending-groups (accessed Dec 24, 2020).

406 Use of high burden country lists for TB by WHO in the post-2015 era meeting of WHO’s Strategic and Technical Advisory Group for TB (STAG-TB). 2015.
